# Mathematical assessment of the impact of the R21/Matrix-M vaccine on the control of malaria in children in Burkina Faso

**DOI:** 10.64898/2026.01.25.26344791

**Authors:** Arnaja Mitra, Abba B. Gumel

**Affiliations:** Department of Mathematics, University of Maryland, College Park, MD, 20742, USA; Institute for Health Computing, University of Maryland, North Bethesda, Maryland, 20852, USA; Department of Mathematics and Applied Mathematics, University of Pretoria, Pretoria 0002, South Africa

**Keywords:** Malaria transmission, R21/Matrix-M vaccine, Age-structured model, Dose-structured vaccination, Delay differential equations, Reproduction number, Vaccine-induced herd immunity threshold

## Abstract

This study is based on the design and analysis of a novel age- and dose-structured model for assessing the population-level impact of the recently-approved R21/Matrix-M malaria vaccine (which is administered in three doses followed by a booster dose) on controlling the spread of malaria in children under five in Burkina Faso. While the current malaria vaccination program in Burkina Faso prioritizes children 0–3 years of age (Group 1 in our model), we also assessed a hypothetical scenario where children 3–5 years of age (Group 2 in our model) are also vaccinated (since children under five years of age suffer the brunt of malaria morbidity and mortality). The vaccination-free version of the model was calibrated using yearly cumulative malaria mortality data for children in Burkina Faso. In addition to establishing well-posedness, we showed that the disease-free equilibrium of the model is locally-asymptotically stable whenever the control reproduction number (ℛ_*v*_) is below one. Conditions for achieving vaccine-induced *herd immunity* (needed for disease elimination) under varying age-group structures and dosage schedules were derived, and a global sensitivity analysis was conducted to identify the parameters of the model that most strongly influence ℛ_*v*_. Simulations of a homogeneous model including only Group 1 indicate that administering only the first dose of the vaccine with baseline bednet usage requires an impractically high herd immunity threshold of 97%. However, with all four doses, herd immunity is achievable without bednet when the required coverage ratios receiving doses 2, 3, and the booster dose are 73% to 90%. With baseline bednets, these ratios drop to just 10%–30%, dramatically improving elimination prospects. In a heterogeneous setting incorporating both Groups 1 and 2, herd immunity can be achieved (with bednet at baseline) by vaccinating either 46% of the total population of Groups 1 and 2 or 75% of individuals in Group 1 alone. Simulations of the full two-group model (with bednet at baseline) show that vaccinating only children in Group 1 with the first dose reduces the cumulative number of new malaria cases and malaria-induced deaths in Group 1 by about 19%–20%, and produces *spillover* reductions of about 11%–12% in the unvaccinated Group 2, indicating a moderate indirect benefit across groups. If children in Group 1 receive all four doses, the reductions in Group 1 increase to about 36%–38%, with larger spillover reductions of about 25%–26% in Group 2. When both groups receive only the first dose, the malaria burden decreases by about 24%–26% in each group. The greatest reductions occur when both groups receive all four doses, yielding decreases of about 43%–46%. These results show that extending Burkina Faso’s current vaccination program to include children in the 3–5-year age group can substantially improve malaria elimination prospects, particularly when combined with bednet usage at baseline levels or higher.

## 1 Introduction

Despite recent progress, malaria, a mosquito-borne disease caused by the *Plasmodium* parasite and transmitted to humans *via* the effective bites of infectious adult female *Anopheles* mosquitoes, continues to impose a major public health burden in endemic areas (causing an estimated 282 million cases and 610, 000 deaths annually, with about 75% of the deaths occurring in children under the age of five in 2024, according to the World Malaria Report 2025 [1]). Of the five *Plasmodium* parasites (*P. falciparum, P. vivax, P. malariae, P. ovale*, and *P. knowlesi*) that cause malaria in humans, *P. falciparum* is the deadliest [2] (accounting for over 90% of global malaria mortality [3, 4]) and most prevalent in Africa [2] (which bears the brunt of malaria burden globally). *Anopheles gambiae* is the primary vector for *P. falciparum* [5].

Although dramatic success has been recorded in the fight against malaria over the last two decades, owing to the large scale and widespread use of chemical insecticides (mostly in the form of long-lasting insecticidal nets and indoor residual spraying [3, 6–14]) to kill the malaria mosquito, these measures have not been fully effective in eliminating the disease, especially in regions with moderate to high malaria transmission rates. Furthermore, such widespread and heavy use of chemical insecticides has resulted in widespread *Anopheles* resistance to all the chemical insecticides used in vector control [8–10, 12, 13, 15–18], threatening global malaria control efforts (such as the United Nations Sustainable Development Goal to reduce the burden of malaria by 90% by 2030, and the ZERO by 40 initiative aimed at malaria eradication by 2040 [19–22]). There is also the problem of emerging resistance to the anti-malaria drug (*artemisinin*) therapy [23–27]. All of these challenges suggest an urgent need to develop new strategies to add to the arsenal of control resources against the disease. The recent approval of two malaria vaccines, RTS,S/AS01, and R21/Matrix-M, by the World Health Organization (WHO) represents a pivotal step toward addressing this issue.

The malaria parasite’s life cycle [28, 29] involves two hosts: humans and mosquitoes. During a blood meal, an infectious female *Anopheles* mosquito injects sporozoites into a human, which infect liver cells and mature into schizonts (i.e., exo-erythrocytic cycle). These release merozoites that infect red blood cells, leading to asexual multiplication and clinical symptoms (i.e., erythrocytic cycle). Some parasites develop into sexual forms (gametocytes), which mosquitoes ingest during another blood meal. Inside the mosquito, the parasites multiply and form sporozoites, which migrate to the mosquito’s salivary glands (i.e., sporogonic cycle), ready to infect a new human host (Figure 1 depicts the life cycle of the malaria parasite in the two hosts). Scientists have been working on the development of effective malaria vaccines, dating back to the pioneering work of Nussenzweig in 1965 [30] and Clyde et al. in the 1970s [30]. These vaccines are typically categorized based on which of the aforementioned parasite life cycle stages (pre-erythrocytic (human liver stage), erythrocytic (human blood stage), and transmission-blocking (mosquito stage)) they target [30]. The first malaria vaccine, RTS,S/AS01 (Mosquirix), developed by GlaxoSmithKline [31], was approved by the WHO on October 6, 2021 [32]. This vaccine, administered in four doses (costing an average of $9.81 per dose [31], shows protective efficacy of 56% over one year and 36% over four years [30]). A more effective vaccine, R21/Matrix-M, developed by the Jenner Institute, University of Oxford and produced by the Serum Institute of India Private Limited (SIIPL) [33], was approved by the WHO in December of 2023 [34]. This is also a four-dose vaccine, and shows 75% efficacy when administered at seasonal sites (areas where malaria transmission is highly seasonal, and vaccines are given before the malaria season begins) and 68% efficacy when administered at any time of the year once children reach the relevant age [33]. The two approved vaccines target the exo-erythrocytic cycle (i.e., both RTS,S/AS01 and R21/Matrix-M are pre-erythrocytic vaccines [30]; vaccines that target erythrocytic cycle and sporogonic cycle are in various stages of clinical trials [30]). While RTS,S/AS01 has been deployed in endemic areas since 2023 [35], R21/Matrix-M is just being rolled out in a few countries in sub-Saharan Africa, including Burkina Faso [35–37]. Since the vaccines are only being rolled out, it is instructive to use mathematical modeling approaches to assess their potential population-level impact on curtailing the spread of malaria in endemic areas. This forms the main objective of the current study, with emphasis on studying the impact of the vaccination program on malaria transmission in Burkina Faso.

**Figure 1.**
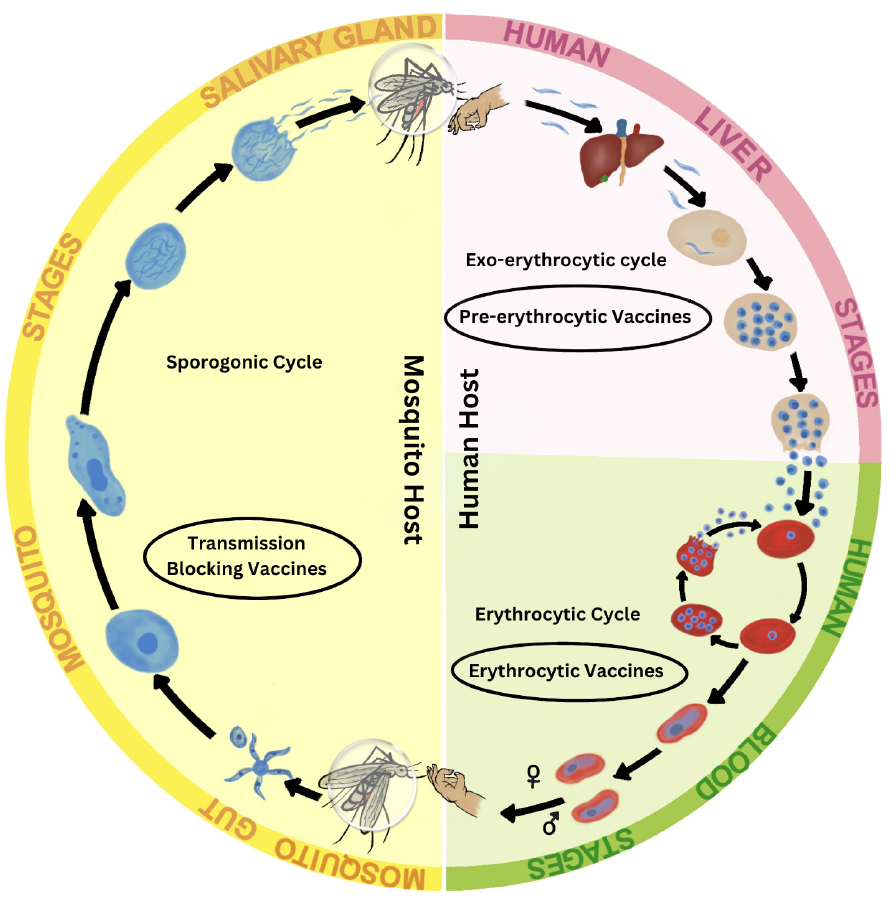
Life cycles of *Plasmodium falciparum* and malaria transmission within the human and mosquito host populations.

Several mathematical models have been developed and used to assess the population-level impact of vaccination programs on malaria dynamics. Some of these models account for key aspects of malaria transmission dynamics, such as boosting of malaria immunity by repeated exposure, seasonality, and intervention strategies against both the vector and the disease [38–43]. For instance, Niger and Gumel [38] developed and used a within-host compartmental malaria model that incorporates human immune responses (cellular and humoral) and an imperfect vaccine, and derived the conditions needed for parasite elimination in the human host. Wenger and Eckhoff [39] employed an agent-based model (using the Epidemiological Modeling Software (known as EMOD)) to quantify vaccine efficacy thresholds and identify optimal deployment strategies within multi-intervention frameworks across varying transmission settings. Thompson et al. [40] also used an agent-based model to demonstrate that seasonally-targeted RTS,S/AS01 vaccination, particularly when combined with chemo-prevention, outperforms age-based anti-malaria vaccination strategies in high-transmission areas. Likewise, Schmit et al. [41] used an agent-based model to evaluate the cost-effectiveness and public health impact of introducing the R21/Matrix-M vaccine across sub-Saharan Africa, showing substantial reductions in malaria burden. Duve et al. [42] used a compartmental model incorporating infected immigrants and vaccination (using RTS,S/AS01) to assess the feasibility of malaria elimination in the South African region, showing that vaccination alone cannot eliminate malaria if malaria-infected immigrants continue to enter the population. The Duve et al. study also showed that, for the case when malaria-infected immigrants are not allowed into the population, the most effective (optimal) study is one that combines vaccination, treatment, and protective interventions (e.g., insecticide-treated nets (ITNs), repellents, insecticide spraying), in line with many other modeling studies for malaria dynamics that include these (multiple) intervention and mitigation strategies [44–46]. Finally, Qu et al. [43] used an age-structured partial differential equation (PDE) model, coupled with ordinary differential equations (ODEs), to analyze malaria vaccination strategies under seasonal malaria transmission. Their study incorporates age-structured PDEs for humans (including a compartment of vaccinated individuals who gain pre-erythrocytic, RTS,S-derived immunity) and modeled anti-malaria immunity dynamics (i.e., immunity acquired through exposure combined with maternal antibodies), along with ODEs for mosquito dynamics. The framework evaluated seasonal versus year-round vaccination strategies for RTS,S/AS01, reporting that, among children under 2 years of age, seasonal vaccination (using the first three doses of RTS,S) can prevent approximately 9% more cases per vaccination than year-round approaches in highly seasonal malaria transmission settings.

The objective of the current study is to develop and use a novel deterministic model for assessing the population-level impact of vaccination as a key intervention, alongside the use of traditional vector-based control strategies (such as the use of insecticide-treated bednet), on the transmission dynamics and control of malaria in Burkina Faso, a malaria-endemic country in Western Africa (and one of the main sites for the clinical trials for R21/Matrix-M in sub-Saharan Africa [33, 47, 48]). Burkina Faso is one of the 20 countries in sub-Saharan Africa currently deploying the R21/Matrix-M vaccine (administered in three doses four weeks apart, and a booster dose, given one year after the third dose) to children (of age between five months and three years). Specifically, the model will incorporate age structure (where the total population of individuals most vulnerable to malaria, notably children under the age of five, is stratified into the vaccine-eligible age group of five months to 3 years, and a second group of children 3 to 5 years of age) and dose-structure (accounting for the aforementioned dose structure). The model, which will take the form of a deterministic system of nonlinear delay-differential equations, will be calibrated, analyzed, and simulated to assess the potential impact of the vaccination program in Burkina Faso, with emphasis on determining whether or not the vaccination program implemented as a sole intervention (or in combination with the traditional insecticide-based intervention) could lead to malaria elimination in Burkina Faso. Since bednet is the main insecticide-based intervention used in malaria-endemic areas, it is considered to be the traditional insecticide-based intervention (i.e., this study does not consider other insecticide-based vector control methods, such as larviciding and adulticiding, due to their reduced overall effectiveness) [14, 49, 50].

Some of the key novel features of the resulting age- and dose-structured malaria vaccination model include the parameterization of the model using observed epidemiological data for malaria dynamics in Burkina Faso [51] to fit the model, and use data from the Phase 3 clinical trial of the R21/Matrix-M vaccine conducted in Burkina Faso [33] to estimate the model parameters related to vaccine efficacy and coverage. The paper is organized as follows. The age- and dose-structured vaccination model is formulated in Section 2. The model is rigorously analyzed, for the existence and asymptotic stability of its disease-free equilibria, in Section 2.3. An expression for the vaccine-induced herd immunity threshold is also derived in this section, for two vaccination-dose scenarios (where only the first-dose is administered and where all four doses are administered). Global sensitivity analyses of the parameters, as well as numerical simulations of the model, are presented in Sections 4 and 6, respectively. Simulations for the full model are carried out and discussed in Section 6. The main findings of the study are summarized in Section 7.

## 2 Formulation of the Malaria Vaccination Model

In this section, an age- and dose-structured model will be formulated to assess the potential impact of vaccination (using the R21/Matrix-M vaccine) on the control of malaria in Burkina Faso, a malaria-endemic country in West Africa. The malaria vaccination program in Burkina Faso entails the vaccination of children of ages between five months and three years with the R21/Matrix-M vaccine. Specifically, they receive the first three doses of the R21/Matrix-M vaccine series (administered one month apart), followed by a booster (given one year after the third dose) [33]. Individuals who have received the first dose are eligible to receive the second dose; likewise, the third and fourth (booster) doses are administered sequentially to those who have completed the preceding two and three doses, respectively. In this section, a mathematical model for assessing the population-level impact of this vaccination program in Burkina Faso is designed, by stratifying the total population of children aged five months to five years at time *t*, denoted by *N*_*h*_(*t*), into the following two age groups:

### Group 1

This is the group of children 5 months to 3 years of age, who are eligible for routine antimalaria immunization in Burkina Faso [33].

### Group 2

This group consists of children in Burkina Faso who are between 3 and 5 years of age, and are not currently eligible for the routine vaccination. Although individuals in this group are not currently eligible for the routine malaria vaccination in Burkina Faso, we consider this group to assess the potential additional benefits of vaccinating individuals in this age group on the burden and possible elimination prospects of malaria in Burkina Faso. The addition of this hypothetical group is justified by the WHO recommendation that children under the age of five who are most vulnerable to malaria should be vaccinated (starting from the age of five months), and that the age range could be adjusted based on the level of malaria transmissibility in the malaria-endemic region being modeled [33, 37, 52].

Let *N*_*h*,1_(*t*) represent the total population of children in Group 1. This population is further subdivided into the mutually-exclusive compartments of susceptible (*S*_1_(*t*)), vaccinated with the first dose only (*V*_1,1_(*t*)), vaccinated with doses one and two only (*V*_2,1_(*t*)), vaccinated with all three doses (*V*_3,1_(*t*)), vaccinated with all three doses and the booster dose (*V*_*b*,1_(*t*)), exposed (*E*_1_(*t*)), infectious (*I*_1_(*t*)), and recovered (*R*_1_(*t*)) individuals, so that:

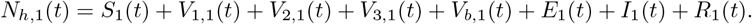

Similarly, let *N*_*h*,2_(*t*) represent the total population of children in Group 2, which is further subdivided into the mutually-exclusive compartments of susceptible (*S*_2_(*t*)), vaccinated with the first dose only (*V*_1,2_(*t*)), vaccinated with doses one and two only (*V*_2,2_(*t*)), vaccinated with all three doses (*V*_3,2_(*t*)), vaccinated with all three doses and the booster dose (*V*_*b*,2_(*t*)), exposed (*E*_2_(*t*)), infectious (*I*_2_(*t*)), and recovered (*R*_2_(*t*)) individuals. Thus,

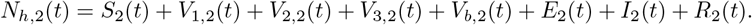

Hence, the total population of individuals in the two groups is given by:

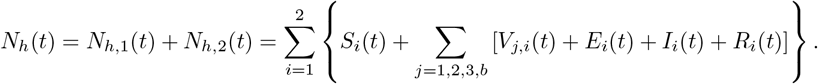

The total adult female *Anopheles* mosquito population (*Anopheles gambiae*, the most prevalent *Anopheles* species in Burkina Faso [1]) at time *t*, denoted by *N*_*m*_(*t*), is divided into the mutually-exclusive compartments of susceptible (*S*_*m*_(*t*)), exposed (*E*_*m*_(*t*)), and infectious (*I*_*m*_(*t*)) mosquitoes, so that

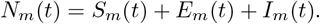

Let *λ*_*mh*_ represent the rate of transmission (or *force of infection*) of malaria from an infectious adult female *Anopheles* mosquito to a susceptible human, following an effective bite. Similarly, let *λ*_*hm*_ represent the rate of malaria transmission from an infectious human to a susceptible adult female *Anopheles* mosquito. It follows that:

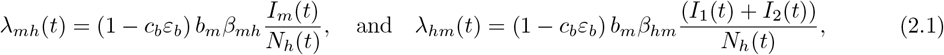

where 0 ≤ *c*_*b*_ < 1 and 0 < *ε*_*b*_ < 1 are the average coverage and efficacy of bednet usage, *b*_*m*_ is the average *per capita* biting rate of mosquitoes on humans, *β*_*mh*_ is the probability of malaria transmission from an infectious mosquito to a susceptible human, and *β*_*hm*_ is the probability of infection of susceptible mosquito following an effective bite on an infectious human (it should be mentioned that, in the formulation of the *forces of infection* in (2.1), the conservation law of mosquito bites (i.e., the total number of bites made by adult female mosquitoes equals the total number of bites received by the human host) was applied [53, 54]).

Based on the above definitions, derivations and assumptions, it follows that the age-structured and dose-structured model for assessing the impact of vaccination (using the R21/Matrix-M vaccine) in Burkina Faso is given by the following deterministic system of nonlinear delay-differential equations with fixed discrete delays, for all *t* ≥ 0 (where a dot represents differentiation with respect to time *t*):

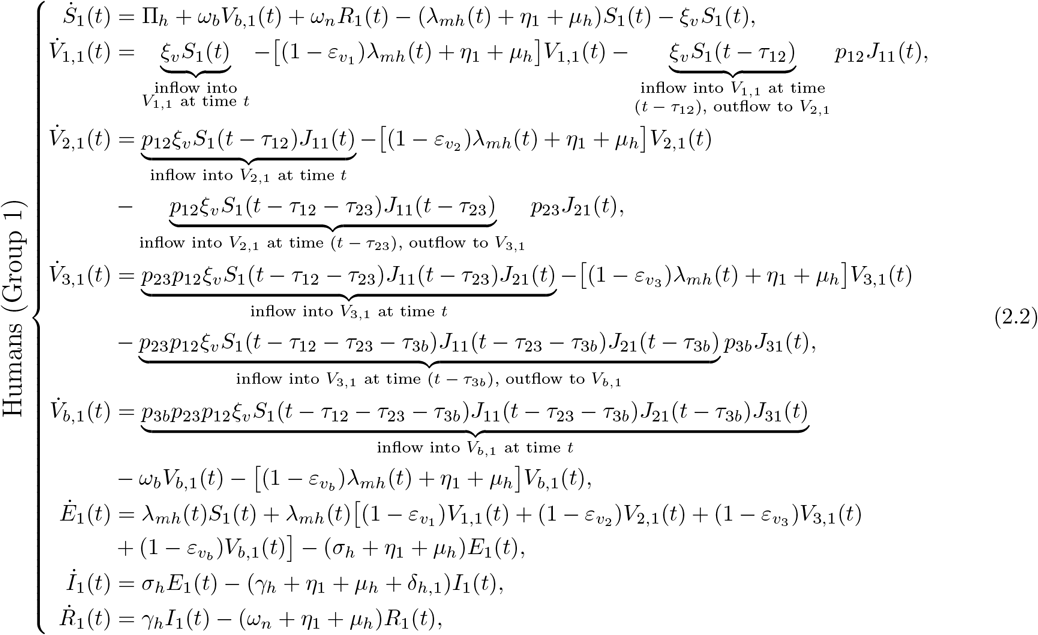

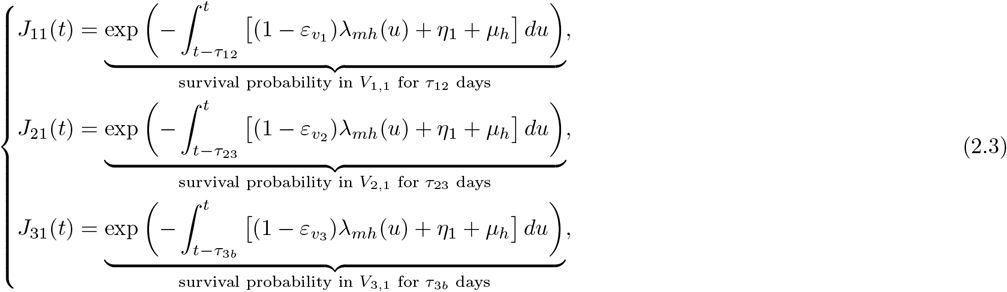

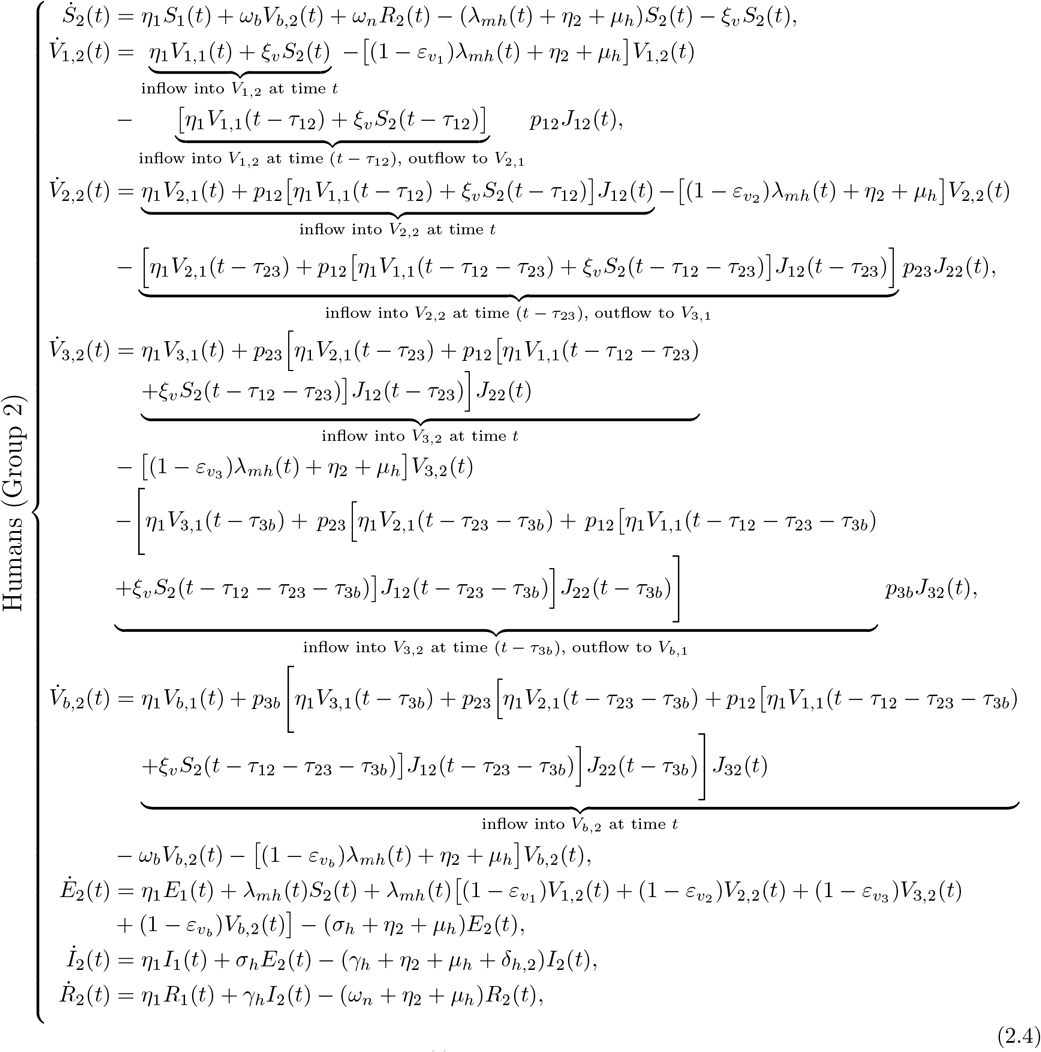

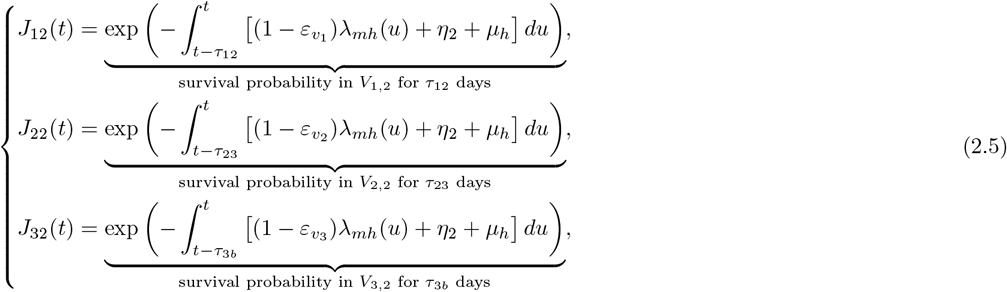

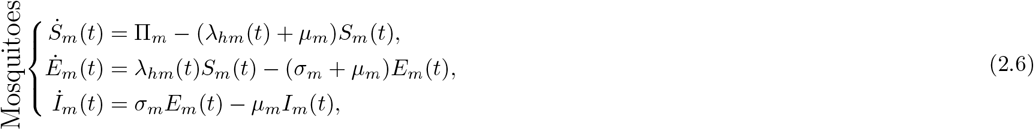

It is convenient to define *τ*:= *τ*_12_ + *τ*_23_ + *τ*_3*b*_, the maximum possible time-delay for an individual to receive all four doses of the malaria vaccine. Furthermore, let *θ* ∈ [−*τ*, 0] and

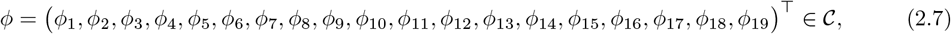

such that *ϕ*_1_(*θ*) = *ϕ*_1_(0) > 0, *ϕ*_2_(*θ*) = *ϕ*_2_(0) > 0, *ϕ*_17_(*θ*) = *ϕ*_17_(0) > 0, *ϕ*_*k*_(*θ*) = *ϕ*_*k*_(0) ≥ 0, for *k* = 3, …, 16, 18, 19, and 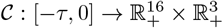 is the Banach space of continuous functions, equipped with the supremum norm:

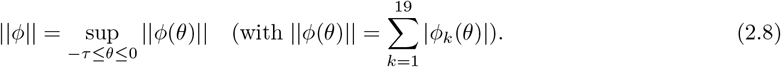

The initial data for the age- and dose-structured model 2.2–2.6 is then given by:

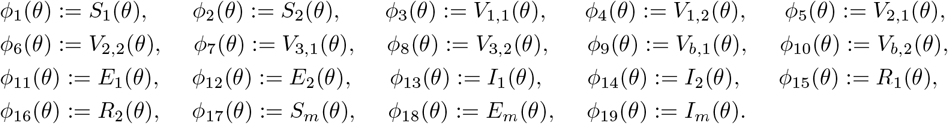

A flow diagram of the model is given in Figure 2, and the state variables and parameters of the model are described in Tables 1 and 2, respectively.

**Table 1.** Description of the state variables of the model (2.2)–(2.6), with *i* = 1, 2.

| State Variables | Description |
| --- | --- |
| $S_i$ | Number of unvaccinated susceptible individuals in Group $i$ |
| $V_{1,i}$ | Number of individuals in Group $i$ vaccinated with the first dose only |
| $V_{2,i}$ | Number of individuals in Group $i$ vaccinated with doses one and two only |
| $V_{3,i}$ | Number of individuals in Group $i$ vaccinated with all three doses |
| $V_{b,i}$ | Number of individuals in Group $i$ vaccinated with all three doses and the booster dose |
| $E_i$ | Number of exposed individuals in Group $i$ |
| $I_i$ | Number of infectious individuals in Group $i$ |
| $R_i$ | Number of recovered individuals in Group $i$ |
| $S_m$ | Number of susceptible mosquitoes |
| $E_m$ | Number of exposed mosquitoes |
| $I_m$ | Number of infectious mosquitoes |

**Table 2.**
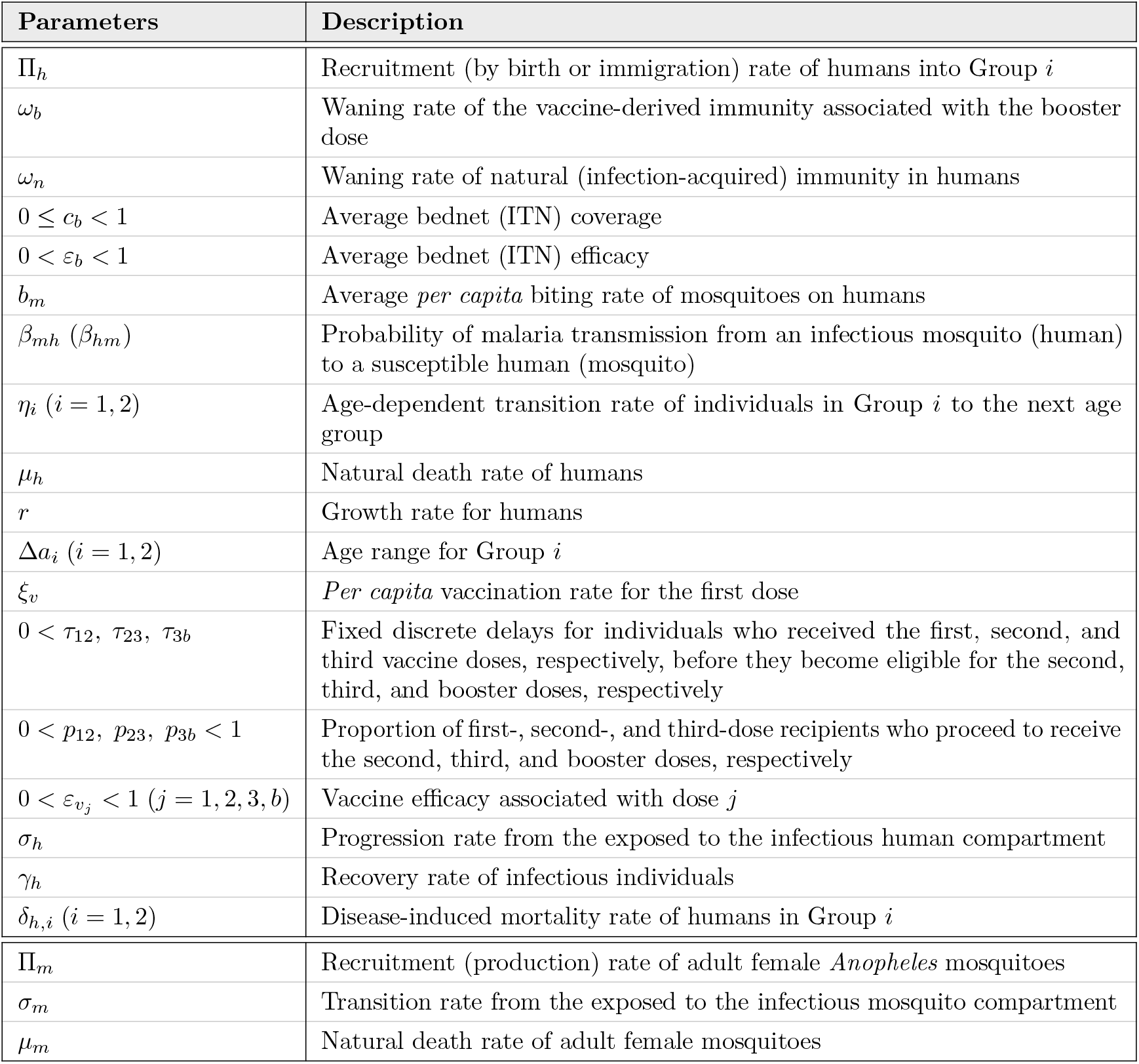
Description of the time-dependent and time-independent parameters of the model (2.2)–(2.6).

| Parameters | Description |
| --- | --- |
| $\Pi_h$ | Recruitment (by birth or immigration) rate of humans into Group $i$ |
| $\omega_b$ | Waning rate of the vaccine-derived immunity associated with the booster dose |
| $\omega_n$ | Waning rate of natural (infection-acquired) immunity in humans |
| $0 \leq c_b < 1$ | Average bednet (ITN) coverage |
| $0 < \varepsilon_b < 1$ | Average bednet (ITN) efficacy |
| $b_m$ | Average *per capita* biting rate of mosquitoes on humans |
| $\beta_{mh}$ ($\beta_{hm}$) | Probability of malaria transmission from an infectious mosquito (human) to a susceptible human (mosquito) |
| $\eta_i$ ($i = 1, 2$) | Age-dependent transition rate of individuals in Group $i$ to the next age group |
| $\mu_h$ | Natural death rate of humans |
| $r$ | Growth rate for humans |
| $\Delta a_i$ ($i = 1, 2$) | Age range for Group $i$ |
| $\xi_v$ | *Per capita* vaccination rate for the first dose |
| $0 < \tau_{12}, \tau_{23}, \tau_{3b}$ | Fixed discrete delays for individuals who received the first, second, and third vaccine doses, respectively, before they become eligible for the second, third, and booster doses, respectively |
| $0 < p_{12}, p_{23}, p_{3b} < 1$ | Proportion of first-, second-, and third-dose recipients who proceed to receive the second, third, and booster doses, respectively |
| $0 < \varepsilon_{v_j} < 1$ ($j = 1, 2, 3, b$) | Vaccine efficacy associated with dose $j$ |
| $\sigma_h$ | Progression rate from the exposed to the infectious human compartment |
| $\gamma_h$ | Recovery rate of infectious individuals |
| $\delta_{h,i}$ ($i = 1, 2$) | Disease-induced mortality rate of humans in Group $i$ |
| $\Pi_m$ | Recruitment (production) rate of adult female *Anopheles* mosquitoes |
| $\sigma_m$ | Transition rate from the exposed to the infectious mosquito compartment |
| $\mu_m$ | Natural death rate of adult female mosquitoes |

**Figure 2.**
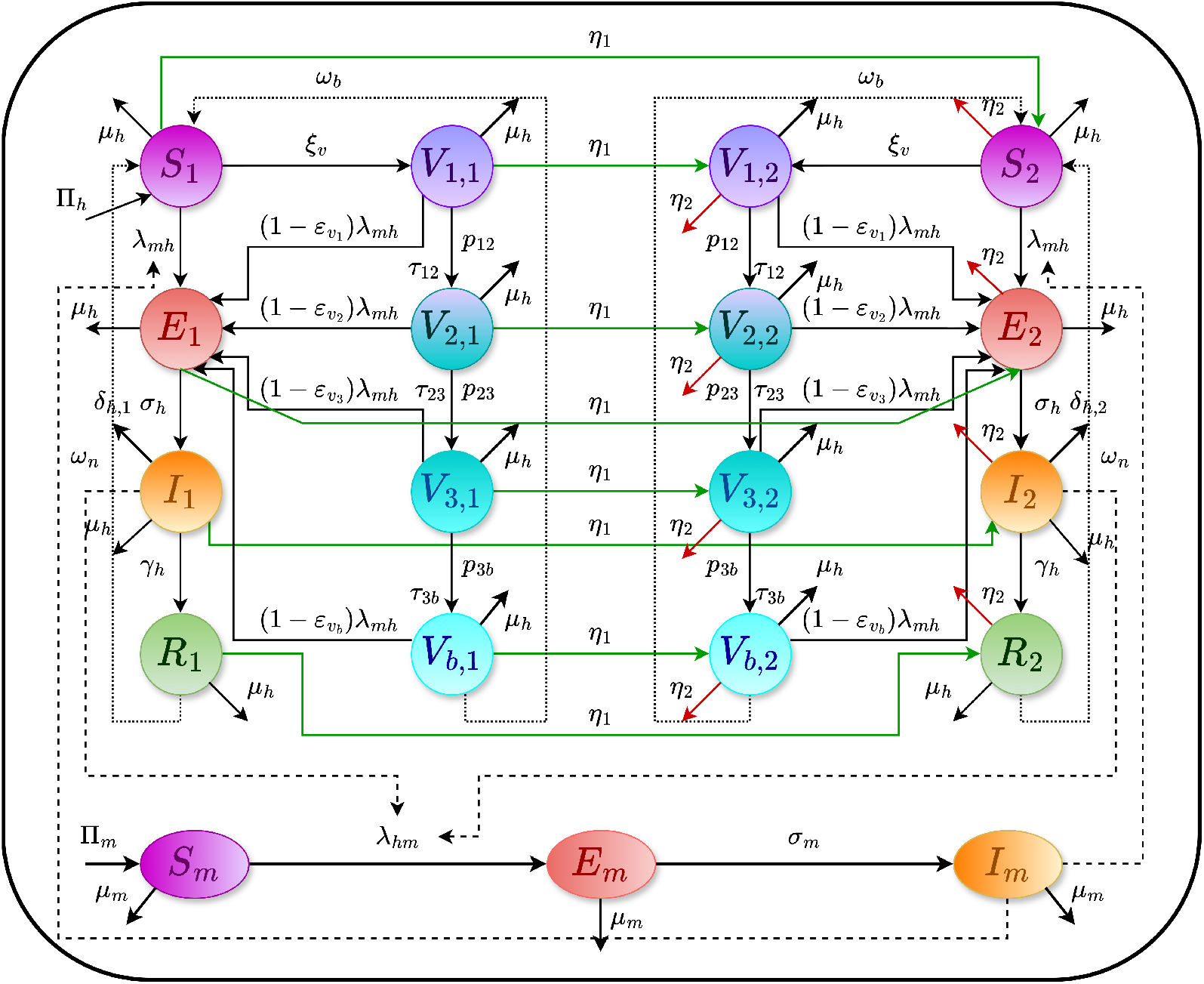
Flow diagram of the age- and dose-structured malaria vaccination model (2.2)–(2.6).

In the model (2.2)–(2.6), Π_*h*_ is the recruitment rate (by birth or immigration) of individuals (it is assumed that all recruited individuals are susceptible and enter into Group 1 only), *ω*_*b*_ is the waning rate of the vaccine-derived immunity associated with the booster dose, *ω*_*n*_ is the waning rate of natural (infection-acquired) immunity, *λ*_*mh*_ and *λ*_*hm*_ are the *forces of infection* defined in (2.1), *ξ*_*v*_ is the *per capita* rate of administration of the first dose, *η*_*i*_ (with *i* = 1, 2) is the age-dependent transition rate out of individuals in age-group *i* due to death or maturation to the next age group, and *µ*_*h*_ is the natural death rate of humans (assumed to be the same in all human epidemiological compartments). The transition parameter *η*_*i*_ is defined as [55]:

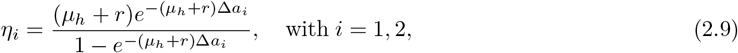

where *r* is the constant growth rate of the human population (assuming exponential equilibrium growth), and Δ*a*_*i*_ is the age range of Group *i*, with *i* = 1, 2 (Δ*a*_*i*_ = 3 for Group 1 and Δ*a*_*i*_ = 2 for Group 2). The term 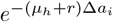 accounts for the probability of staying in the current age group (i.e., the individual did not die or mature to the next age group). Hence, the term 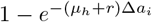 represents the total probability of an individual leaving the current age group due to either death (at an effective rate, 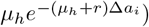 or aging to the next age group (at an effective rate, 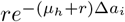). Hence, *η*_*i*_ is the ratio of the sum of the effective death and maturation rates for individuals in group *i* (given by 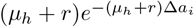) to the overall probability of an individual leaving the age group *i*, due to death or maturation (given by, 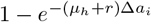).

Furthermore, 0 < *τ*_12_, *τ*_23_, *τ*_3*b*_ are fixed discrete delays (in days) for individuals who received the first, second, and third vaccine doses, respectively, before they become eligible for the second, third, and booster doses, respectively. Similarly, 0 < *p*_12_, *p*_23_, *p*_3*b*_ < 1 denote the proportions of individuals who, after receiving the first, second, and third vaccine doses, respectively, proceed to receive the second, third, and the booster doses, respectively. The parameter 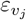 (with *j* = 1, 2, 3 and *b*) represents the vaccine efficacy associated with dose *j, σ*_*h*_ is the progression rate of individuals from the exposed class (*E*_*i*_) to the infectious class (*I*_*i*_), with *i* = 1, 2, *γ*_*h*_ is the recovery rate of infectious individuals (assumed to be the same for each of the two groups), and *δ*_*h,I*_ (with *i* = 1, 2) is the disease-induced mortality of humans in group *i* (it is assumed that *δ*_*h*,1_ > *δ*_*h*,2_). Finally, Π_*m*_ is the recruitment rate of adult female *Anopheles* mosquitoes into the mosquito population (all assumed to be susceptible), *σ*_*m*_ is the transition rate of exposed mosquitoes (in the *E*_*m*_ class) to the infectious class (*I*_*m*_) and *µ*_*m*_ is the natural death rate of adult female mosquitoes.

The main assumptions made in the formulation of the model (2.2)–(2.6) include:

a. We assume a large and well-mixed total human and vector populations, with exponentially-distributed waiting time in each epidemiological (and vector ecological) compartment.
b. Recruitment into Group 2 occurs only by maturation, at the rate *η*_1_ given by (2.9) (i.e., there is no recruitment due to immigration into Group 2).
c. No vertical or congenital (mother-to-child) transmission of malaria is assumed (congenital malaria is rare, with global prevalence estimated to be about 6.9% [56, 57]).
d. The R21/Matrix-M vaccine is imperfect, with protective efficacy that wanes over time, at the rate *ω*_*b*_ [47, 58] *(since the first three doses are administered only a month apart, it is assumed that the vaccine does not wane for individuals who received these doses; the vaccine is assumed to wane only at the booster stage). Furthermore, infection-acquired (natural) immunity wanes over time, at the rate ω*_*n*_ [59–61].
e. Exposed (i.e., newly infected) individuals are assumed to be incapable of transmitting malaria to susceptible mosquitoes. This is a simplifying assumption needed for mathematical tractability.
f. No reinfection of recovered individuals is assumed (this is also a simplifying assumption for mathematical tractability). Furthermore, it is assumed that asymptomatic transmission of malaria does not occur (although recent evidence show that this could be significant for malaria transmission dynamics [62, 63]).

The model (2.2)–(2.6) is an extension of many prior malaria models in the literature that incorporate age-structure and/or vaccination, such as the models in [46, 64–66], by, *inter alia*:

i. Incorporating dose-structure (with three primary vaccine doses, *V*_1,*i*_, *V*_2,*i*_, and *V*_3,*i*_, followed by a booster dose, *V*_*b,i*_; with *i* = 1, 2) of the recently approved malaria vaccine R21/Matrix-M. This extends the age-structured malaria model presented by Forouzannia and Gumel [64] (which did not include vaccination). It also extends the malaria vaccination model presented by Naandam et al. [46] (which did not have dose-structure or age-structure). It is also a slight extension of the age-structured and dose-structured malaria vaccination models presented in [65, 66] (their models lumped the first three doses into one epidemiological compartment, followed by another compartment for the booster dose; as against using three compartments for the primary doses and another for the booster in our study).
ii. A key feature of this study is the incorporation of time delays associated with the administration of successive vaccine doses. In particular, we introduce delay parameters *τ*_12_ (28 days), *τ*_23_ (28 days), and *τ*_3*b*_ (365 days), representing the time intervals between receiving the first and second doses, the second and third doses, and the third and booster doses, respectively. These delays capture the realistic vaccination schedule of administering three doses four weeks apart, followed by a booster dose given one year after the third dose. This extension generalizes the framework in [65, 66], where such inter-dose delays were not considered.
iii. Bednet-based intervention is not considered in the malaria vaccination or age-structured models in [46, 64, 65].
iv. Adding an ecological compartment for exposed (i.e., newly-infected but not infectious) mosquitoes (*E*_*m*_). This was not considered in the models presented in [64, 65].

### 2.1 Baseline values of the parameters of the model

In this section, the derivation of the numerical values of each of the parameters of the model (2.2)–(2.6) will be discussed in the context of malaria transmission dynamics in Burkina Faso.

#### 2.1.1 Human parameters

The average life expectancy of humans in Burkina Faso is estimated to be about 59.2 years [67, 68] (for the period 2012 to 2021). Hence, *µ*_*h*_ = 1*/*(59.2 × 365) ≈ 0.0000463 *per* day. Furthermore, using demographic data for Burkina Faso from the World Bank for the period 2012 to 2021 [69], it can be shown that the annual growth rate for Burkina Faso is *r* = 0.0000762 *per* day. Similarly, following Hethcote [55], the transition rates *η*_*i*_ (for *i* = 1, 2) are calculated from (2.9) using the formula:

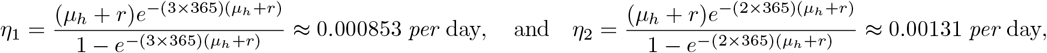

respectively. Data from the Institute for Health Metrics and Evaluation [70] show that the total population of children under the age of 5 in Burkina Faso (calculated for the period between 2012 to 2021), is estimated to be 3, 725, 843. It is assumed that children in Group 1 (i.e., of age 0 to 3 years) constitute 61% of this number. Hence, *N*_*h*,1_(*t*) = 2, 272, 764. Thus, at the disease-free equilibrium, Π_*h*_*/*(*η*_1_ + *µ*_*h*_) = 2, 272, 764, so that Π_*h*_ = 2, 044 *per* day. Since the R21/Matrix-M vaccine is only being rolled out now in the 20 countries in sub-Saharan Africa [36, 37], no real data exist for estimating the dose-structured vaccination rate. Consequently, data for RTS,S/AS01 vaccination coverage (note that the RTS,S/AS01 vaccine has been deployed in sub-Saharan Africa since 2023 [35]) are available for the first dose. It was estimated that 6,557 children in Burkina Faso received the first dose of RTS,S/AS01 during February of 2024 [71] (representing 36% of the monthly target in Burkina Faso, with a total target population of 248,986 children). Hence, we estimate the daily rate of administration of the first dose of the R21/Matrix-M vaccine in Burkina Faso (assuming a 30-day month) to be approximately 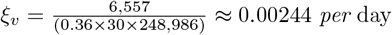. There is currently no data on the waning rate of the booster dose of the R21/Matrix-M vaccine (as the clinical trial to determine this is currently ongoing [33]), we estimate the waning rate by comparing it with that of the RTS,S/AS01 vaccine (which wanes within 2.5 to 4 years after the administration of the booster dose, depending on the malaria transmission intensity-moderate or high-in the community [58]). Assuming the upper bound of four years for the booster dose of the RTS,S/AS01 vaccine to wane, and using the 67% efficacy of the booster dose of the R21/Matrix-M vaccine [33], the waning of the R21/Matrix-M booster dose is assumed to obey the exponential function *W* (*t*) = *a* exp(−*b*(*t* −365)), where *a* = 0.67 is the efficacy of the booster dose and *b* = 0.003154 is the waning rate corresponding to the assumed 4-year duration for the R21/Matrix-M vaccine to wane starting 1-year after booster administration (Figure 3 shows the profile of the exponential function *W* (*t*) used to derive the waning rate of the R21/Matrix-M booster dose). Hence, based on this analysis, the waning rate of the R21/Matrix-M booster vaccine is set at *ω*_*b*_ = 0.003154 *per* day.

**Figure 3.**
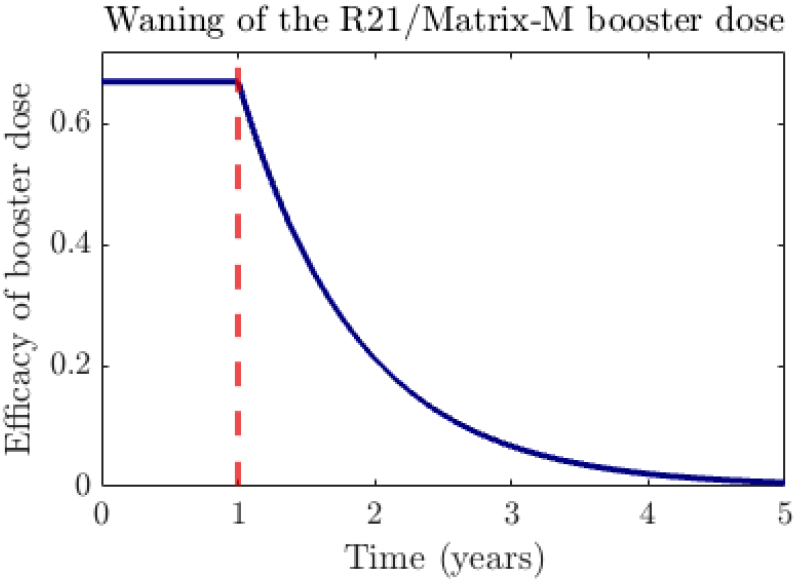
Profile of the exponential function, *W* (*t*) = 0.67exp(−0.003154(*t* − 365)) (blue curve) used to estimate the waning rate of the R21/Matrix-M vaccine, assuming waning begins one year post-administration (dashed vertical red line) and reaches near zero within four years.

The waning rate of natural immunity in humans (under 5 years of age) is reported in [61, 72] as *ω*_*n*_ = 0.0027 *per* day. The value of *c*_*b*_ is estimated by taking the average of the reported bednet coverages for children under five in Burkina Faso during 2014 (estimated to be 75.3%), 2018 (54.4%) and 2021 (67.4%) [73]. That is, we set *c*_*b*_ = (0.753 + 0.544 + 0.674)*/*3 = 0.657. In line with [74], we set *ε*_*b*_ = 0.85. The probability of malaria transmission (*β*_*mh*_) *per* bite from an infectious female mosquito to a susceptible human is assumed to be the same across the two age groups, and is set as *β*_*mh*_ = 0.181 (which falls within the range (0.072 − 0.64) reported in [75], and is also consistent with the estimated value in [26]).

As the primary three doses are administered four weeks apart, and the booster dose is administered one year after the third dose, the fixed discrete delays, *τ*_12_, *τ*_23_, and *τ*_3*b*_, are set at *τ*_12_ = *τ*_23_ = 28 days, and *τ*_3*b*_ = 365 days [33]. The proportions *p*_12_, *p*_23_, and *p*_3*b*_, of individuals who received doses 2,3, and the booster dose, are currently unknown, but are set at the optimistic value of 50% for these doses. That is, *p*_12_ = *p*_23_ = *p*_3*b*_ = 0.5. Results from clinical trials in Burkina Faso conducted in children 5 to 36 months of age (i.e., Group 1 in our study) [33] show that the efficacy of the R21/Matrix-M vaccine (against all clinical malaria episodes) is 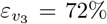 days after the third dose, and reduces to 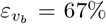 after the booster dose [33]. These efficacy figures are assumed to be the same for the 0–60 months age group (Groups 1 and 2). While specific vaccine efficacy for the first and second doses of the R21/Matrix-M vaccine are not currently available, several controlled and clinical trials have shown a strong positive correlation between vaccine-induced anti-NANP (Asn–Ala–Asn–Pro) IgG (immunoglobulin G) antibody levels and vaccine efficacy [33, 47, 48]. For instance, Datoo et al. [47] showed that anti-NANP IgG levels of 610 EU/ml (95% CI), 6, 352 EU/ml (95% CI), 11, 438 EU/ml (95% CI), and 10, 156 EU/ml (95% CI) were observed 28 days after doses 1, 2, 3, and booster dose, respectively, in the age group 5-17 months. Notably, participants had no detectable baseline anti-NANP IgG levels before receiving Dose 1 [47]. Based on these antibody levels, and since higher antibody titers correspond to higher vaccine efficacy, we assume a proportional relationship between vaccine efficacy and log(titer). This assumption is justified because immune responses often follow a logarithmic or saturating behavior [76]. Thus, assuming that antibody levels remain the same for the extended age group (0–60 months), the vaccine efficacy for first and second doses of the R21/Matrix-M vaccine are estimated as: 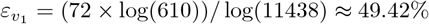, and 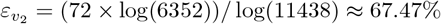, respectively. The progression rate from the exposed class to the infectious class for individuals in Group *i* (*i* = 1, 2) is assumed to be the same, and set at *σ*_*h*_ = 0.083 *per* day [75]. Similarly, the recovery rates for individuals in each group is set at *γ*_*h*_ = 0.0025 *per* day [75].

#### 2.1.2 Mosquito parameters

The average total human population in Burkina Faso is estimated from Worldometer [77] to be approximately 19,597,808 (calculated for the period 2012 to 2021). Considering the vector-host ratio (total adult female mosquitoes to total humans) in Burkina Faso to be 2: 1 [61], the average total number of adult female mosquitoes is set at *N*_*m*_ = (2 × 19, 597, 808) = 39, 195, 616. Thus, at the disease-free equilibrium, Π_*m*_ = *N*_*m*_ × *µ*_*m*_ = 2, 061, 689 *per* day. The probability of infection (*β*_*hm*_) from infectious individuals to susceptible female *Anopheles* mosquitoes is set to *β*_*hm*_ = 0.012, which is within the range used in [61] and is slightly below the value used for adult humans in [75, 78]. It is estimated that, after biting an infectious human, newly-infected female *Anopheles* mosquitoes take about 7 to 10 days (with a mean of 8.5 days) to progress to the infectious class [79]. Hence, the parameter (*σ*_*m*_) is set at 1*/*8.5 *per* day. Addawe and Lope [61] estimated that *Anopheles gambiae* has an average lifespan of 23 days (23 *±* 8.29 days). Hence, the natural death rate (*µ*_*m*_) for female *Anopheles* mosquitoes is set at *µ*_*m*_ = 1*/*19 ≈ 0.0526 *per* day. The values of the remaining three parameters of the model (namely *δ*_*h*,1_, *δ*_*h*,2_, and *b*_*m*_) are unknown and will be estimated by fitting the vaccination-free model with malaria mortality data in Subsection 2.2. The values of all the known parameters described above are tabulated in Table 3.

**Table 3.** Baseline values and ranges of the known parameters of the model (2.2)–(2.6).

| Parameter | Baseline value | Range | Source |
| --- | --- | --- | --- |
| $\Pi_h$ | 2,044 day<sup>-1</sup> | (1,635 – 2,453) day<sup>-1</sup> | [80] |
| $\omega_b$ | 0.003154 day<sup>-1</sup> | (0.002523 – 0.003785) day<sup>-1</sup> | [47, 58] |
| $\omega_n$ | 0.0027 day<sup>-1</sup> | (0.000055 – 0.011) day<sup>-1</sup> | [61, 72] |
| $c_b$ | 0.657 (dimensionless) | (0.5256 – 0.7884) (dimensionless) | [73] |
| $\varepsilon_b$ | 0.85 (dimensionless) | (0.68 – 1.00) (dimensionless) | [74] |
| $\beta_{mh}$ | 0.181 (dimensionless) | (0.072 – 0.64) (dimensionless) | [26, 75] |
| $\eta_1$ ($\eta_2$) | 0.000853 (0.00131) day<sup>-1</sup> | (0.000682 – 0.001024) [(0.001048 – 0.001572)] day<sup>-1</sup> | [55] |
| $\mu_h$ | 0.0000463 day<sup>-1</sup> | (0.000037 – 0.000056) day<sup>-1</sup> | [67, 68] |
| $r$ | 0.0000762 day<sup>-1</sup> | (0.000061 – 0.000091) day<sup>-1</sup> | [69] |
| $\xi_v$ | 0.00244 day<sup>-1</sup> | (0.00195 – 0.00293) day<sup>-1</sup> | [33, 71] |
| $\tau_{12}, \tau_{23}$ | 28 days | (22 – 34) days | [33] |
| $\tau_{3b}$ | 365 days | (292 – 438) days | [33] |
| $p_{12}, p_{23}, p_{3b}$ | 0.5 (dimensionless) | (0.4 – 0.6) (dimensionless) | Assumed |
| $\varepsilon_{v_1}$ ($\varepsilon_{v_2}$) | 49.42 (67.47) % | (39.54 – 59.30) [(53.98 – 80.96)] % | [33] |
| $\varepsilon_{v_3}$ ($\varepsilon_{v_b}$) | 72 (67) % | (57.6 – 86.4) [(53.6 – 80.4)] % | [33] |
| $\sigma_h$ | 0.083 day<sup>-1</sup> | (0.067 – 0.2) day<sup>-1</sup> | [72, 75] |
| $\gamma_h$ | 0.0025 day<sup>-1</sup> | (0.0014 – 0.017) day<sup>-1</sup> | [75] |
| $\Pi_m$ | 2,061,689 day<sup>-1</sup> | (1,649,351 – 2,474,027) day<sup>-1</sup> | [81–83] |
| $\beta_{hm}$ | 0.012 (dimensionless) | (0.01–0.27) (dimensionless) | [61, 75, 78] |
| $\sigma_m$ | 0.1 day<sup>-1</sup> | (0.029 – 0.33) day<sup>-1</sup> | [72, 79, 84] |
| $\mu_m$ | 0.0526 day<sup>-1</sup> | (0.032 – 0.068) day<sup>-1</sup> | [61] |

### 2.2 Data fitting and parameter estimation

In this subsection, the model (2.2)–(2.6) without vaccination, given by equation (D.1) of Appendix D, is fitted with the observed yearly cumulative malaria mortality data for children under 5 years of age in Burkina Faso (over the period 1980–2021 [51]), to estimate the three unknown parameters of the model (namely, *δ*_*h*,1_, *δ*_*h*,2_, and *b*_*m*_). Specifically, the model fitting is performed using a nonlinear least-squares optimization implemented *via* MATLAB’s *lsqcurvefit* function, which minimizes the sum of squared differences between the observed yearly cumulative mortality and the corresponding simulated values obtained from the vaccination-free model (D.1). The model is fitted using a segment of the data from 1980 to 2015, and the remaining data (from 2016 to 2021) is used to cross-validate the model. The results obtained, depicted Figure 4, show a very good fit and cross-validation to the cumulative mortality data. MATLAB’s built-in bootstrapping (*bootstrp*) and confidence intervals (*prctile*) routines were used for the parameter estimation and computation of the 95% confidence intervals for the estimated parameters obtained from the fitting. In particular, 10, 000 bootstrap replicates of the observed cumulative mortality data (resampled with replacement) and their corresponding fitted parameter estimates are generated to construct the bootstrap distribution for parameter estimation. The estimated values of the three unknown parameters obtained from the fitting, along with their respective 95% credible intervals are summarized in Table 4. These estimated values and ranges are reasonable, and are consistent with those obtained in several malaria modeling studies, such as [72, 75, 85].

**Table 4.**
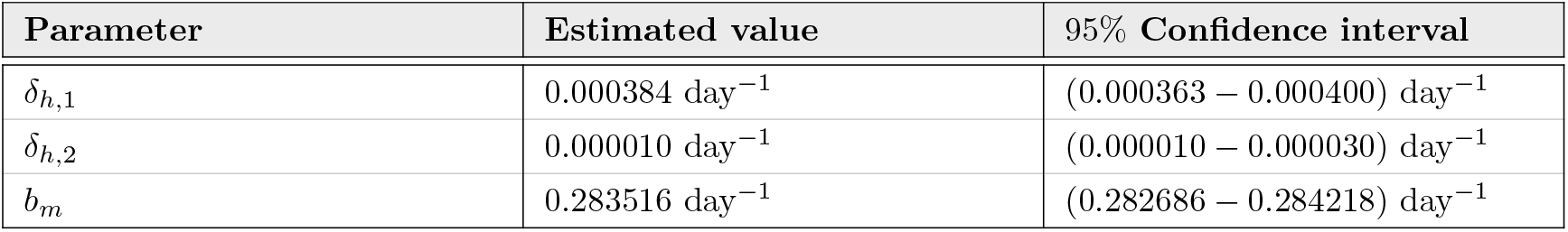
Values of the unknown parameters of the model (2.2)–(2.6).

**Figure 4.**
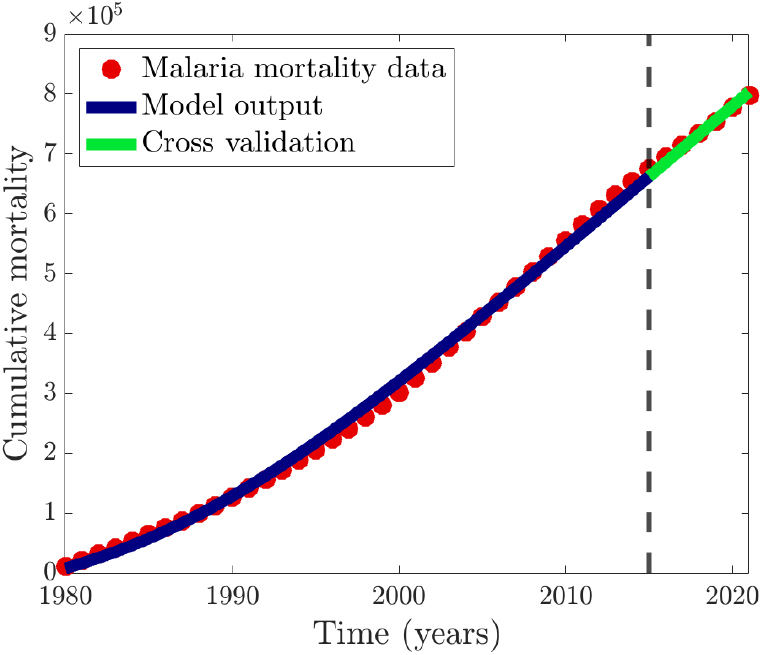
Data fitting and cross-validation of the vaccination-free version of the model (2.2)–(2.6), given by equation (D.1) of Appendix D, to estimate the three unknown parameters (*δ*_*h*,1_, *δ*_*h*,2_ and *b*_*m*_) using yearly cumulative malaria mortality data for children under 5 years of age in Burkina Faso for the period from 1980 to 2021 [51]. The fitting is carried out using least-squares regression and the baseline values of the known parameters of the model tabulated in Table 3. The data fitting (blue curve) was carried out using the yearly cumulative malaria mortality data (red dots) from 1980 to 2015 (to the left of the dashed vertical black line), and the cross-validation (green curve) was carried out using the yearly cumulative malaria mortality data (red dots) for the remaining six years, from 2016 to 2021 (to the right of the dashed vertical black line) in Burkina Faso. The estimated baseline values of the three unknown parameters of model (2.2)–(2.6), with their 95% confidence intervals, are given in Table 4.

### 2.3 Basic qualitative properties of the model

In this section, the basic qualitative properties of the model (2.2)–(2.6) will be assessed to establish its well-posedness with respect to the existence, uniqueness and boundedness of its solutions. Since the model monitors the temporal dynamics of human and mosquito populations, all its parameters and state variables are assumed to be non-negative for all time *t* ≥ 0 (with *S*_*i*_(0) (for *i* = 1, 2) and *S*_*m*_(0) taken to be strictly positive, to ensure the presence of humans and mosquitoes in the population). It is convenient to let:

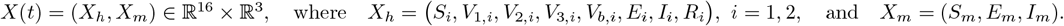

represent the vector of the state variables of the model (2.2)–(2.6). Recall from (2.7) that

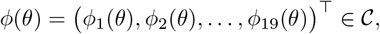

and consider the positive cone 𝒞_+_ ⊆ 𝒞, defined by:

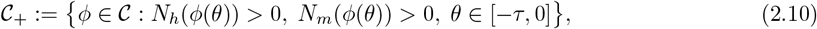

so that all the terms in the expressions for the force of infection, given by (2.1), are well-defined on 𝒞_+_. Let

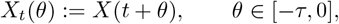

be the history segment of the solution of the model from time *t* − *τ* to *t*. Using this notation, the model (2.2)–(2.6) can then be rewritten as

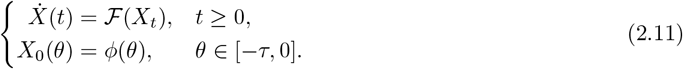

where the vector-valued function 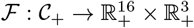 is defined component-wise by the right-hand sides of the system (2.2)–(2.6), and the initial history *ϕ* is given by (2.7). We claim the following result.

#### Theorem 2.1

(Existence and uniqueness of solutions). *Suppose that all parameters of the model* (2.2)*–*(2.6) *are non-negative. Furthermore, let τ* > 0 *(as defined in Section 2) denote the maximum possible time-delay in the model, and let* 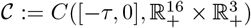 *denote the Banach space of continuous functions equipped with the supremum norm*

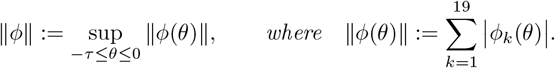

*Suppose that the initial history function ϕ, as given in* (2.7), *lies in the biologically-feasible subset* 𝒞_+_ ⊆ 𝒞 *(defined in* (2.10)*). Then, there exists any δ* > 0 *such that the initial-value problem* (2.11) *has a unique solution of the form* 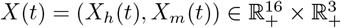, *with t* ∈ [−*τ, δ*) *and X*_0_(*θ*) = *ϕ*(*θ*) *(i*.*e*., *X*(*t* + *θ*) = *ϕ*(*θ*) *for θ* ∈ [−*τ*, 0]).

The proof of the Theorem 2.1 is given in the Appendix A. Consider, next, the following biologically-feasible region for the model (2.2)–(2.6) (for *t* ≥ 0):

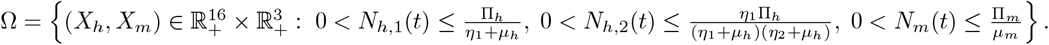

#### Theorem 2.2.

*Consider the model* (2.2)*–*(2.6) *with S*_1_(0) > 0, *S*_2_(0) > 0 *and S*_*m*_(0) > 0. *Then, all solutions of the model are bounded for all time t* ≥ 0. *Furthermore, the region* Ω *is positively-invariant with respect to the flow generated by the model* (2.2)*–*(2.6).

*Proof*. By adding the equations in the model (2.2)–(2.6), it can be shown that the equations for the rate of change of the total populations of humans in Group 1, humans in Group 2, and mosquitoes are given, respectively, by:

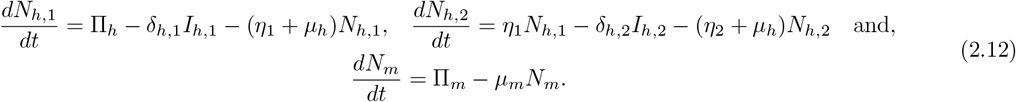

so that (by the non-negativity of the state variables and parameters of the model (2.2)–(2.6)),

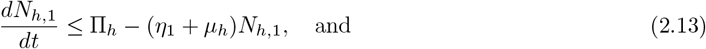

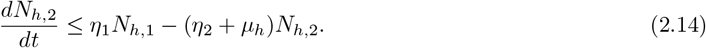

It follows from (2.13)(b y using a standard comparison theorem [86]), that 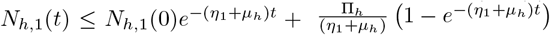. Hence, if 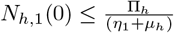, then 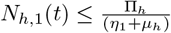 for all *t* ≥ 0. Using this bound in (2.14) gives 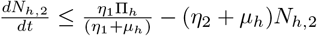. Thus, by a standard comparison theorem [86],

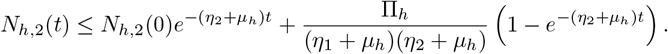

Similarly, it follows from (2.12) that 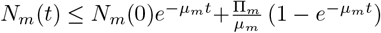. Hence, if 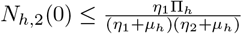 and 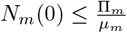, then 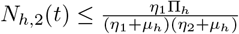, and 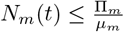, respectively, for all *t* ≥ 0. These results imply that the total populations of humans in Group 1, humans in Group 2, and mosquitoes, *N*_*h*,1_(*t*), *N*_*h*,2_(*t*) and *N*_*m*_(*t*), are bounded above by 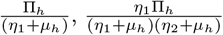 and 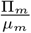, respectively, for all *t* ≥ 0. Since, *S*_*i*_(*t*) > 0for *i* = 1, 2, and *S*_*m*_(*t*) > 0, it follows that these total populations are bounded below by zero. Consequently, 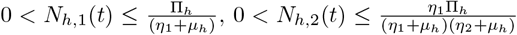, and 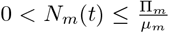 hold for all *t* ≥ 0. Hence, the total populations of humans in Group 1, humans in Group 2, and mosquitoes, *N*_*h*,1_(*t*), *N*_*h*,2_(*t*) and *N*_*m*_(*t)* are bounded in Ω (and so all the state variables of the model (2.2)–(2.6) are bounded in Ω). Thus, every solution of the model with initial condition in Ω remains in Ω for all *t* ≥ 0. Hence, the bounded region Ω is positively-invariant with respect to the flow generated by the model (2.2)–(2.6).

As a consequence of the above theorem, it is sufficient to study the dynamics of the model (2.2)–(2.6) in the region Ω, where the model well-posed mathematically and epidemiologically [87].

## 3 Asymptotic Stability of Disease-free Equilibrium of the Model

The model (2.2)–(2.6) has a unique disease-free equilibrium (DFE), denoted by ℰ_0_, given by:

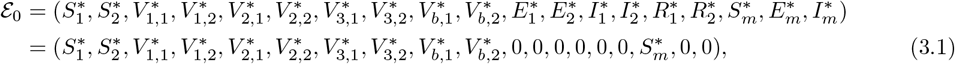

where,

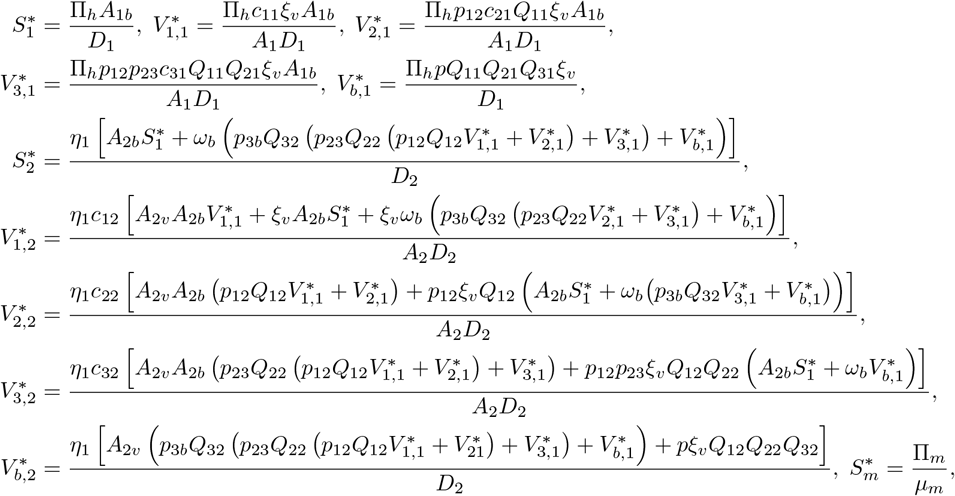

with *A*_1_ = *η*_1_ + *µ*_*h*_, *A*_2_ = *η*_2_ + *µ*_*h*_, *A*_1*v*_ = *A*_1_ + *ξ*_*v*_, *A*_1*b*_ = *A*_1_ + *ω*_*b*_, *A*_2*v*_ = *A*_2_ + *ξ*_*v*_, *A*_2*b*_ = *A*_2_ + *ω*_*b*_, *p* = *p*_12_*p*_23_*p*_3*b*_, *Q*_11_ = exp (−*A*_1_*τ*_12_), *Q*_21_ = exp (−*A*_1_*τ*_23_), *Q*_31_ = exp (−*A*_1_*τ*_3*b*_), *Q*_12_ = exp (−*A*_2_*τ*_12_), *Q*_22_ = exp (−*A*_2_*τ*_23_), *Q*_32_ = exp (−*A*_2_*τ*_3*b*_), 0 < *c*_11_ = 1 − *p*_12_*Q*_11_ < 1, 0 < *c*_21_ = 1 − *p*_23_*Q*_21_ < 1, 0 < *c*_31_ = 1 − *p*_3*b*_*Q*_31_ < 1, 0 < *c*_12_ = 1 − *p*_12_*Q*_12_ < 1, 0 < *c*_22_ = 1 − *p*_23_*Q*_22_ < 1, 0 < *c*_32_ = 1 − *p*_3*b*_*Q*_32_ < 1, *D*_1_ = *A*_1*v*_*A*_1*b*_ − *pξ*_*v*_*ω*_*b*_*Q*_11_*Q*_21_*Q*_31_, and *D*_2_ = *A*_2*v*_*A*_2*b*_ − *pξ*_*v*_*ω*_*b*_*Q*_12_*Q*_22_*Q*_32_ (here *D*_1_, *D*_2_ > 0; see Appendix B).

The local asymptotic stability of the disease-free equilibrium of the model (2.2)–(2.6) will now be analyzed.

### 3.1 Local asymptotic stability of disease-free equilibrium in model (2.2)**–**(2.6)

The local asymptotic stability of the DFE, ℰ_0_, of the model (2.2)–(2.6) will be explored using the next-generation operator method [88, 89]. Specifically, using the notation in [88], the matrices *F* and *V*, for the new infection terms and the linear transitions between the infected compartments of the model, are given, respectively, by:

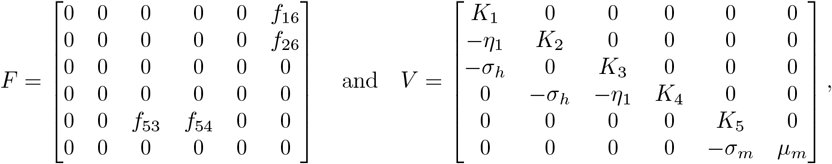

where, 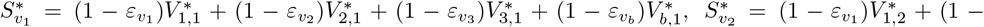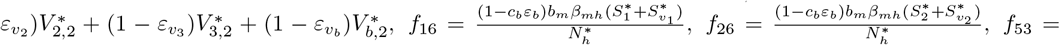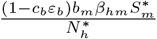, and 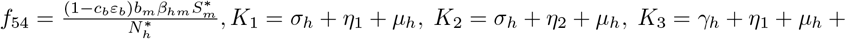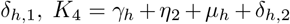, and *K*_5_ = *σ*_*m*_ + *µ*_*m*_. It is convenient to define the following threshold quantity (where *ρ* denotes the spectral radius):

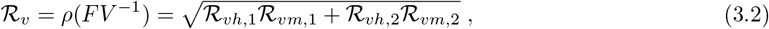

with ℛ_*vh*,1_, ℛ_*vh*,2_, ℛ_*vm*,1_, and ℛ_*vm*,2_ are defined as follows:

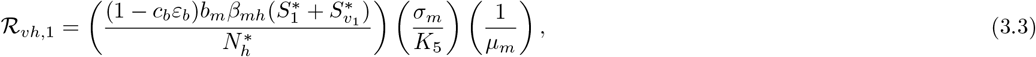

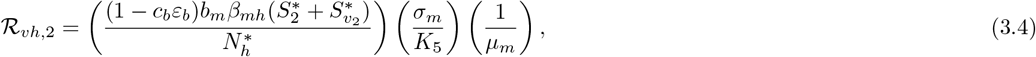

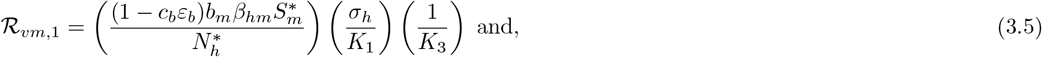

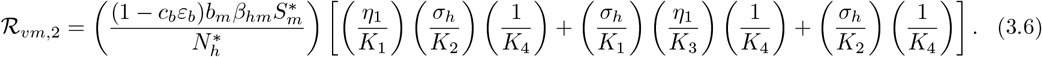

Substituting (3.3)–(3.6) into (3.2), and simplifying, gives:

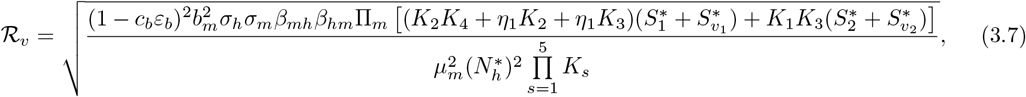

where, 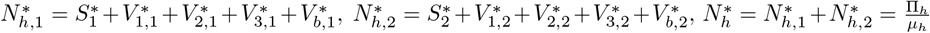, and *K*_*s*_ (with *s* = 1, …, 5) are as defined above. The result below follows from Theorem 2 in [88].

#### Theorem 3.1.

*The disease-free equilibrium*, ℰ_0_, *of the model* (2.2)*–*(2.6) *is locally-asymptotically stable whenever* ℛ_*v*_ < 1, *and unstable whenever* ℛ_*v*_ > 1.

The threshold quantity ℛ_*v*_ is the *control reproduction number* of the model. It measures the average number of new cases generated by a single infectious individual (or mosquito) introduced into a susceptible host (mosquito and human) populations where a certain proportion of humans are vaccinated and/or consistently sleep under a bednet. Furthermore, ℛ_*vh,i*_ (with *i* = 1, 2) are the constituent control reproduction numbers for the infection of susceptible humans in Group *i* by infectious adult female mosquitoes. Similarly, ℛ_*vm,i*_, with *i* = 1, 2, represents the control reproduction numbers for the infection of susceptible adult female mosquitoes following effective bite on infectious humans in Group *i*.

#### Epidemiological interpretation of the terms in the control reproduction number *R*_*v*_

The quantities, ℛ_*vh*,1_, ℛ_*vh*,2_, ℛ_*vm*,1_, and ℛ_*vm*,2_, used in the expression for the control reproduction number ℛ_*v*_, (3.2), are interpreted epidemiologically as follows.

##### Interpretation of ℛ_*vh*,1_

The term ℛ_*vh*,1_, given by (3.3), is the product of three factors; (i) the infection rate of susceptible individuals in Group 1 by an infectious mosquito near the disease-free equilibrium, given by 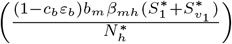; (ii) the probability that an exposed mosquito survives the exposed class (*E*_*m*_) and progresses to the infectious class (*I*_*m*_), given by 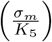; and (iii) the average duration an individual mosquito spends in the infectious class (*I*_*m*_), given by 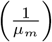.

##### Interpretation of ℛ_*vh*,2_

The term ℛ_*vh*,2_, given by (3.4), is the product of three factors; (i) the infection rate of susceptible individuals in Group 2 by an infectious mosquito near the disease-free equilibrium, given by 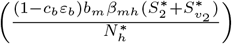; (ii) the probability that an exposed mosquito survives the exposed class (*E*_*m*_) and progresses to the infectious class (*I*_*m*_), given by 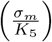; and (iii) the average duration an individual mosquito spends in the infectious class (*I*_*m*_), given by 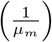.

##### Interpretation of ℛ_*vm*,1_

The term ℛ_*vm*,1_, given by (3.5), is the product of three factors; (i) the infection rate of susceptible mosquitoes by an infectious human in Group 1 near the disease-free equilibrium, given by 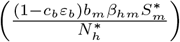; (ii) the probability that an exposed individual in Group 1 survives the exposed class (*E*_1_) and progresses to the infectious class (*I*_1_), given by 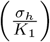; and (iii) the average duration an individual in Group 1 spends in the infectious class (*I*_1_), given by 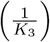.

##### Interpretation of ℛ_*vm*,2_

The term ℛ_*vm*,2_, given by (3.6), consists of two components. The first component represents the infection of susceptible mosquitoes by an infectious individual from Group 1, who subsequently matures into the corresponding infectious individual of Group 2 near the disease-free equilibrium. This component is expressed as the product of two factors; (i) the infection rate of susceptible mosquitoes by an infectious individual in Group 2 near the disease-free equilibrium, given by 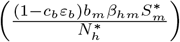; and (ii) the sum of two terms. (a) The first term is the product of the probability that an exposed individual from Group 1 matures into the exposed class of Group 2, given by 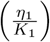; the probability that an exposed individual from Group 2 survives the exposed class (*E*_2_) and progresses to the infectious class (*I*_2_), given by 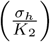; and the average duration in the infectious class (*I*_2_) of Group 2, given by 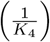. (b) The second term is the product of the probability that an exposed individual in Group 1 survives the exposed class (*E*_1_) and progresses to the infectious class (*I*_1_), given by 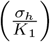; the probability that an infectious individual from Group 1 matures into the infectious class of Group 2, given by 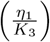; and the average duration in the infectious class (*I*_2_) of Group 2, given by 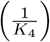. Furthermore, the second component of ℛ_*vm*,2_ represents the infection of susceptible mosquitoes by an infectious individual from Group 2 who progressed to the infectious class (*I*_2_) near the disease-free equilibrium. This is the product of; the infection rate of susceptible mosquitoes by an infectious individual in Group 2 near the disease-free equilibrium, given by 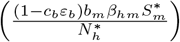; the probability that an exposed individual in Group 2 survives the exposed class (*E*_2_) and progresses to the infectious class (*I*_2_), given by 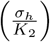; and the average duration in the infectious class (*I*_2_) of Group 2, given by 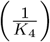.

The epidemiological implication of Theorem 3.1 is that a small influx of malaria-infected individuals will not lead to a large outbreak (i.e., malaria can be eliminated from the community) if control interventions, such as vaccination and the use of bednet, are able to reduce and maintain the control reproduction number (ℛ_*v*_) at a value less than unity. In the absence of vaccination and other control strategies (such as the use of bednet), the control reproduction number (ℛ_*v*_) reduces to the *basic reproduction number* (ℛ_0_)), given by:

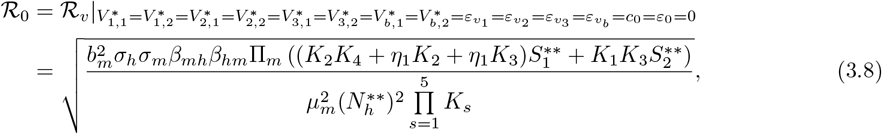

with 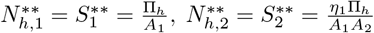, and 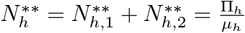. The basic reproduction number measures the average number of new cases generated by a single infectious individual (or mosquito) introduced into a completely susceptible host (mosquito and human) populations. Table 7 depicts the values of the basic and control reproduction numbers of the one-group (homogeneous) and two-group (heterogeneous) models for the various dosing scenarios, showing that the basic and control numbers of the heterogeneous model are consistently lower than those for the homogeneous model.

## 4 Sensitivity Analysis of the Model

Although the values of all the parameters of the model (2.2)–(2.6) are either known from the literature or estimated by fitting the model to data (as given in Tables 3 and 4), uncertainties are expected to occur in these estimates [90]. Hence, it is essential to assess how such uncertainties impact the sensitivity of the parameters of the model with respect to a chosen response function (such as the value of the control reproduction number). In particular, Latin Hypercube Sampling method [91] will be used to conduct a global sensitivity analysis to determine the parameters that have the highest impact on the value of a chosen response function [90, 92]. This method entails defining each parameter in the response function as a distribution, with a range from which samples are drawn without replacement [90, 91]. In this study, the control reproduction number (ℛ_*v*_) is chosen as the response function (since it is a standard metric for quantifying the disease burden in the community [87–89]). It is convenient to introduce the following composite parameters (where the parameters *β*_*hm*_, *β*_*mh*_, *b*_*m*_, *c*_*b*_ and *ε*_*b*_ are as defined in Tables 3 and 4):

a. the effective malaria transmission rate from an infectious human to a susceptible mosquito 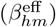, defined by 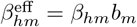,
b. the effective malaria transmission rate from an infectious mosquito to a susceptible human 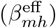, defined by 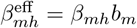, and
c. the overall bednet effectiveness (bed^eff^), defined by bed^eff^ = *c*_*b*_*ε*_*b*_.

Using these definitions, the control reproduction number, defined in (3.7), can now be re-written as:

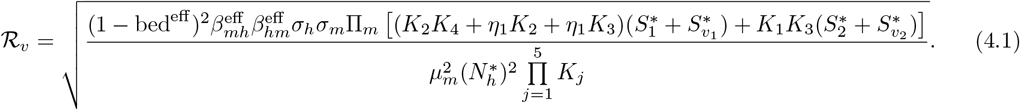

Since the values of the demographic parameters (Π_*h*_, *r*, and *µ*_*h*_) are precisely known from census data [67–69,80] and the values of the three delay parameters (*τ*_12_, *τ*_23_, and *τ*_3*b*_) are also fixed (according to the strict R21/Matrix-M vaccination schedule approved by WHO [34]), the sensitivity analysis will be conducted on the 22 parameters in the expression for ℛ_*v*_ given in (4.1). For the PRCC computation, it is assumed that each parameter in the expression for the response function follows a uniform distribution, and samples of the values of each parameter are drawn from its range given in Tables 3, 4, and 5 (it should be noted that the baseline values for the three composite parameters given in Table 5 were obtained by multiplying the corresponding baseline values of their constituent parameters, as given in Tables 3 and 4, and their ranges were obtained by evaluating these products over the sampled ranges of the constituent parameters). Specifically, for the PRCC computation, the range of each parameter is sub-divided into 1, 000 equally spaced intervals, and parameter values are sampled from these sub-intervals without replacement, giving a 1, 000 × 22 parameter matrix (hypercube). Parameters with PRCC values close to +1 or −1 indicate strong positive or negative correlations with the selected response function, respectively. A general rule of thumb is that a parameter is regarded as highly influential if its PRCC value is at least 0.5 in magnitude [92].

**Table 5.**
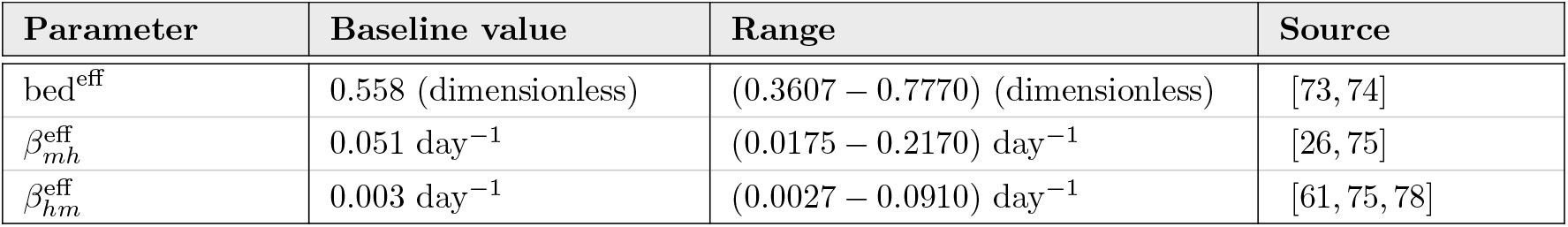
Baseline values and ranges of the composite (effective) parameters in the expression for the control reproduction number (ℛ_*v*_), given by (4.1), used in the PRCC computations.

| Parameter | Baseline value | Range | Source |
| --- | --- | --- | --- |
| $\text{bed}^{\text{eff}}$ | 0.558 (dimensionless) | (0.3607 – 0.7770) (dimensionless) | [73, 74] |
| $\beta_{mh}^{\text{eff}}$ | 0.051 $\text{day}^{-1}$ | (0.0175 – 0.2170) $\text{day}^{-1}$ | [26, 75] |
| $\beta_{hm}^{\text{eff}}$ | 0.003 $\text{day}^{-1}$ | (0.0027 – 0.0910) $\text{day}^{-1}$ | [61, 75, 78] |

**Table 6.** Summary of HIT requirements for the current (i.e., individuals in Group 1) and proposed extended malaria vaccination programs in Burkina for the case where only the first-dose is administered (note that the current malaria vaccination program involves giving all four doses to individuals in Group 1).

| Vaccination program | Total population considered | HIT requirement | Approx. number to be vaccinated | Model |
| --- | --- | --- | --- | --- |
| Homogeneous model (current program, with only the first dose administered) | Children 0 to 3 years of age (Group 1 only) | Vaccinate at least 96.53% of the total population (i.e., $v_{1,1}^* \geq 0.9653$) | $\approx 2.1$ million | Eq. (E.1) |
| Heterogeneous model (proposed extended program, with only the first dose administered) | Children 0 to 5 years of age (Groups 1 and 2) | Either vaccinate:<br>(i) At least 75% of Group 1 and no one in Group 2 (that is, set $v_{1,1}^* \geq 0.75$ and $v_{1,2}^* = 0$) or<br>(ii) At least 46% of Groups 1 and 2 ($n_1^* v_{1,1}^* + n_2^* v_{1,2}^* \approx 0.46$) | $\approx 1.7$ million | Eq. (I.1) |

**Table 7.** Comparison of control and basic reproduction numbers of the various models considered in this study under various vaccination (and dosing) scenarios. The expressions for the reproduction numbers for the various scenarios are given in Appendix C. The values of all parameters used in generating this table are given in Tables 3 and 4. In the computation of the control reproduction numbers, the bednet coverage and efficacy are set at their baseline values (i.e., (*c*_*b*_, *ε*_*b*_) = (0.657, 0.85)). Notations: 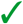 = vaccinated; 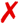= not vaccinated; N/A = not included.

| Scenario | Group 1 vaccination | Group 2 vaccination | Basic reproduction number | Control reproduction number | Model |
| --- | --- | --- | --- | --- | --- |
| 1 | ✓ (1<sup>st</sup> dose) | N/A | 3.13 | 1.25 | Eq. (E.1) |
| 2 | ✓ (4 doses) | N/A | 3.13 | 1.06 | Eqs. (F.1)–(F.3) |
| 3 | ✓ (1<sup>st</sup> dose) | × | 2.61 | 1.08 | Eq. (G.1) |
| 4 | ✓ (4 doses) | × | 2.61 | 0.98 | Eqs. (H.1)–(H.4) |
| 5 | ✓ (1<sup>st</sup> dose) | ✓ (1<sup>st</sup> dose) | 2.61 | 1.03 | Eq. (I.1) |
| 6 | ✓ (4 doses) | ✓ (4 doses) | 2.61 | 0.85 | Eqs. (2.2)–(2.6) |

The result of the PRCC computations, with respect to the response function (ℛ_*v*_), are depicted in Figure 5 (see also Table 8 in Appendix J), from which it follows that the top-five parameters that have the highest influence on the value of the response function ℛ_*v*_ (and, hence, the malaria burden in the population) are:

**Table 8.** Partial rank correlation coefficient (PRCC) values for the parameters (excluding the demographic and delay parameters) of the model (2.2)–(2.6) are computed at day 365, using the control reproduction number (ℛ_*v*_), as the response functions. PRCC values with an absolute magnitude greater than 0.5 are shown in bold, denoting strong correlations (positive and negative correlations indicated by ‘+’ and ‘−’, respectively). Furthermore, the associated *p*-values are computed for each parameter-response function pair, with statistical significance levels are denoted as follows: ‘^*†*^’ for *p* > 0.05, ‘*’ for 0.01 < *p* < 0.05, ‘**’ for 0.001 < *p* < 0.01, and ‘***’ for *p* < 0.001. Except for bed^eff^ (= *c*_*b*_*ε*_*b*_), *p*_12_, *p*_23_, *p*_3*b*_, and 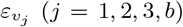, which are dimensionless, all other parameters (including 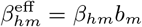 and 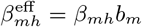) have baseline values and ranges expressed in units of *per* day, as given in Tables 3, 4, and 5.

| Parameter | Baseline value | PRCC ($\mathcal{R}_v$) |
| --- | --- | --- |
| $\omega_b$ | 0.003154 | $-0.0022$ † |
| $\omega_n$ | 0.0027 | $+0.0176$ † |
| $\text{bed}^{\text{eff}}$ | 0.558 | **$-0.7301$** *** |
| $\beta_{mh}^{\text{eff}}$ | 0.051 | **$+0.8148$** *** |
| $\eta_1$ | 0.000853 | $+0.1636$ *** |
| $\eta_2$ | 0.00131 | $+0.1434$ *** |
| $\xi_v$ | 0.00244 | $-0.0350$ † |
| $p_{12}$ | 0.5 | $-0.0429$ † |
| $p_{23}$ | 0.5 | $+0.0582$ † |
| $p_{3b}$ | 0.5 | $+0.0161$ † |
| $\varepsilon_{v_1}$ | 0.4942 | $-0.0828$ ** |
| $\varepsilon_{v_2}$ | 0.6747 | $-0.0829$ ** |
| $\varepsilon_{v_3}$ | 0.72 | $-0.0757$ * |
| $\varepsilon_{v_b}$ | 0.67 | $-0.0259$ † |
| $\sigma_h$ | 0.083 | $+0.0762$ * |
| $\gamma_h$ | 0.0025 | **$-0.8065$** *** |
| $\delta_{h,1}$ | 0.000385 | $-0.0130$ † |
| $\delta_{h,2}$ | 0.00001 | $+0.0108$ † |
| $\Pi_m$ | 2,061,689 | $+0.2401$ *** |
| $\beta_{hm}^{\text{eff}}$ | 0.003 | **$+0.8770$** *** |
| $\sigma_m$ | 0.1 | $+0.3570$ *** |
| $\mu_m$ | 0.0526 | **$-0.7849$** *** |

**Figure 5.**
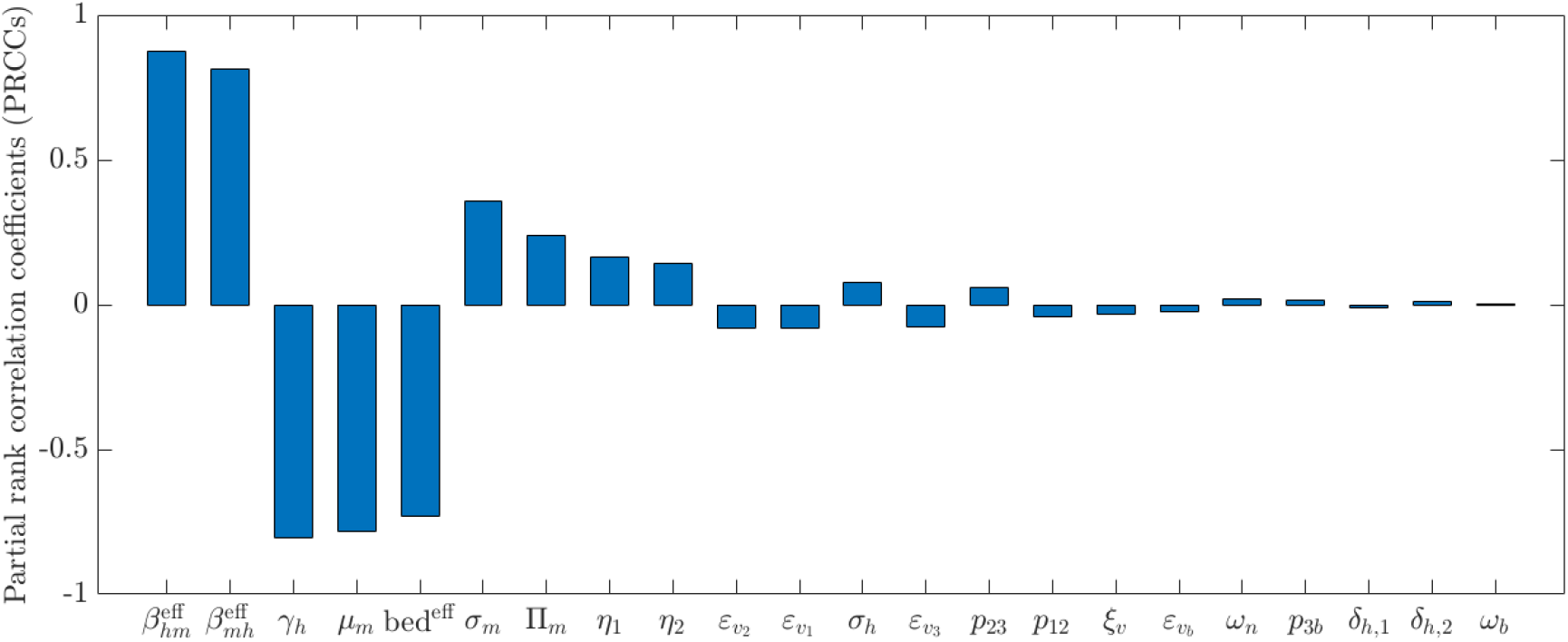
Partial rank correlation coefficients (PRCCs) of the parameters in the chosen response function (ℛ_*v*_), given by (4.1), of the model (2.2)–(2.6). The baseline values and ranges of the parameters used are as given in Tables 3, 4, and 5. All parameters are assumed to follow a uniform distribution.

a. The effective transmission rate of malaria from an infectious human to a susceptible mosquito (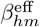; PRCC=+0.877).
b. The effective transmission rate of malaria from an infectious mosquito to a susceptible human (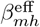; PRCC=+0.8148).
c. The recovery rate of infectious humans (*γ*_*h*_; PRCC=−0.8065).
d. The natural death rate of adult female mosquitoes (*µ*_*m*_; PRCC=−0.7849).
e. The overall bednet effectiveness parameter (bed^eff^; PRCC=−0.7301).

The implication of these results is that public health strategies that decrease (increase) the values of the positively (negatively)-correlated parameters will decrease malaria burden in the community. Specifically, strategies, such as routine anti-malaria vaccination using R21/Matrix-M vaccine with high coverage in the targeted population, using high quality bednets, IRS, insect repellents, larviciding, elimination of mosquito breeding habitats etc., that decrease the transmission parameters (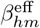 and 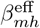) will decrease malaria burden in the targeted population (zero to five-year age group). On the other hand, strategies that increase the values of the negatively-correlated parameters (*γ*_*h*_, *µ*_*m*_ and bed^eff^) will decrease malaria burden. This can be achieved by improved diagnostics and prompt treatment using quality anti-malarial drugs, such as the artemisinin-based combination therapies (increase *γ*_*h*_), using improved vector control strategies, such as IRS, LLINs, and using natural predators like bats, dragonflies, birds, frogs, and lizards etc. (increase *µ*_*m*_), and using high quality bednet with high coverage (increase bed^eff^). It is expected that vaccination against malaria also increases recovery in vaccinated individuals who acquired breakthrough infection [93].

## 5 Computation of Herd Immunity Threshold (HIT)

Although, the Phase 3 trial conducted in Burkina Faso administered vaccination to children between the ages of 5 months and 3 years [33], in reality, vaccinating all susceptible children under the 3 years of age is rarely feasible for various reasons, such as inefficient vaccine delivery and services, the cost of vaccine introduction and sustaining all four doses, shortages of trained personnel and facilities, or parental refusal or unwillingness to vaccinate their children [94–96]. Consequently, it is crucial to determine the minimum proportion of the susceptible members of the community that must be vaccinated in order to provide indirect protection to those who cannot (or are not) vaccinated (i.e., it is necessary to determine the threshold for achieving vaccine-induced herd immunity [97, 98]). This is done in this section. Specifically, since the Phase 3 clinical trials conducted in Burkina Faso only covered children of ages 5 months to 3 years (representing the age group at the highest risk of severe malaria and death) [99], we will first compute the *herd immunity threshold* for this setting (i.e., where only individuals in Group 1 are vaccinated), for the case without (*c*_*b*_ = *ε*_*b*_ = 0) and with (0 ≤ *c*_*b*_ < 1 and 0 < *ε*_*b*_ < 1) bednet usage in the community. Furthermore, the herd immunity threshold for Group 1 vaccination will be computed for the case where members of this group received only the first dose or all four doses.

### 5.1 Vaccine-induced HIT for Group 1-only model

In this subsection, the vaccine-induced HIT will be rigorously derived for the special case of the model (2.2)–(2.6) where the total population consists of Group 1 individuals only (i.e., no Group 2 individuals in the population), and that these individuals are vaccinated either with the first dose only or with all four doses of the R21/Matrix-M vaccine.

#### 5.1.1 Vaccination with first dose only

The equations for the Group 1-only model, where only the first dose is administered, are given in Appendix E (equation (E.1)). The biologically-feasible region of this model is given by:

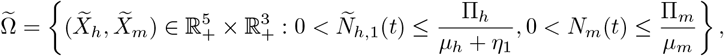

where 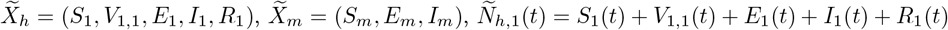, and *N*_*m*_(*t*) = *S*_*m*_(*t*) + *E*_*m*_(*t*) + *I*_*m*_(*t*), and the disease-free equilibrium of this special case is given by:

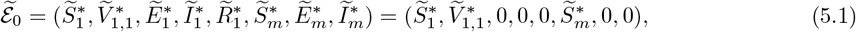

with 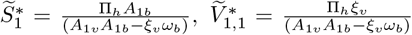, and 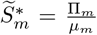 (note that the quantities *A*_1*b*_ and *A*_1*v*_ are defined in Section 3, and that 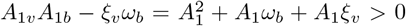). The associated next generation matrices are:

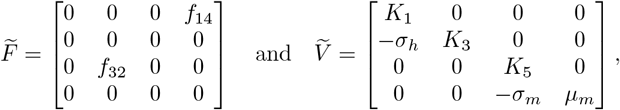

where, 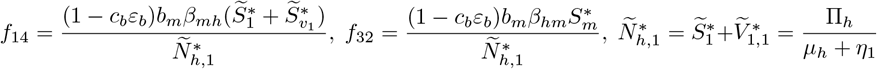, and 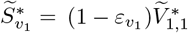 (with *K*_1_, *K*_3_ and *K*_5_ as defined in Subsection 3.1). It can be shown that, for this special case,

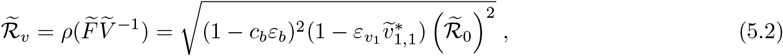

where 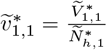 represents the proportion of susceptible individuals in Group 1 who received the first dose of the R21/Matrix-M vaccine (only) at the disease-free equilibrium, and 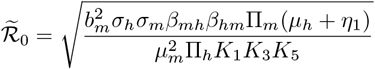. The HIT is computed by setting 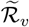 to one and solving for the vaccination coverage 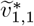. Doing so gives:

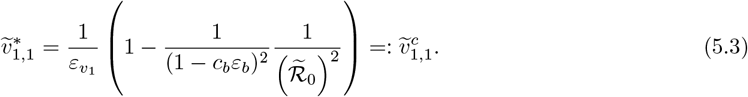

It should be noted that, since 0 < (1 − *c*_*b*_*ε*_*b*_) ≤ 1 and 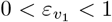, the quantity 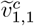 is strictly positive and less than one (i.e., 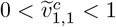), provided the following inequalities hold, respectively:

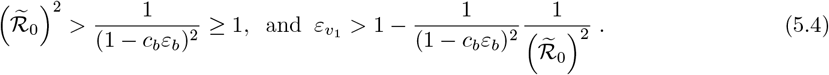

It follows from equation (5.2) that 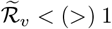 if 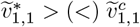, with equality holding when 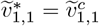. This result is summarized as follows.

##### Theorem 5.1.

*Consider the special case of the model* (2.2)*–*(2.6) *where the total population consists of Group 1 individuals only (i*.*e*., *Group 2 individuals are excluded) and only the first dose of the R21/Matrix-M vaccine is administered, given by* (E.1) *of Appendix E*. *Furthermore, suppose the inequalities in* (5.4) *hold (so that*, 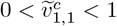*). Vaccine-induced herd immunity can be achieved in the population if the vaccine coverage at steady-state* 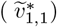 *exceeds the threshold value* 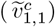, *given by* (5.3).

Theorem 5.1 implies that vaccine-induced herd immunity can be achieved under the current vaccination program in Burkina Faso (note that bednets are also used), which targets only children in the 0-3 year age bracket (i.e., Group 1) [33], if the steady-state vaccine coverage 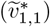 exceeds the HIT 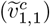.

Figure 6 depicts contour plots of the control reproduction number 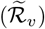 of the Group 1-only model (E.1), as a function of the steady-state vaccine coverage 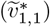 and efficacy 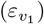, showing a marked decrease in 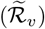 with increasing vaccine coverage and efficacy (as expected). For the case where bednets are not used (i.e., *c*_*b*_ = *ε*_*b*_ = 0) and the vaccine efficacy is at the baseline value of 49.42%, the contours in Figure 6 (a) show that, although the vaccination program significantly reduces the disease burden (as measured in terms of reduction in the reproduction number; see Table 7), it is unable to eliminate the disease (i.e., herd immunity cannot be attained in this case even if 100% of the population (i.e., 100% of Group 1) is vaccinated). Specifically, for this case (with 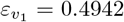), the herd immunity threshold, computed from (5.3), is 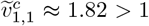 (this is due to the fact, for this scenario, 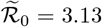, and, consequently, the second inequality in (5.4) does not hold). This result, however, shows that if a vaccine with much higher efficacy is used (e.g., if a new vaccine with efficacy of at least 90% is developed and used to vaccinate the Group 1 individuals), then vaccination alone can lead to elimination of the disease in the Group 1-only population if the coverage in its usage is also high enough (at least 90%). In other words, for the case of the model where the total population consists of Group 1 individuals, vaccination alone (with a highly efficacious vaccine) can lead to malaria elimination in the population even if bednets are not used in the community. However, the development and widespread availability of such a highly efficacious vaccine, coupled with the required very high coverage in its usage (to achieve HIT), seems to be generally impracticable at the current time. Hence, the R21/Matrix-M vaccine needs to be complemented with other traditional vector-based control measures in order to achieve maximum results. In particular, our simulations show, for the case when the R21/Matrix-M vaccine (with the baseline efficacy fixed at 49.42%) is combined with bednet usage (at the baseline efficacy of 85% and coverage set at 65.7% [73, 74]), malaria elimination is feasible if at least 96.53% of the total population (i.e., children in Group 1) received the first dose (Figure 6(b)). Thus, these results show that complementing the aforementioned vaccination program with traditional vector-based control measures increases the likelihood of malaria elimination in children under the age of 3 years in Burkina Faso. Since, as of 2024, Burkina Faso has a strong record of high vaccination coverage against other diseases, such as Bacillus Calmette-Guérin (BCG, 98%), Diphtheria tetanus toxoid and pertussis (DTP, first dose: 95%), Poliomyelitis (first dose: 91%), Rubella (first dose: 88%), and Measles (first dose: 88%) [100], it is possible that the aforementioned coverage level needed to achieve malaria elimination can be realized.

**Figure 6.**
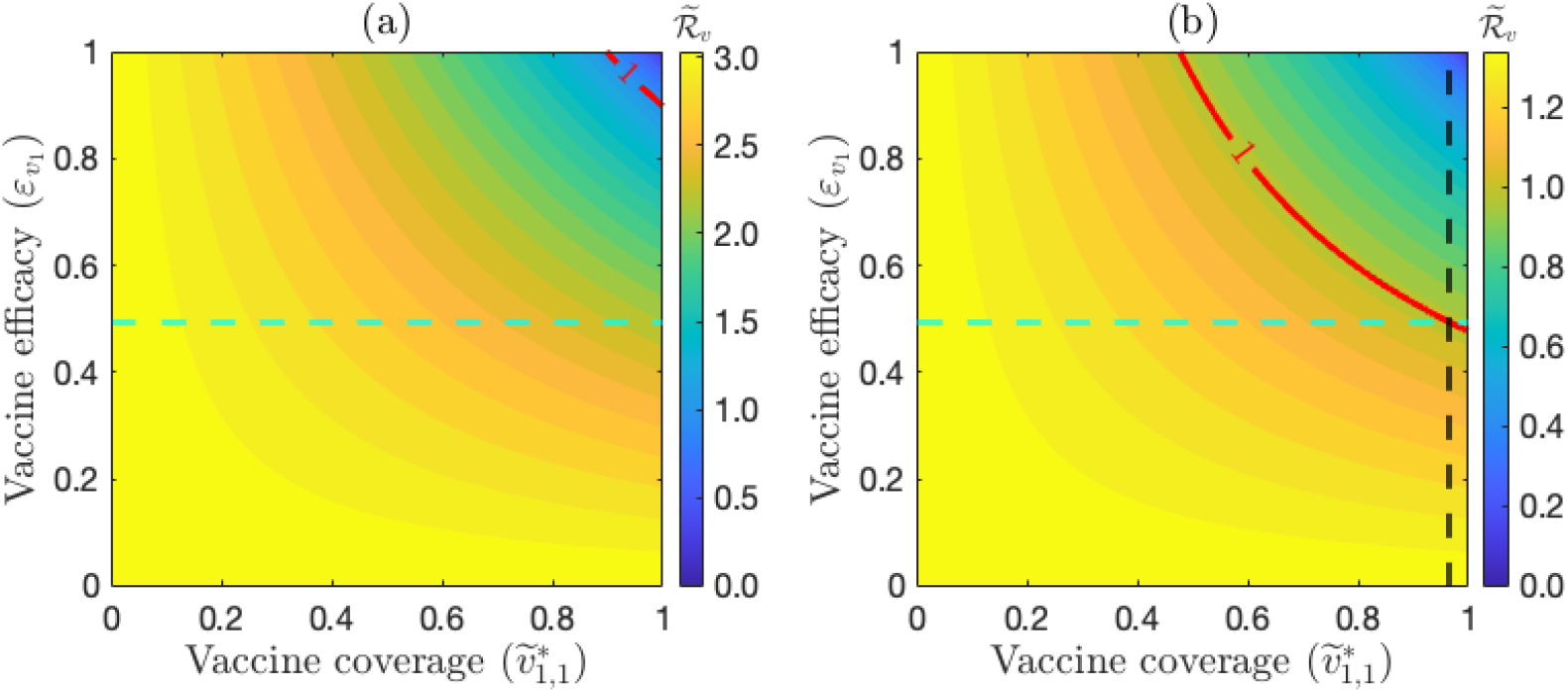
Contour plots of the control reproduction number 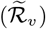 of the model (E.1), as a function of the steady-state vaccine coverage 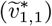 and vaccine efficacy 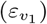, for children in Burkina Faso under the age of 3 (i.e., children in Group 1) who received the first dose (only) of the R21/Matrix-M vaccine. (a) no bednet coverage (i.e., *c*_*b*_ = *ε*_*b*_ = 0), and (b) with bednet usage at baseline levels (i.e., *c*_*b*_ = 0.657 and *ε*_*b*_ = 0.85). The red curve represents the level curve 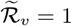. The dashed green horizontal line corresponds to 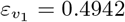, which is the baseline efficacy of the first dose of the R21/Matrix-M vaccine [33]. The dashed black vertical line, drawn through the intersection point of the red curve with the dashed green line, indicate the corresponding vaccine coverage level, 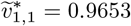, required to achieve the HIT in the targeted group. Parameter values used to generate the contour plots are as given in Tables 3 and 4.

#### 5.1.2 Vaccination with all four doses

We now seek to compute the HIT for the Group 1-only special case of the model (2.2)–(2.6) where these individuals receive all four doses of the vaccine. The equations for this special case of the model are given in Appendix F (equations (F.1)–(F.3)). The DFE of this special case is given by (where 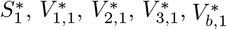, and 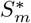 are as defined in Section 3 above):

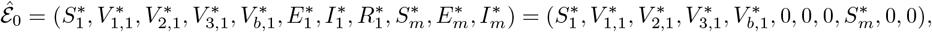

and the associated control reproduction number for this special case of the model is given by:

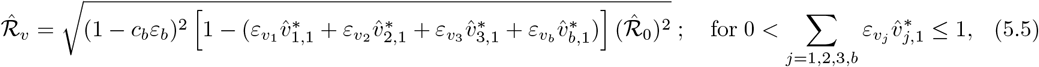

where,

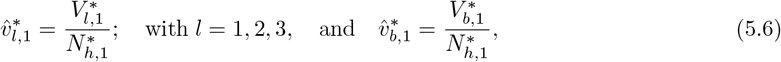

are the proportions of susceptible individuals in Group 1 who received the first *l* doses of the R21/Matrix-M vaccine and those who received all three doses and the booster dose, respectively, and

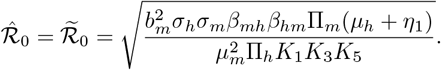

Setting 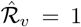 in equation (5.5), and simplifying, gives with 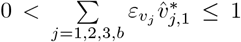 provided 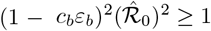:

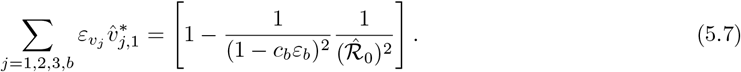

Solving for 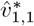 (i.e., the proportion of individuals in Group 1 who received the first dose) from (5.7) gives:

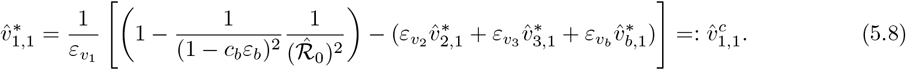

Similarly, solving for the ratio 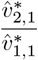 (by solving for 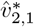 from (5.7) and dividing with 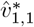), for the proportion of first-dose recipients who went on to receive the second dose, gives:

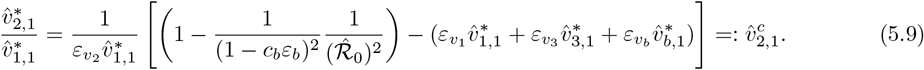

Furthermore, the ratio of second-dose recipients who received the third dose 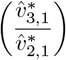, obtained by solving for 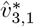 from (5.7) and dividing with 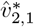, is given by:

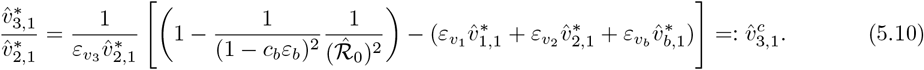

Finally, the ratio of individuals who received all three doses and proceeded to receive the booster dose 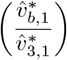, obtained by solving for 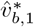 from (5.7) and dividing with 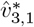, is given by:

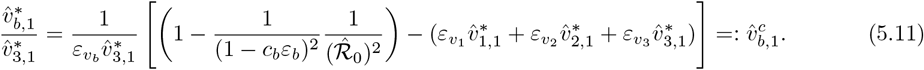

It should be noted that, since 0 < 1 − *c*_*b*_*ε*_*b*_ ≤ 1, and 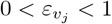 (for *j* = 1, 2, 3, *b*), each of the quantities 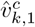 (for *k* = 1, 2, 3, *b*, with *k* ≠ *j*), given, respectively, by (5.8)-(5.11), are strictly positive and less than one (i.e., 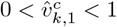, provided the following inequalities hold, respectively with 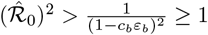:

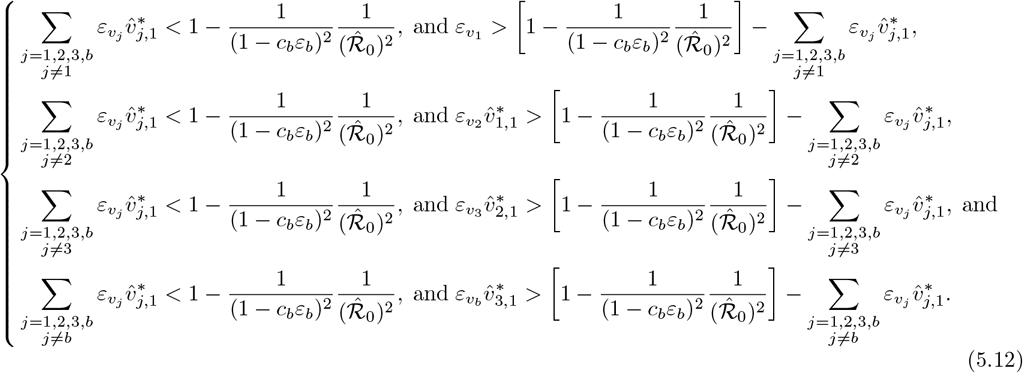

It follows from (5.8)–(5.11) that 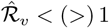 whenever one of the following conditions hold: 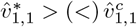, with 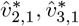, and 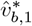 fixed at their respective steady-state baseline values (computed from (5.6) using the baseline parameter values in Tables 3 and 4); 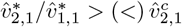, with 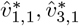, and 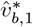 fixed at their respective steady-state baseline values (computed from (5.6) using the baseline parameter values in Tables 3 and 4); 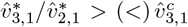, with 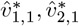, and 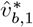 fixed at their respective steady-state baseline values (computed from (5.6) using the baseline parameter values in Tables 3 and 4); or 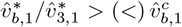, with 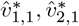, and 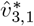 fixed at their respective steady-state baseline values (computed from (5.6) using the baseline parameter values in Tables 3 and 4). The equality holds when 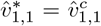, or 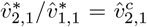, or 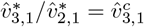, or 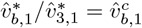. This result is summarized as follows.

##### Theorem 5.2.

*Consider the special case of the model* (2.2)*–*(2.6) *where the total population consists of Group 1 individuals only (i*.*e*., *Group 2 individuals are excluded) and all four doses of the R*21*/Matrix-M vaccine are administered, given by* (F.1)*–*(F.3) *of Appendix F*. *Furthermore, suppose the inequalities in* (5.12) *hold (so that*, 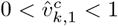, *for k* = 1, 2, 3, *b, with k* ≠ *j* = 1, 2, 3, *b), and* 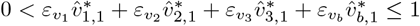 *(so that* 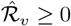*). Vaccine-induced herd immunity can be achieved in the population if the steady-state vaccine coverage associated with the first dose*, 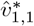, *or the successive dose coverage ratios* 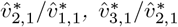, *or* 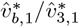, *exceed their corresponding critical threshold values* 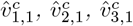, *or* 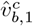 *respectively, as given in* (5.8)*–*(5.11). *Specifically, HIT can be achieved in the population under any of the following four scenarios:*

i. 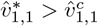 *with* 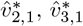, *and* 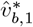 *fixed at their respective steady-state baseline values, or*
ii. 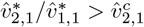 *with* 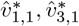, *and* 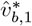 *fixed at their respective steady-state baseline values, or*
iii. 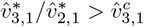 *with* 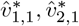, *and* 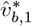 *fixed at their respective steady-state baseline values; or*
iv. 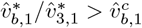 *with* 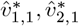, *and* 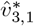 *fixed at their respective steady-state baseline values*.

Like in the case of Theorem 5.1, the epidemiological implication of Theorem 5.2 is that, for the case where all four doses of the R21/Matrix-M vaccine are administered, HIT can be achieved under the current vaccination program in Burkina Faso (which targets children under the age of 3 years) if the steady-state vaccine coverages (or dose coverage ratios) given in Items (*i*) − (*iv*) of Theorem 5.2 exceed their corresponding threshold values.

The results of Theorem 5.2 are numerically illustrated by simulating the Group 1-only model with all four doses administered, given by equations (F.1)–(F.3), using the baseline values of the parameters in Tables 3 and 4. Specifically, Figure 7 depicts contour plots of the associated control reproduction number 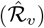, as a function of vaccine efficacy and the steady-state coverage of the *j* (*j* = 1, 2, 3, and *b*) dose(s) of the vaccine, for the case without (Figures 7 (a)-(d)) and with (Figures 7 (e)-(h)) bednet usage in the population (for simulations, we redefined 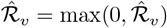 to avoid the regions where the inequality 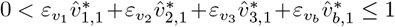 does not hold (i.e., 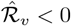), shown in shaded dark blue regions of the plots in Figure 7).

**Figure 7.**
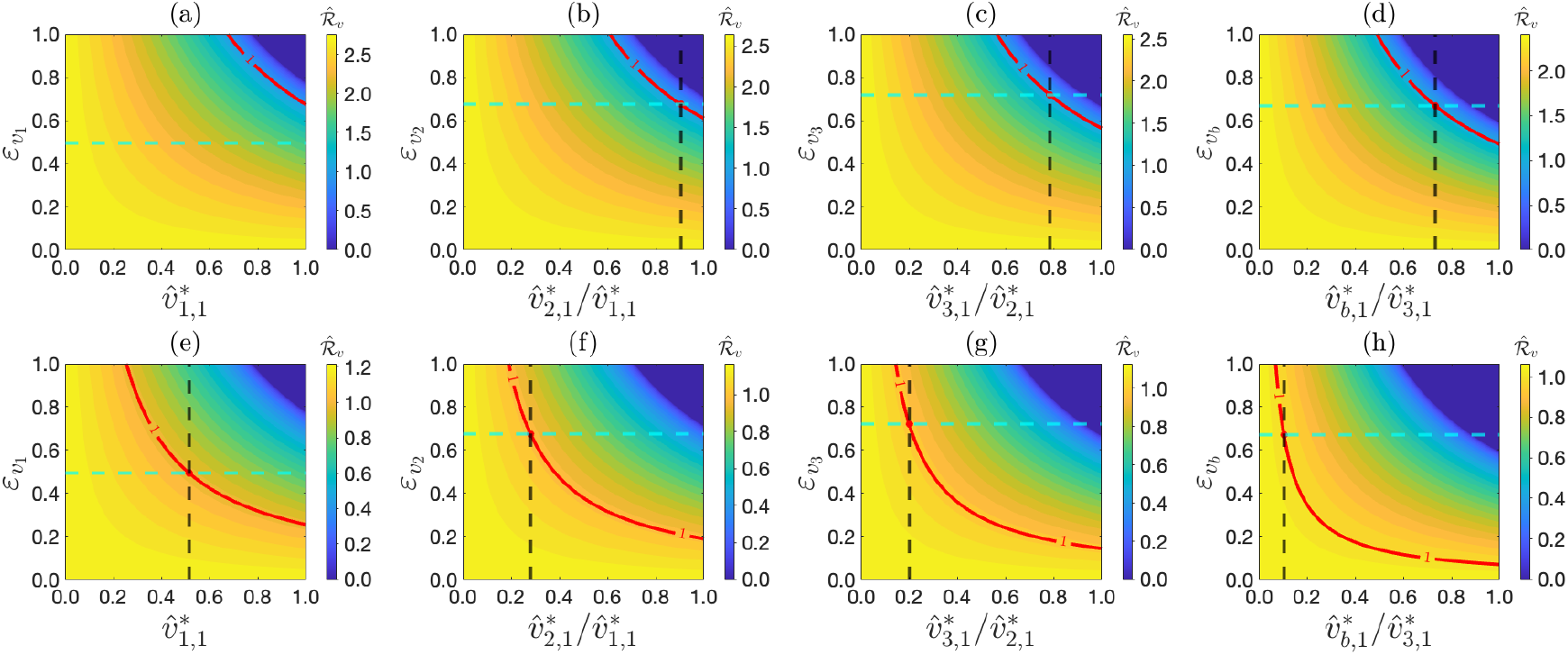
Contour plots of the control reproduction number 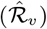 of the Group 1-only model (F.1)–(F.3), as a function of the vaccine efficacy and steady-state coverage of the *j* vaccine doses (*j* = 1, 2, 3, and *b*), administered to children in the 0-3 year age group in Burkina Faso (with all four doses administered) generated using the baseline values of the parameters in 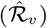 given in Tables 3 and 4. In the simulations, 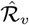 is computed as 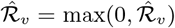 where the inequality 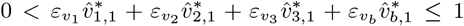 does not hold. The top row (panels (a)–(d)) corresponds to the scenario where bednets are not used in the community (i.e., *c*_*b*_ = *ε*_*b*_ = 0), while the bottom row (panels (e)–(h)) corresponds to the scenario where bednets are used in the community and at their baseline levels (i.e., *c*_*b*_ = 0.657 and *ε*_*b*_ = 0.85). Columns 1–4 explore the impact of varying one vaccine dose at a time, while holding all remaining doses at their respective steady-state baseline values. Column 1 (panels (a), (e)) depicts the first-dose efficacy 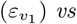. the steady-state first-dose coverage 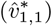 with 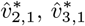, and 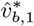 fixed. Column 2 (panels (b), (f)) depicts the second-dose efficacy 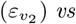. the steady-state dose-coverage ratio 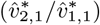 with 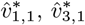, and 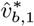 fixed. Column 3 (panels (c), (g)) depicts the third-dose efficacy 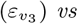. the steady-state dose-coverage ratio 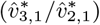 with 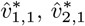, and 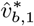 fixed. Column 4 (panels (d), (h)) depicts the booster-dose efficacy 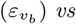. the steady-state dose-coverage ratio 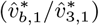 with 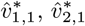, and 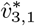 fixed. The red curve represents the level curve 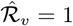. The dashed green horizontal lines in Columns 1 to 4 correspond to the baseline efficacies of the first, second, and third primary doses and the booster dose, respectively, as given in Table 3. The dashed black vertical lines, drawn through the intersection points of the red curves with the dashed green lines, indicate the corresponding steady-state coverage (or successive dose-coverage ratios) required to achieve the herd-immunity threshold in the population. In particular, by substituting the baseline parameter values in Table 3, together with 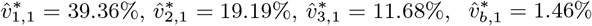, and 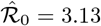, into the expressions in (5.8)-(5.11), the threshold quantities 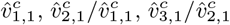, and 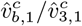 take the values 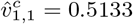 (panel (e)); 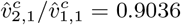 (panel (b)) and 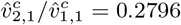 (panel (f)); 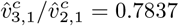 (panel (c)) and 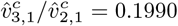 (panel (g)); 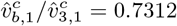 (panel (d)) and 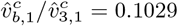 (panel (h)), respectively.

The contours in Figure 7 show a marked decrease in the value of the reproduction number as the vaccine efficacy and coverage increase, regardless of whether or not bednets are used in the community. However, HIT (or elimination) can only be achieved if the baseline efficacy and coverage of each dose is high enough. For instance, for the case without bednet usage, HIT cannot be achieved even if the entire population is vaccinated with all four doses of the vaccine (Figure 7(a)). In this case, HIT, computed from equation (5.8), is 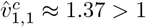 (since 1 − *c*_*b*_*ε*_*b*_ = 1 and 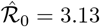); consequently, the second inequality in (5.12) does not hold. However, this figure shows that malaria can be eliminated if the efficacy of the first dose is increased to at least 67% (while maintaining the other doses at their baseline efficacy values) provided the coverage is high enough (nearly 100%). Completing the vaccination program, for this scenario, with bednet usage at baseline level (i.e., with efficacy and coverage set at 85% and 65.7%, respectively [73, 74]), elimination can be achieved if at least 51.33% of the population received the first dose at steady-state (Figure 7(e)), with the remaining doses (i.e., doses 2,3, and booster) maintained at their respective steady-state levels.

Elimination can be achieved without bednet usage if at least 90.36% of first-dose recipients receive the second dose (Figure 7(b)), or 78.37% of second-dose recipients received the third-dose (Figure 7(c)) or 73.12% of the third-dose recipients received the booster dose (Figure 7(d)), while all other doses are fixed at their respective baseline values. When bednets are used in the population (at baseline levels), the aforementioned coverages needed to achieve elimination decrease dramatically to 27.96% when first-dose recipients received the second dose (Figure 7(f)), or to 19.90% when second-dose recipients get the third dose (Figure 7(g)) or to 10.29% when third-dose recipients received the booster dose (Figure 7(h)). Thus, the simulations in Figure 7 show that malaria can be eliminated in the Group 1-only population using vaccination alone (with all vaccinated individuals receiving all four doses of the R21/Matrix-M vaccine) provided the vaccination coverage is high enough (ranging from 73% − 90%, as depicted in Figures 7 (b), (c), and (d)). Using bednets, even at their baseline levels, drastically reduces the vaccination coverage needed to achieve elimination (down to between 10% to 51%). In other words, combining the vaccination program with bednet usage (even at baseline level) dramatically increases the likelihood of malaria elimination in the 0-3 year age group in Burkina Faso.

However, it should be mentioned that, although the current malaria vaccination program in Burkina Faso focuses primarily on vaccinating children in the 0-3 year age group (i.e., individuals in Group 1), considered to be the highest-risk group to malaria morbidity and mortality [33, 99], children of ages 3-5 years are also at high risk of malaria morbidity and mortality [2]. Hence, it is instructive to theoretically evaluate the impact of vaccinating children in the 3-5-year age group (i.e., children in Group 2 of our model), in addition to children in Group 1, with the goal of potentially enhancing protection for the entire vulnerable group of Children under 5 years of age. Specifically, it is instructive to determine the HIT for this scenario (where children in Groups 1 and 2 are vaccinated). The derivation of the HIT for this scenario is done (for mathematical tractability) for the special case of the full (two-group) model (2.2)–(2.6) where individuals in each group received only the first dose, as described below.

### 5.2 HIT for the case where both groups are vaccinated: first dose only

In this subsection, HIT will be computed for the special case of the model (2.2)–(2.6) where individuals in both groups are vaccinated with the R21/Matrix-M vaccine, but all vaccinated individuals receive only the first dose, with bednet usage at baseline levels (i.e., *c*_*b*_ = 0.657 and *ε*_*b*_ = 0.85). The equations for this special case of the model are given by (I.1) in Appendix I. The objective here is to determine the optimal values of the steady-state vaccination coverages, 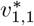 (for Group 1) and 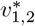 (for Group 2) defined in (5.6), required to minimize the total proportion of vaccinated individuals 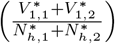 in both groups which also ensures 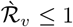 (see (C.9) in Appendix C), so that the disease can be eliminated in both groups. In other words, the objective is to determine the optimal values of 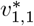 and 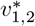 such that the total proportion of vaccinated individuals at steady-state, given by

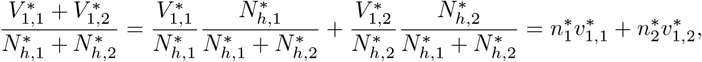

is minimized and that the associated control reproduction number 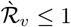. Formally, the objective is to solve the following optimization problem:

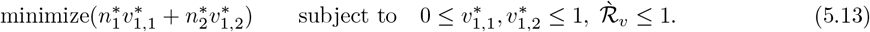

We solve this optimization problem using the geometric technique introduced in [101]. It can be seen, by setting 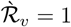 in (C.9) and solving for 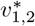 that:

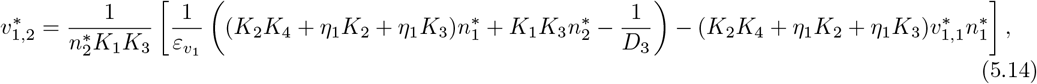

where 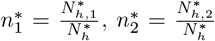, and 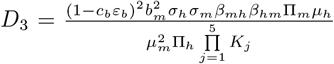. Since 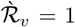 is a linear function of 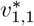 and 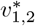 (see equation (5.14)), the optimization problem (5.13) is linear. Figure 8 depicts the constraint line 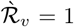, as a function of 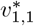 and 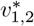, given in (5.14), and the level curves of the objective function, 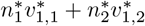. It follows from this figure that the constraint line 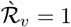 (orange line in Figure 8) intersects the 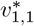 and 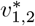 axes precisely at the following two points (note that the 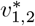 −intercept falls outside the constraint 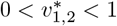):

**Figure 8.**
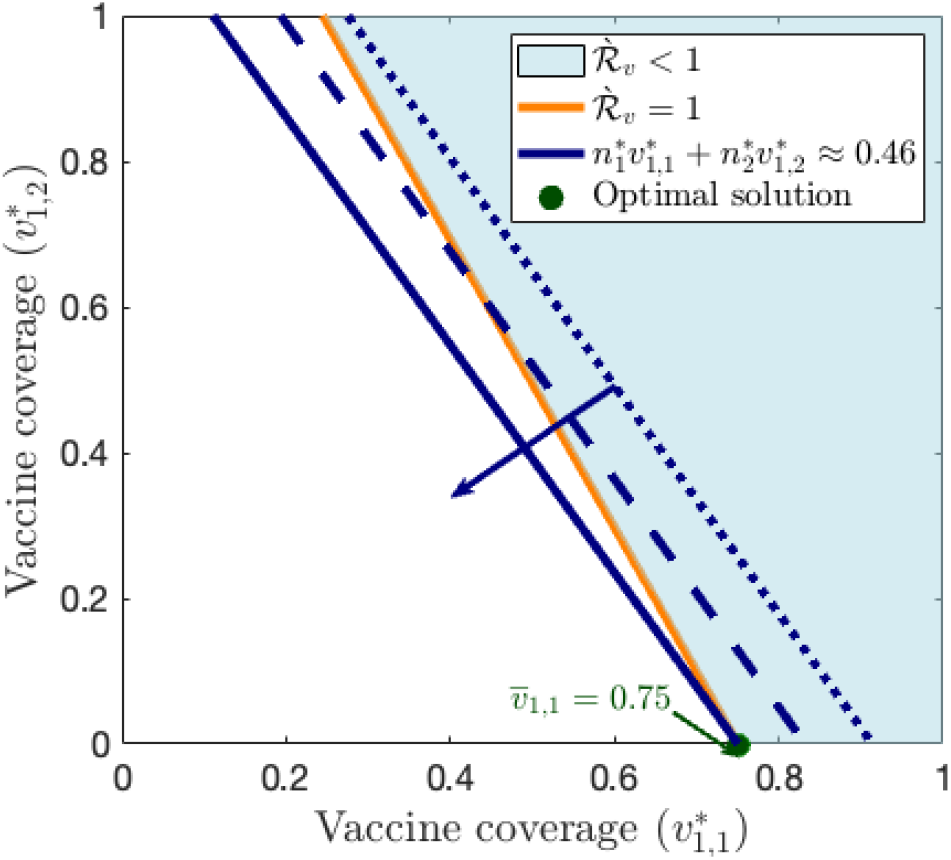
Geometrical illustration (using baseline parameter values) of the optimization problem (5.13) for minimizing the total steady-state first-dose vaccination coverage in both groups, with bednet usage fixed at its baseline levels (*c*_*b*_ = 0.657 and *ε*_*b*_ = 0.85). The orange line is the constraint boundary 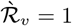, and the blue lines are level sets of the objective function 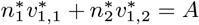, where 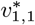 and 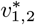 denote the steady-state first-dose vaccination coverages in Groups 1 and 2, respectively, *A* > 0 is the level, 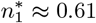 and 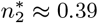 are the corresponding population fractions evaluated at the disease-free steady-state, respectively. The solid blue arrow indicates the direction of decreasing values of the level *A* (minimization), so that the level sets shift toward the origin. The sky-blue shaded region corresponds to the region in the 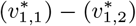 plane where 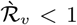. The minimum objective value satisfying the elimination constraint 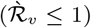 is *A* ≈ 0.46 (46%). The optimal vaccine coverage pair 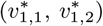 occurs at the 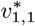-intercept, corresponding to 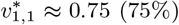 and 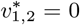 (illustrated with the solid green dot on the 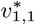 axis). Parameter values used in generating this figure are as given in Tables 3 and 4.

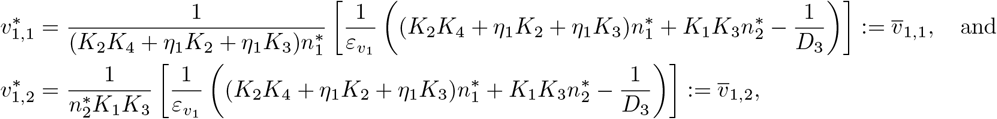

respectively. The slope 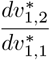 along the constraint line 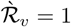 (given by (5.14)) is:

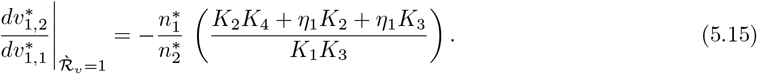

Similarly, the slope along the line 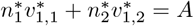 (where *A* > 0 is the level) is given by:

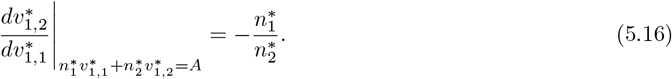

Using the baseline parameter values given in Tables 3 and 4, and noting that 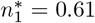 and 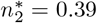 at disease free equilibrium, it follows from (5.15) and (5.16) that

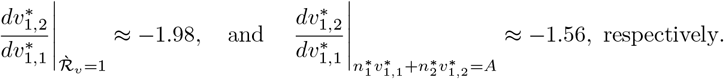

Hence, the orange line in Figure 8 is steeper than the blue lines (since, |− 1.98| > |− 1.56|). Therefore, in line with [101], the optimal solution occurs at the 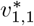-intercept (i.e., 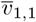), with the corresponding HIT coverage pair of 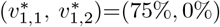, as illustrated by the solid green dot on the 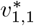 axis of Figure 8. This result shows two main scenarios for achieving HIT. The first is that 75% of children in Group 1 are vaccinated with the first dose only (and children in Group 2 are not vaccinated). The second is to vaccinate 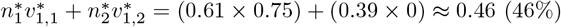 of the total population (of Groups 1 and 2).

Considering Burkina Faso’s high vaccination coverage for childhood diseases, these relatively modest coverage levels for the R21/Matrix-M vaccine (i.e., vaccinating either 75% of Group 1 or 46% of both groups with the first dose) are realistically attainable in Burkina Faso. The HIT requirements when both groups are considered and administered with first dose only (i.e., either vaccinating 75% of Group 1 only or vaccinating 46% of both groups) is equivalent to vaccinating about 1.7 million children of ages 0 to 3 or 0 to 5 years of age, respectively, in Burkina Faso. It should be recalled from Subsection 5.1.1 that when the total population only consists of children in Group 1 (i.e., for the case of the homogeneous model (E.1)), at least 96.53% of these (Group 1) individuals need to be vaccinated to achieve HIT in this group (this is equivalent to vaccinating 2.1 million children in the 0 to 3-year age group). In other words, if the total population targeted for vaccination consists of children in the 0 to 3-year age group (i.e., children of ages 3 to 5 are excluded), more vaccine resources (2.1 million doses) will be needed to achieve HIT in Group 1 (only), in comparison to the scenario where the expanded 0 to 5-year age bracket is considered for vaccination (which achieves HIT in both groups by vaccinating 1.7 million children in this larger age cohort).

Thus, this study shows that the current malaria vaccination program in Burkina Faso, which solely targets children 0 to 3 years of age for vaccination, may be suboptimal compared to an expanded program that additionally administers the vaccine to children of ages 3 to 5, even for the case when only the first dose is administered. Malaria elimination (in both groups) is far more likely in Burkina Faso using the proposed extended strategy (of vaccinating children 0 to 5 years of age), and require less vaccine resources (1.7 million vs. 2.1 million doses), than using the current strategy of only considering children ages 0 to 3 years.

The seeming anomaly or inconsistency related to the 96.53% HIT requirement for Group 1 if the homogeneous model (E.1) is used, compared to the 75% requirement for Group 1 to achieve HIT in both groups (i.e., the case where the heterogeneous model (I.1) is used) can be explained as follows. First, based on the baseline values of the parameters used in our simulations (given in Tables 3 and 4) for the homogeneous model and the heterogeneous model, the values of the basic and control reproduction numbers of the homogeneous model are consistently lower than those for the heterogeneous model (see Table 7), suggesting that lower effort (i.e., fewer vaccine doses) may need to be administered in order to effectively control or eliminate the disease when the heterogeneous model is used, as against the case with the homogeneous model. Secondly, the intuition behind the decline in the value of the basic reproduction number when transitioning from a one-group (homogeneous) to a two-group (heterogeneous) model lies in the structure of the force of infection (see equations (2.1)). In the heterogeneous model, the force of infection from humans to mosquitoes is averaged over the total human population at time *t* (i.e., (*I*_1_(*t*) + *I*_2_(*t*))*/N*_*h*_(*t*) rather than *I*_1_(*t*)*/N*_*h*,1_(*t*), where *N*_*h*_(*t*) denotes the combined human population of Groups 1 and 2, and *N*_*h*,1_(*t*) represents the total human population of Group 1 (see Figure 9(a)). This results in a reduced transmission impact *per* infectious human. Furthermore, in the two-group model, mosquitoes bite individuals from both groups rather than exclusively from Group 1. As a consequence, the mosquito infection burden is also averaged over the total human population of two groups (*I*_*m*_(*t*)*/N*_*h*_(*t*)) rather than averaged over just one human group (*I*_*m*_(*t*)*/N*_*h*,1_(*t*)), as shown in Figure 9(b), resulting in a lower ratio. This can be interpreted as a reduced transmission impact *per* infected mosquito. Together, these effects explain the observed reduction in the basic reproduction number in the two-group (heterogeneous) model. In any case, this result is consistent with the results reported in [101] (and also in [102] and [103]) which show that the HIT for heterogeneous model is lower than that for a corresponding homogeneous model. The simulations are performed using the DDE23 and ODE45 solvers implemented in MATLAB to generate this figure (and all other figures in the numerical simulation section 6) [104–106].

**Figure 9.**
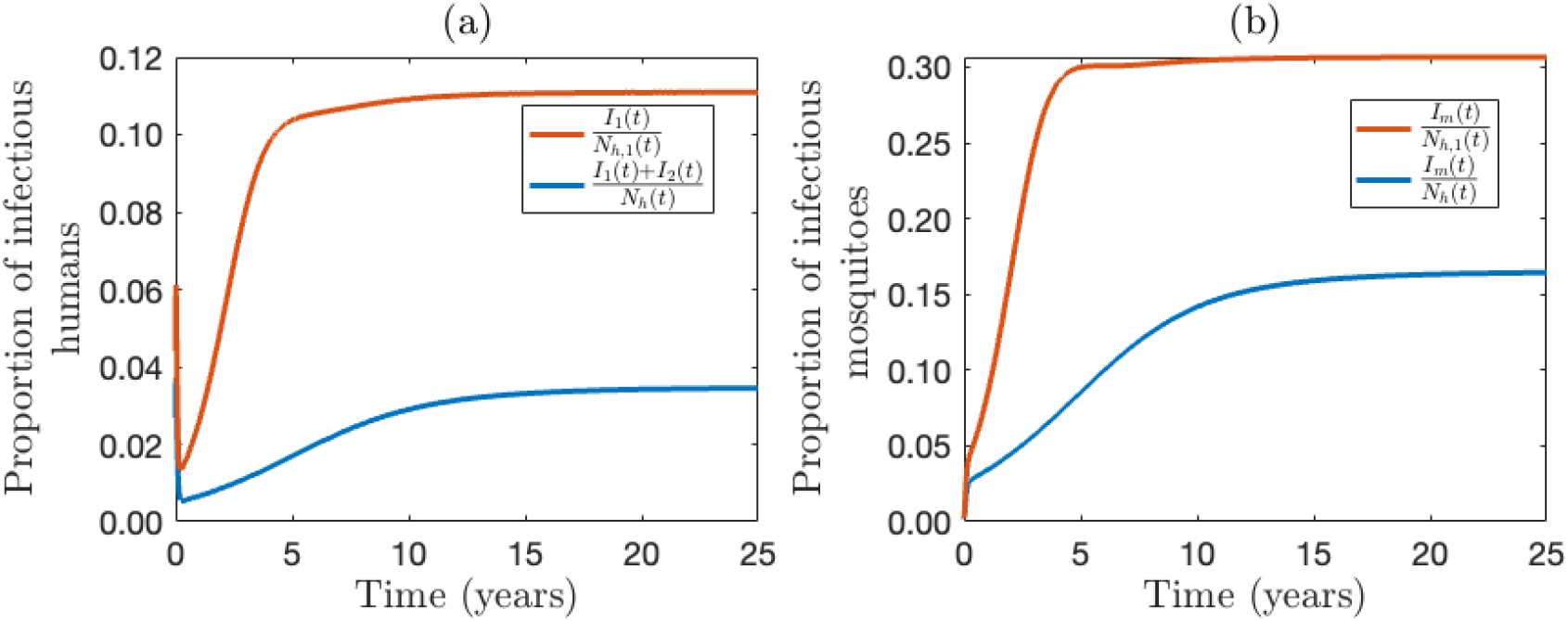
Simulation results for the vaccination-free versions of the homogeneous (F.1)–(F.3) (Group 1 only) and heterogeneous (2.2)–(2.6) (Groups 1 and 2) models. (a) Proportion of infectious humans in Group 1 (*I*_1_(*t*)*/N*_*h*,1_(*t*)), as a function of time, for the homogeneous model (red curve) and the total proportion of infectious humans for the heterogeneous model, (*I*_1_(*t*) + *I*_2_(*t*))*/N*_*h*_(*t*)) (blue curve). (b) Proportion of infectious mosquitoes given by *I*_*m*_(*t*)*/N*_*h*,1_(*t*) for the homogeneous model (red curve) and by *I*_*m*_(*t*)*/N*_*h*_(*t*) for the heterogeneous model (blue curve). Parameter values used in the simulations are as given in Tables 3 and 4.

It is worth mentioning that although the 75% vaccination requirement for Group 1 to achieve HIT in the 0 to 5-year age group using the heterogeneous model is attainable in Burkina Faso (considering its track-record of very high coverage for vaccine-preventable childhood diseases), our simulations show that HIT still be achieved if the 75% target cannot be achieved, provided enough individuals in Group 2 are also vaccinated. The contour plots in Figure 10 show that, for the case where only 34% of Group 1 can get the first dose, HIT can be achieved in the 0 to 5-year age group if at least 81% of Group 2 individuals receive the first dose (Figure 10(a)). Similarly, if only 54% of Group 1 can get the first dose, at least 42% of Group 2 need to be vaccinated with the first dose to achieve HIT (Figure 10(b)). For the more optimistic scenario of 74% (i.e., just slightly below the 75% optimal level) coverage in Group 1 can be achieved, a mere 3% vaccination coverage of Group 2 will lead to HIT (Figure 10(c)). In other words, the contours in Figure 10 show that combining modest coverages of Groups 1 and 2 can also lead to HIT in both groups, confirming that the prospect for malaria elimination in Burkina Faso using the proposed expanded vaccination program (of additionally vaccinating children of 3 to 5 years of age) are remarkably promising with modest coverages.

**Figure 10.**
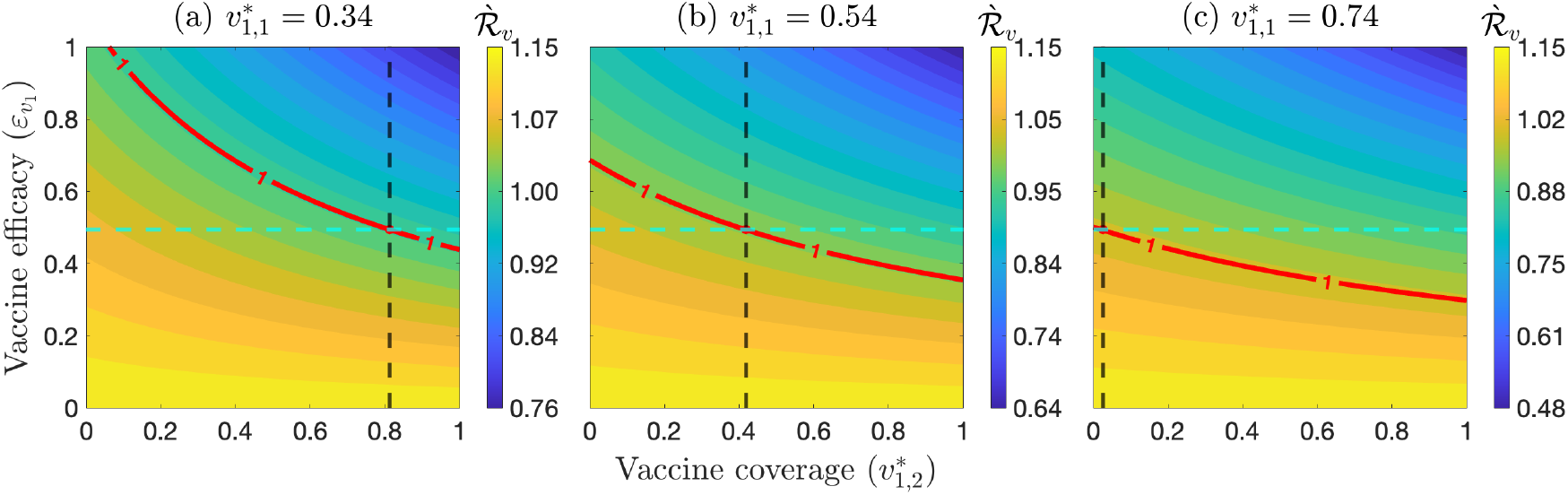
Contour plots of the control reproduction number (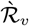; given by (C.9)) for the two-group heterogeneous model (I.1), as a function of efficacy of the first-dose 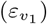 and coverage for individuals in Group 2 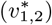, for various fixed values of the first-dose coverage in Group 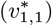, with bednet usage at baseline levels (*c*_*b*_ = 0.657 and *ε*_*b*_ = 0.85). (a) 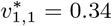; (b) 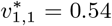, and (c) 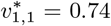. The red curve represents the level curve 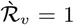. The dashed green horizontal line corresponds to the baseline efficacy of the first-dose 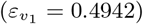. The dashed black vertical line in each panel is drawn through the intersection of the red curve with the dashed green horizontal line, and gives the corresponding steady-state vaccine coverage in Group 2 required to achieve herd immunity (specifically, 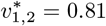 for (a), 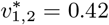 for (b), and 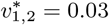 for (c)).

## 6 Assessment of Proposed Extended Strategy for Burkina Faso

In this section, the extended strategy we propose (which, in addition to vaccinating the 0 to 3-year age group, also entails vaccinating the 3 to 5-year age group) will be analyzed by simulating the two-group (heterogeneous) model (2.2)–(2.6) (with bednet usage at baseline level), or its variants (depending on which group is vaccinated and how many doses are administered) using the baseline values of the parameters given in Tables 3 and 4. Specifically, the population-level of the following four vaccination scenarios will be assessed:

### Vaccination Scenario 1

Simulations of the model (2.2)–(2.6) where only individuals in Group 1 are vaccinated (i.e., individuals in Group 2 are not vaccinated), and only the first dose is administered. Here, the relevant sub-model to be simulated is (G.1).

### Vaccination Scenario 2

This is the same Scenario 1, except that vaccinated individuals in Group 1 receive four doses. Here, the relevant model is (H.1)–(H.4).

### Vaccination Scenario 3

Simulations of the model (2.2)–(2.6) where individuals in both groups are vaccinated with the first dose only. Here, the relevant model is (I.1).

### Vaccination Scenario 4

This scenario is the same as Scenario 3, but with all four doses administered (the relevant model here is (2.2)–(2.6)).

The objective is to assess the impact of vaccinating one group on the incidence and disease-induced mortality in the vaccinated group, as well as the potential *spillover* to the other (unvaccinated) group (i.e., assessing whether administering the vaccination program to one group only causes a reduction in disease burden in the unvaccinated group). Since routine use of the R21/Matrix-M malaria vaccine in Burkina Faso began with a national launch on 15 August 2025 (in Pouytenga), marking the scale up of malaria vaccination to all 70 districts in the country (although malaria vaccination was introduced earlier, in February 2024, in 27 districts) [107], our simulations for assessing the effect of vaccination will start from 2025, and the simulations will run for 10 years (until 2035). The initial conditions for the state variables of the various models used in these simulations are obtained by simulating them during the pre-vaccination period of 2015-2024.

### 6.1 Assessing the Impact of Vaccination Scenario 1

The impact of the Vaccination Scenario 1 is assessed by simulating the model (G.1), given in Appendix G, using the parameter values in Tables 3 and 4 (for these parameter values, the corresponding steady-state first dose vaccination coverage is 37.58% (i.e., 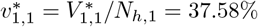). The simulation results obtained, depicted in Figure 11, show a substantial reduction in the cumulative number of new cases (Figure 11(a), blue curve) and disease-induced mortality (Figure 11(b), blue curve) in Group 1, in comparison to the vaccination-free scenario (illustrated by red curves). Specifically, under this scenario, up to 20.07% and 19.03% reduction in the cumulative number of new cases and mortality can be recorded, respectively, by the end of the simulation period (2035). These simulations also show a modest reduction (i.e., spillover) in the cumulative number of new cases (Figure 11(c)) and mortality (Figure 11(d)) in the unvaccinated Group 2, as a result of vaccinating Group 1 (specifically, up to 11.42% and 12.76% of cumulative new cases and mortality can, respectively, be prevented in Group 2 if individuals in Group 1 are vaccinated at the baseline levels under Scenario 1). Thus, this study shows that, in addition to inducing a moderate reduction in the vaccinated group, the Vaccination Scenario 1 can induce a modest reduction (spillover) of disease burden in the unvaccinated (3 to 5-year old) group in Burkina Faso.

**Figure 11.**
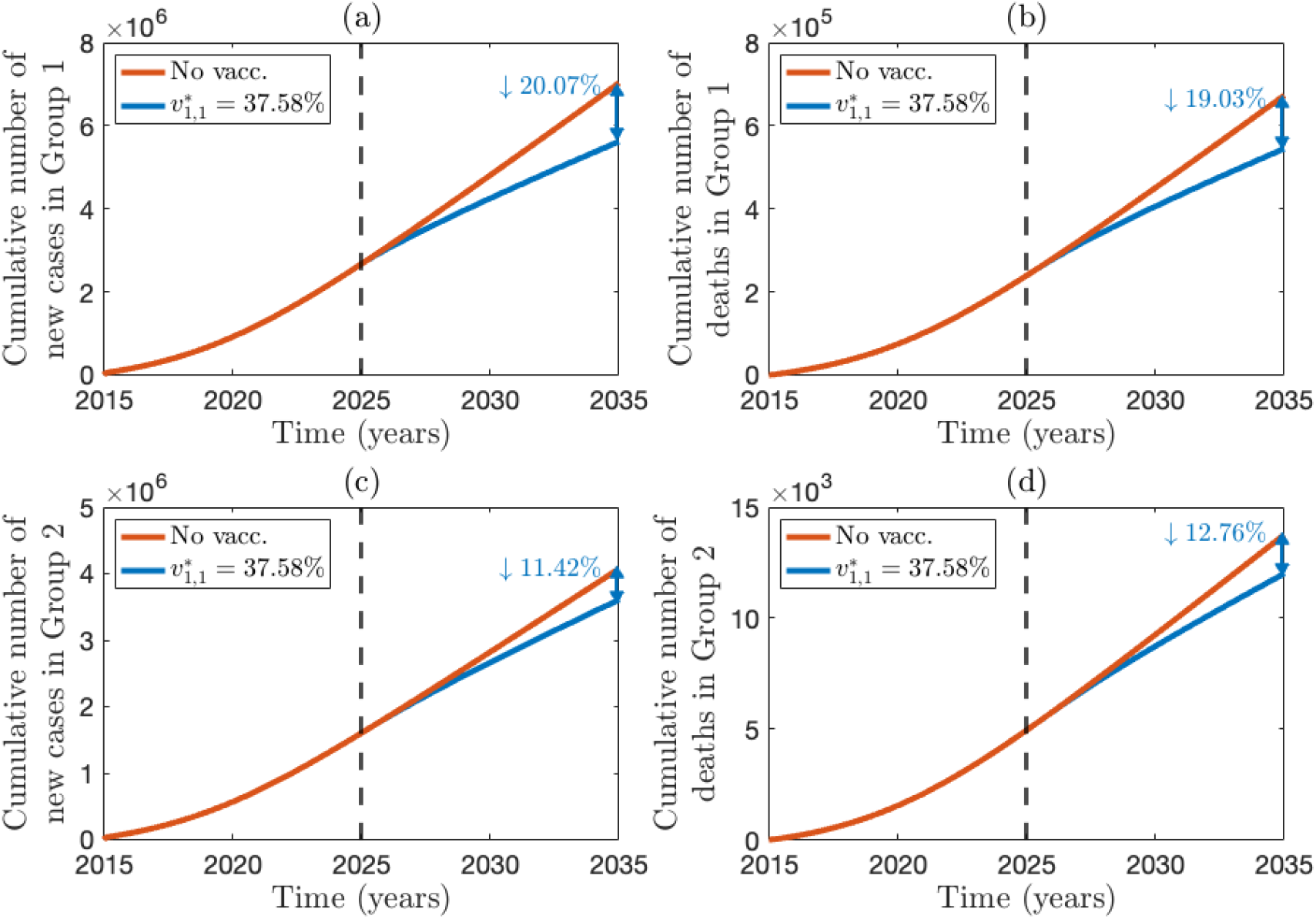
Simulation of the model (G.1) for Vaccination Scenario 1, showing the model prediction for the cumulative number of new cases and disease-induced mortality in the vaccinated (Group 1) and the unvaccinated (Group 2) populations for the period from 2025-2035, as a function of time. Simulations were ran for the pre-vaccination period from 2015-2024 (illustrated to the left of the dashed black vertical line). (a) Cumulative new malaria cases in Group 1, (b) cumulative malaria-induced mortality in Group 1, (c) cumulative new malaria cases in Group 2, and (d) cumulative malaria-induced mortality in Group 2. The blue curve corresponds to the scenario with vaccination of individuals in Group 1 at baseline coverage and efficacy levels (so that 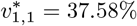), while the red curve represents the vaccination-free (worst-case) scenario (where all vaccination-related parameters, such as the vaccination rate *ξ*_*v*_, are set to zero in the model). All parameters used in the simulations are as given by their baseline in Tables 3 and 4.

### 6.2 Assessing the Impact of Vaccination Scenario 2

The same simulations (as in Scenario 1 above) are carried out to assess the impact of vaccinating Group 1 only, but with all four doses. Here, the simulations are carried out using the model (H.4), given in Appendix H. The results obtained, presented in Figure 12, show a more sizable reduction in cumulative new cases (Figure 12(a); 38.75%) and malaria-induced mortality (Figure 12(b); 36.57%), in comparison to those achieved under the Vaccination Scenario 1 (where the corresponding reductions were only 20.07% and 19.03%, respectively), by the end of the simulation period (2035). Similarly, a more sizable spillover is achieved for the cumulative new cases (Figure 12(c); 25.88%) and malaria-induced mortality (Figure 12(d); 26.92%), in comparison to those recorded under Scenario 1 (where the corresponding reductions were 11.42% and 12.76%, respectively) at the end of the simulation period (2035). Hence, these simulations show that vaccinating Group 1 individuals (only) with all four doses of the vaccine (Vaccination Scenario 2), at baseline coverage levels (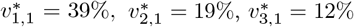, and 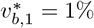), provides stronger direct protection for Group 1 and greater indirect (spillover) protection for Group 2 than the Vaccination Scenario 1.

**Figure 12.**
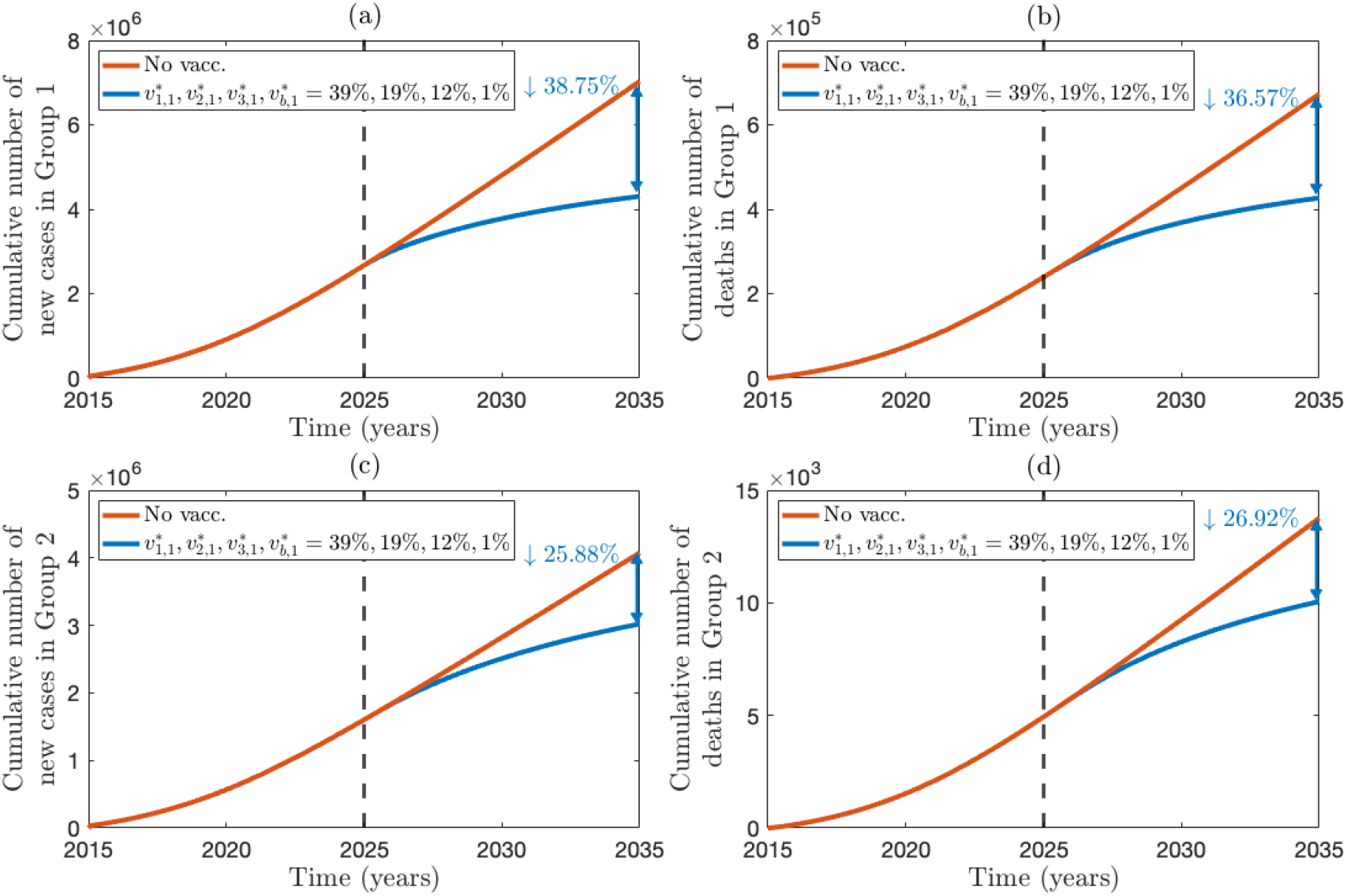
Simulation of the model (H.1)–(H.4) for Vaccination Scenario 2, showing the model prediction for the cumulative number of new cases and disease-induced mortality in the vaccinated (Group 1) and the unvaccinated (Group 2) populations for the period from 2025-2035, as a function of time. Simulations were ran for the pre-vaccination period from 2015-2024 (illustrated to the left of the dashed black vertical line). (a) Cumulative new malaria cases in Group 1, (b) cumulative malaria-induced mortality in Group 1, (c) cumulative new malaria cases in Group 2, and (d) cumulative malaria-induced mortality in Group 2. The blue curve corresponds to the scenario with vaccination of individuals in Group 1 at baseline coverage and efficacy levels (so that 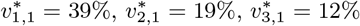, and 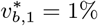), while the red curve represents the vaccination-free (worst-case) scenario (where all vaccination-related parameters, such as the vaccination rate *ξ*_*v*_, are set to zero in the model). All parameters used in the simulations are as given by their baseline in Tables 3 and 4.

### 6.3 Assessing the Impact of Vaccination Scenario 3

Here, the model (I.1) is used to assess the impact of vaccinating both groups, but with only the first dose administered (i.e., Scenario 3). For these simulations (using the baseline values of the parameters in Tables 3 and 4), the steady-state vaccination coverage for Groups 1 and 2 are 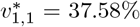 and 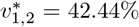, respectively. The results obtained, depicted in Figure 13, show a 26.59% (Figure 13(a)) and 25.07% (Figure 13(b)) reduction in the cumulative new cases and malaria-induced mortality, respectively, in Group 1 (these represent a modest increase in the percent reduction compared to those achieved using the corresponding Vaccination Scenario 1, which achieved 20.07% and 19.03% reductions, respectively), by the end of the simulation period (2035). Similarly, this figure show a significant spillover reduction of the cumulative new cases (25.07%; see Figure 13(c)) and malaria-induced mortality (24.12%; see Figure 13(d)) in Group 2 (these represent a significant increase in the reductions (spillover) recorded using the Vaccination Scenario 1, which stood at 11.42% and 12.76% reductions, respectively) at the end of the simulation period (2035).

**Figure 13.**
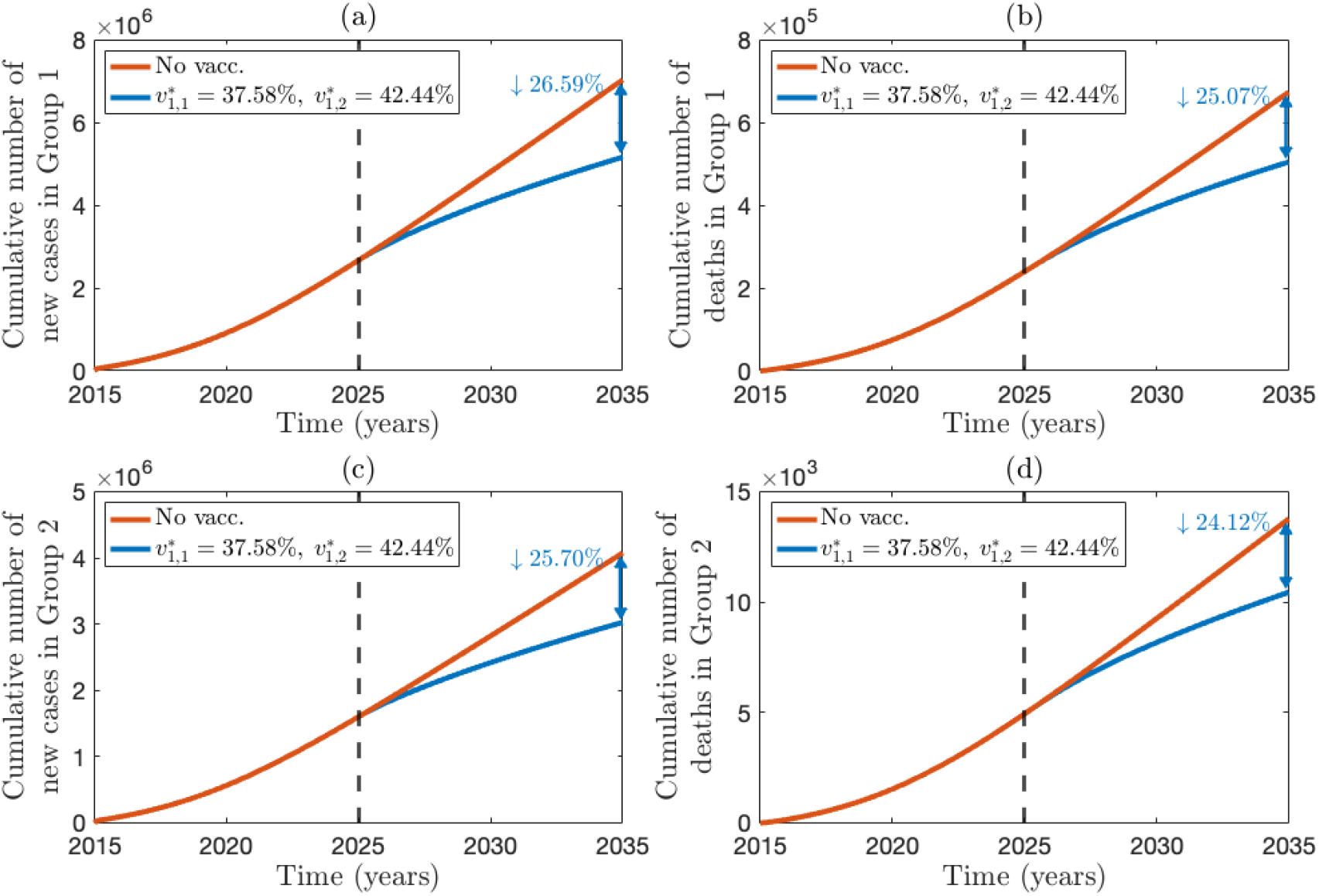
Simulation of the model (I.1) for Vaccination Scenario 3, showing the model prediction for the cumulative number of new cases and disease-induced mortality in the vaccinated (Group 1) and the unvaccinated (Group 2) populations for the period from 2025-2035, as a function of time. Simulations were ran for the pre-vaccination period from 2015-2024 (illustrated to the left of the dashed black vertical line). (a) Cumulative new malaria cases in Group 1, (b) cumulative malaria-induced mortality in Group 1, (c) cumulative new malaria cases in Group 2, and (d) cumulative malaria-induced mortality in Group 2. The blue curve corresponds to the scenario with vaccination of individuals in Group 1 at baseline coverage and efficacy levels (so that 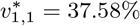 and 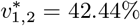), while the red curve represents the vaccination-free (worst-case) scenario (where all vaccination-related parameters, such as the vaccination rate *ξ*_*v*_, are set to zero in the model). All parameters used in the simulations are as given by their baseline in Tables 3 and 4.

### 6.4 Assessing the Impact of Vaccination Scenario 4

In this section, the model (2.2)–(2.6) is simulated to assess the impact of vaccinating both groups with all four doses (for this setting, the steady-state vaccine coverages for the four doses for Group 1 are 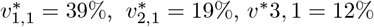, and 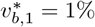,respectively; while those for Group 2 are 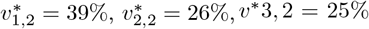, and 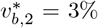, respectively). The results obtained, depicted in Figure 14, show a marked reduction in both the cumulative new cases (Figure 14 (a)) and malaria-induced mortality (Figure 14 (b)) throughout the simulation period. Specifically, it can be seen from Figures 14(a) and 14(b) that up to 46.41% and 43.86% reductions in the cumulative new cases and mortality are recorded at the end of the simulation period, respectively (this represents a significant increase in the reductions of the cumulative new cases and mortality recorded under the corresponding Vaccination Scenario 2, where only individuals in Group 1 are vaccinated with the four doses, which resulted in 38.75% and 36.57% reductions, respectively). Furthermore, the simulations in Figure 14 show a marked reduction in the cumulative new cases (Figure 14(c)) and disease-induced mortality (Figure 14(d)). Specifically, up to 46.51% cumulative new cases and 43.36% malaria-induced mortality can be prevented at the end of the simulation period under this scenario. These simulations show that giving all four doses to both groups (Figure 14) resulted in greater reductions in disease burden (almost twice), compared to Scenario 3 where both groups are vaccinated with only the first doses (i.e., giving four doses, even at baseline level, resulted in much greater reductions in disease burden than giving a single dose). This strategy (Scenario 4) also does much better than Scenario 2, where only individuals in Group 1 are vaccinated but all four doses (compare the corresponding 46.41% and 43.86% reductions achieved under Scenario 4, with the 38.75% and 36.57% reductions achieved under Scenario 2). In other words, additionally vaccinating Group 2 always does better than a strategy that focuses on vaccinating Group 1 only. Thus, this study shows that our proposed extended vaccination strategy (which entails vaccinating both Groups 1 and 2) is far more effective (i.e., reduces more cases and malaria-induced mortality) than a strategy that focuses on vaccinating only individuals in Group 1. In summary, the simulations in Figure 14 show that vaccination (regardless of which group gets vaccinated and how many doses are administered) always reduces disease burden, in comparison to the worst-case (vaccination-free) scenario (compare blue curves with red curves in Figure 11). Furthermore, these reductions increase with increasing number of doses administered to the vaccinated group or groups (compare Figures 11 and 12; and also Figures 13 and 14). Significant spillover is achieved using Vaccination Strategies 1 and 2, and the reductions increase with increasing number of doses administered (compare Figures 11 and 12).

**Figure 14.**
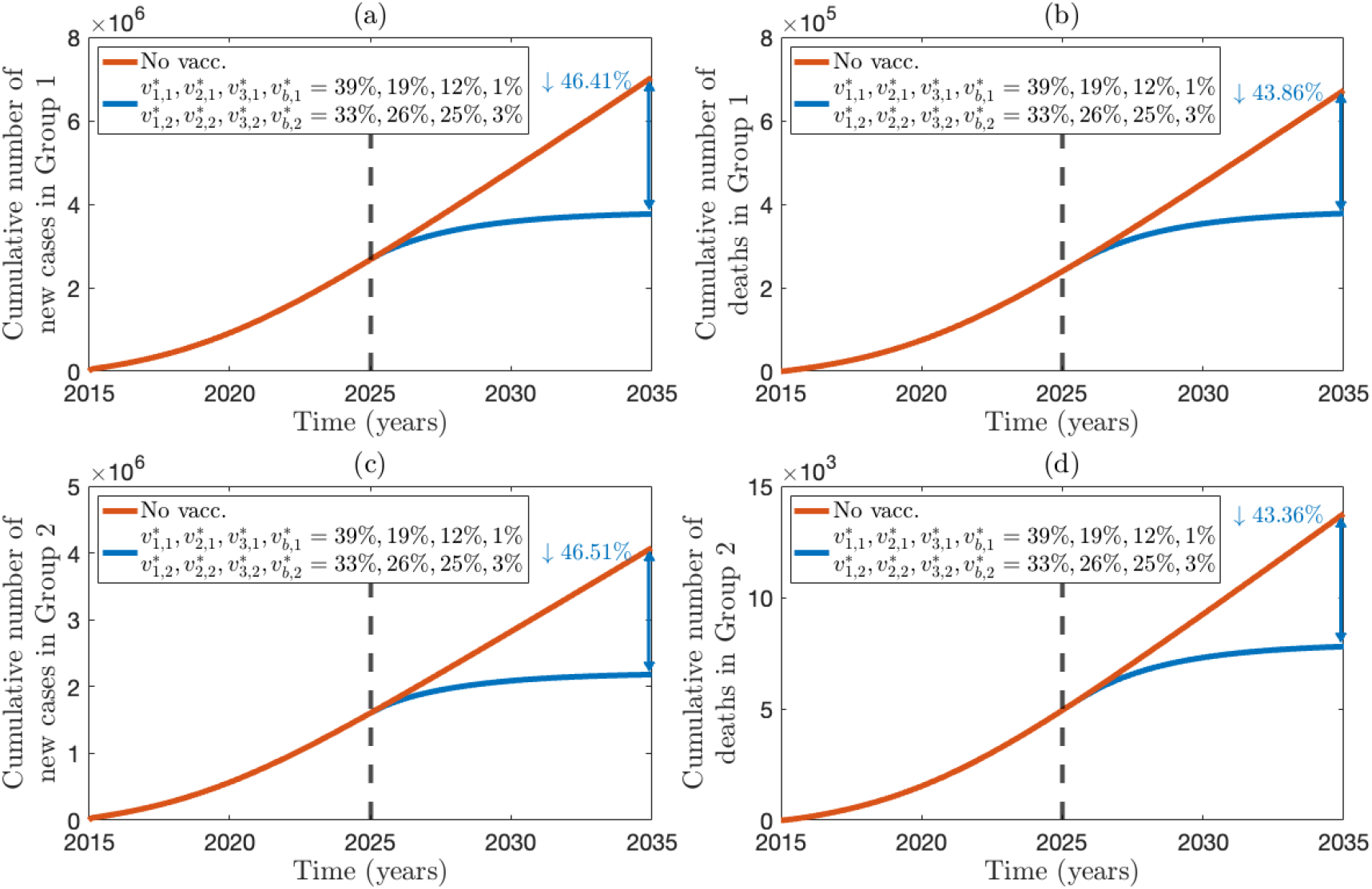
Simulation of the model (2.2)–(2.6) for Vaccination Scenario 4, showing the model prediction for the cumulative number of new cases and disease-induced mortality in the vaccinated (Group 1) and the unvaccinated (Group 2) populations for the period from 2025-2035, as a function of time. Simulations were ran for the pre-vaccination period from 2015-2024 (illustrated to the left of the dashed black vertical line). (a) Cumulative new malaria cases in Group 1, (b) cumulative malaria-induced mortality in Group 1, (c) cumulative new malaria cases in Group 2, and (d) cumulative malaria-induced mortality in Group 2. The blue curve corresponds to the scenario with vaccination of individuals in Group 1 at baseline coverage and efficacy levels (so that 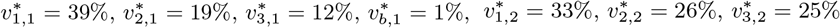, and 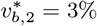), while the red curve represents the vaccination-free (worst-case) scenario (where all vaccination-related parameters, such as the vaccination rate *ξ*_*v*_, are set to zero in the model). All parameters used in the simulations are as given by their baseline in Tables 3 and 4.

## 7 Discussion and Conclusions

Malaria is a life-threatening mosquito-borne disease caused by *Plasmodium* parasites and transmitted to humans *via* the bites of infectious female *Anopheles* mosquitoes, and it remains a major global public health burden, causing an estimated 282 million cases and 610,000 deaths in 2024 [1]. The burden is borne disproportionately by sub-Saharan African countries, where the deadliest parasite, *P. falciparum* is prevalent [2]; in particular, young children under five years of age bear a disproportionate share of malaria morbidity and mortality [2]. In moderate-to high-endemic settings such as Burkina Faso, malaria transmission persists despite the implementation of established control measures, including insecticide-treated nets (ITNs), indoor residual spraying (IRS), and artemisinin-based combination therapy, underscoring the need to assess the population-level impact of the new vaccines recommended by the World Health Organization (WHO) against *P. falciparum* malaria in young children, namely RTS,S/AS01 (developed by GlaxoSmithkline, and deployed since 2023 [31,32,35]) and R21/Matrix-M (developed by the Jenner Institute, University of Oxford in collaboration with the Serum Institute of India Private Limited (SIIPL), and rolled out in several sub-Saharan African countries, including Burkina Faso, during the 2024 [33–35]). The WHO recommends these vaccines to be administered to children starting from the age of 5 months to 3 years of age (although the age range can be extended to children under five years of age in endemic areas if needed [33, 37, 52]), who bear the aforementioned disproportionate burden (e.g., they account for 75% of malaria mortality [2]). Specifically, In Burkina Faso, the ongoing malaria vaccination program targets children in the 0–3 year age group, where the first three primary doses are administered 28 days (four weeks) apart, and a booster dose is given one year after receiving the third dose. The purpose of the current study was to use a mathematical modeling framework, backed by empirical data, to assess the population-level impact of the most recent malaria vaccine, R21/Matrix-M, as the main intervention, in conjunction with insecticide-treated bednet use, on malaria transmission dynamics and control in Burkina Faso (one of the main sites for the clinical trials of the R21/Matrix-M vaccine in sub-Saharan Africa [35–37]). To achieve the objectives of this study, we developed, calibrated, and simulated a new mathematical modeling framework for assessing the population impact of vaccination in reducing malaria burden in children in Burkina Faso. Although malaria vaccination is currently recommended for children in the 0–3 year age group in Burkina Faso, the modeling framework we proposed also considered the older 3–5-year age group (thereby enabling us to explore the hypothetical scenario of whether or not the overall effectiveness of the current vaccination program can be enhanced by also vaccinating the older age group). Consequently, the model we proposed, which was based on stratifying the total population of children in the 0 to 5 years age bracket in Burkina Faso into the 0–3-year age group (Group 1) and the 3–5-year age group (Group 2) and also incorporate bednet usage, takes the form of a deterministic system of age- and dose-structured delay differential equations.

To estimate some of the unknown parameters of the model, the corresponding vaccination-free model was fitted to observed yearly cumulative malaria mortality data for children under five years of age in Burkina Faso, for the period 1980–2021. The disease-free equilibrium of the full (vaccination) model was shown to be locally-asymptotically stable whenever an epidemiological threshold quantity, known as the control reproduction number (ℛ_*v*_), is less than one. The epidemiological implication of this result is that malaria can be eliminated from the under-five population in Burkina Faso if the vaccination program (complemented with the bednet usage) can reduce (and maintain) the control reproduction number to a value less than one, provided that the initial number of infected individuals is sufficiently small. The global sensitivity analysis, using Latin Hypercube Sampling and Partial Rank Correlation approach, revealed that parameters related to effective malaria transmission from humans to mosquitoes and mosquitoes to humans (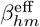 and 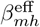), recovery rate (*γ*_*h*_), the natural death rate of adult female mosquitoes (*µ*_*m*_), and overall bednet effectiveness (bed^eff^) have the highest impact on the value of the control reproduction number, ℛ_*v*_ (the standard metric used to quantify disease burden in the community). Thus, this study showed that by targeting these parameters (which greatly influence disease burden), appropriately (i.e., by using the appropriate interventions that target these parameters), malaria can be effectively controlled or eliminated in the targeted population of the 0–5-year age group in Burkina Faso. This result is consistent with those reported in [72] for a vaccination-free malaria model in a high transmission setting such as Nigeria in West Africa, and it also supports the findings in [75], where a vaccination-free malaria model was considered for KwaZulu-Natal, a province in South Africa.

For the case where the total population consists only of children under three years of age (i.e., individuals in Group 1; and Group 2 are excluded) and they received only the first dose of the vaccine, the simulations showed that, for this case where bednets at baseline level are used in the community, vaccine-induced herd immunity can be achieved (i.e., the disease will be eliminated in the population) if almost all (at least 96.53%) of these children are vaccinated with the first dose (such herd immunity cannot be achieved in the absence of bednet use in the community). If these individuals received all four doses (and bednets are not used in the community), the herd immunity threshold (HIT) can be achieved if the ratios of individuals who received doses 2, 3, and the booster dose range between 73% and 90% (which is generally attainable in Burkina Faso). These percentages dramatically decrease to 10% to 30% if bednets are used in the community. Thus, the simulations showed that having individuals in Group 1 receive all four doses is more effective in achieving herd immunity than receiving only the first dose (as expected), and that the effectiveness of the vaccination program to achieve herd immunity is greatly enhanced by complementing it with bednet usage (even at the baseline level).

Simulations for the hypothetical case where the total population now consists of individuals in Groups 1 and 2 (i.e., children of ages 0 to 5 in Burkina Faso), but only individuals in Group 1 are vaccinated (and they received only the first dose) and bednets are used at baseline level (Vaccination Scenario 1 in our study), showed, in addition to significant reduction of disease burden (in terms of cumulative new cases and disease-induced mortality) in Group 1 (20.07% and 19.03%, respectively), a moderate spillover reduction of cumulative new cases (11.42%) and mortality (12.76%) in the unvaccinated Group 2, in comparison to the worst-case (vaccination-free) scenario. Thus, vaccinating one group resulted in a reduction of malaria burden in the other (unvaccinated) group. In comparison to the worst case scenario, these reductions are more pronounced in the vaccinated (38.75% and 36.57%, respectively) and unvaccinated (25.88% and 26.92%, respectively) groups when individuals in Group 1 received all four doses (Vaccination Scenario 2 in this study).

We simulated the potential impact of our proposed extended vaccination strategy for Burkina Faso, where individuals in the 3 to 5-year age group (i.e., individuals in Group 2) are also vaccinated (recall that the current malaria vaccination strategy in Burkina Faso is to vaccinate individuals in the 0 to 3-year age group (i.e., Group 1) only), and both groups received only the first dose of the vaccine and bednets are used at baseline level (this strategy is termed Vaccination Scenario 3 in our study). Our simulation results showed, in comparison to the worst-case scenario, moderate reductions in the cumulative number of new malaria cases and malaria-induced mortality in both groups (specifically, up to 26.59% and 25.70% reduction in the cumulative number of new cases in Group 1 and Group 2, respectively, were recorded. Similarly, this resulted in a 25.07% and 24.12% reduction in the cumulative mortality in Group 1 and Group 2, respectively). Furthermore, this proposed extended vaccination strategy reduces the vaccination coverage required to achieve HIT in both groups (i.e., in the entire 0 to 5-year age group). In particular, for this scenario, our results showed that HIT can be achieved in the total population (of children within the 0 to 5-year age bracket) by vaccinating 46% of the total population of 0 to 5-year old in Burkina Faso (which is also equivalent to vaccinating 75% of the total population of children in the 0 to 3-year age group and not vaccinating anyone in the 3 to 5-year age group). This HIT requirement (of either vaccinating 46% of Groups 1 and 2 or 75% of Group 1) is equivalent to vaccinating about 1.7 million children in the 0 to 5-year age group in Burkina Faso. It should be recalled, for the case where the total population consists of 0 to 3-year old children and they also received the first dose only, the results obtained from simulating this homogeneous (Group 1-only) model showed that HIT can only be achieved in this population if at least 96.53% of them are vaccinated with the first dose (this corresponds to vaccinating approximately 2.1 million children in the 0–3 year age group in Burkina Faso). Thus, vaccinating the broader population of Groups 1 and 2 can achieve herd immunity using fewer vaccine resources (1.7 million *vs*. 2.1 million doses) than a strategy that targets only Group 1, suggesting that the current malaria vaccination strategy in Burkina Faso may be suboptimal, compared to the proposed extended malaria vaccination program (for the special case where only the first dose is administered; it should be noted that the current malaria vaccination program in Burkina Faso administers all four doses to Group 1, and not just the first dose only). In summary, this study showed that the vaccination coverage level needed to achieve HIT is lower for the heterogeneous (two-group) than for the homogeneous (one-group) model. This finding is consistent with those reported in [101–103], which also demonstrated that HIT levels for heterogeneous models can be lower than those for their corresponding homogeneous models.

Higher reductions in the cumulative number of new malaria cases and malaria-induced mortality were recorded for the case where both groups are vaccinated and they received all four doses, as against the first dose only scenario discussed above. In this case (i.e., Vaccination Scenario 4 in our study), up to 46.41% and 46.51% reductions in cumulative new cases (in comparison to the worst-case) scenario were achieved in Group 1 and Group 2, respectively. Similarly, up to 43.86% and 43.36% reductions in Group 1 and Group 2, respectively, were achieved, in comparison to the worst-case scenario. Thus, the simulations suggested that extending vaccination to include both groups consistently outperforms a strategy that targets only the younger group (Group 1), and that completing the full four-dose vaccination schedule provides substantially greater direct and spillover benefits than administering the first dose only, thereby supporting the proposed extended vaccination strategy for reducing malaria burden among children under five years of age in Burkina Faso. In other words, our simulation results showed that extending the malaria vaccination program to include both groups (Vaccination Scenarios 3 and 4) consistently outperforms a strategy that targets only individuals in Group 1 (Vaccination Scenarios 1 and 2), and that completing the full four-dose vaccination schedule provides substantially greater direct benefits (compare Vaccination Scenarios 1 and 2; and also Vaccination Scenarios 3 and 4) and greater spillover benefits (compare Vaccination Scenarios 1 and 2) than administering the first dose only. In summary, the proposed extended vaccination strategy (i.e., vaccinating both groups 1 and 2) offers improved prospects for enhancing malaria vaccination effectiveness in Burkina Faso, compared to their current malaria vaccination program (which targets vaccinating individuals in Group 1 only).

Some limitations of the model developed in this study include the exclusion of older age groups (i.e., individuals older than five years of age) and not accounting for malaria transmission by asymptomatic infected individuals. The authors hope to address these limitations in a future study. It is worth noting that the modeling framework presented in this study, for malaria transmission dynamics in Burkina Faso, can easily be adapted and applied to other malaria-endemic countries, regions, or jurisdictions interested in assessing the population-level impact of vaccination, together with traditional insecticide-based vector control interventions (such as the use of bednets or indoor residual spraying). Overall, this study showed that malaria elimination among children in Burkina Faso in the 0–3 year age group is feasible by either administering the first dose of the R21/Matrix-M vaccine at sufficiently high coverage or administering all three primary doses together with the booster dose at moderate coverage, provided that bednets are used at baseline levels. The results further showed that extending the current malaria vaccination program in Burkina Faso to also include older children in the 3–5-year age bracket strongly enhances the prospect of malaria elimination in the high-risk (children in the 0–5-year age bracket) population in Burkina Faso, even when only the first dose of the vaccine is administered. Furthermore, our proposed extended vaccination strategy generally requires fewer vaccine resources, compared to the current vaccination program, to achieve herd immunity in Burkina Faso.

## Data Availability

All data used in the manuscript are publicly available

## Acknowledgments

ABG acknowledges the support, in part, of the National Science Foundation (Grant Number: DMS-2052363; transferred to DMS-2330801).

## A Proof of Theorem 2.1

*Proof*. The general idea of the proof is to show that to each initial data or history of the state variables of the delay differential equation (DDE) system (2.2)–(2.6), there exists a solution for the system, and also show that if there are any two arbitrary solutions, *X*_1_(*t*) and *X*_2_(*t*) for the DDE system with the same history function *ϕ* on [−*τ*, 0], then *X*_1_(*t*) = *X*_2_(*t*) for all *t* in their common interval of existence. The proof is based on using the properties of retarded (time-delay) functional differential equation (RFDE), as described in [108] (and also used in [109]). Recall the map

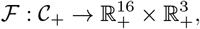

defined by the right-hand sides of the model system (2.2)–(2.6), where 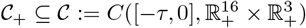. The model can then be re-written as:

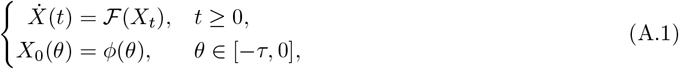

where *X*(*t*) is the vector of the state variables of the model, *X*_*t*_ ∈ 𝒞_+_ is defined, for each *t* ≥ 0, as *X*_*t*_(*θ*) = *X*(*t* + *θ*) for *θ* ∈ [−*τ*, 0], and *ϕ* is the initial history function of the state variables of the model, as defined in Section 2. Hence, to prove Theorem 2.1, it suffices to show that ℱ is continuous and locally Lipschitz on the Banach space 𝒞_+_.

### Continuity of ℱ

Let Φ ∈ 𝒞_+_ be any arbitrary history function for the state variables of the model. It follows then that each component of ℱ (Φ) in the model system (2.2)–(2.6) (or, equivalently, (A.1)) is a finite sum of the following terms: (i) linear terms in Φ(0) and Φ(−*τ*_*l*_) (where *τ*_*l*_ = *τ*_12_, *τ*_23_, *τ*_3*b*_, *τ*_12_ + *τ*_23_, *τ*_23_ + *τ*_3*b*_, *τ*_12_ + *τ*_23_ + *τ*_3*b*_), (ii) force of infection terms (which are well-defined on 𝒞_+_, since *N*_*h*_(Φ(0)) > 0 and *N*_*m*_(Φ(0)) > 0 on 𝒞_+_), and (iii) survival terms *J*_*fi*_(Φ), with *f* = 1, 2, 3, and *i* = 1, 2, given by exponential of integrals of bounded linear functions of Φ over intervals of length at most *τ* (see (2.3) and (2.5)). Each of these terms (or operations) is continuous with respect to Φ in the supremum norm || · || on 𝒞_+_. Hence, since the finite sum of continuous functions is also continuous, the vector field ℱ is continuous on 𝒞_+_.

### Lipschitzian property of ℱ

We now show that ℱ is locally Lipschitz in a neighborhood of every 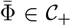. Consider any arbitrary 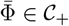, and define

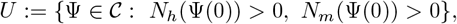

where 𝒞 is the Banach space defined in Section 2. Since the maps Ψ ↦*N*_*h*_(Ψ(0)) and Ψ ↦ *N*_*m*_(Ψ(0)) are continuous on 𝒞, the set *U* is open in 𝒞. Furthermore, owing to the fact that 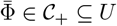, there exists *ρ*_0_ > 0 such that

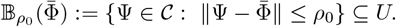

To control the denominators in the force of infection terms, we first establish a uniform positive lower bound for the total human population (*N*_*h*_(Ψ(0))) in a neighborhood of 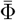 as follows:

#### Lemma A.1

(Uniform positivity of *N*_*h*_(Ψ(0)) near 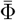). *Let* 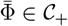. *Then there exist constants ρ*_1_ > 0, *and K*_*h*_ > 0 *such that for every* Ψ ∈ 𝒞 *with* 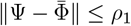,

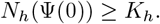

*In particular, all terms in* ℱ *that contain N*_*h*_(Ψ(0)) *in the denominator are well defined on* 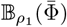.

*Proof*. Since 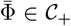, it follows that 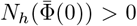. Consider the map *G*_*h*_: 𝒞_+_ → ℝ, defined by *G*_*h*_(Ψ) = *N*_*h*_(Ψ(0)). The evaluation map Ψ → Ψ(0) is continuous on 𝒞_+_ and the total human population *N*_*h*_ is linear in the 16 human state variables of the model. Hence, *G*_*h*_ is continuous at 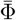, and 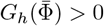 (since 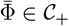). It follows, by the continuity of *G*_*h*_ at 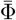, that there exists a *δ* > 0 such that whenever 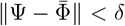, then 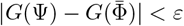, for some *ε* > 0. Choose, without loss of generality, 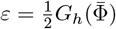. Then,

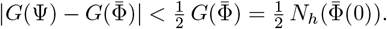

Let 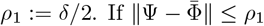, then 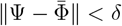, and

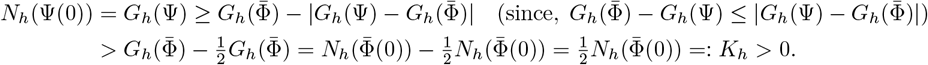

This completes the proof.

Consider, *ρ*:= min{*ρ*_0_, *ρ*_1_}. Then, for all 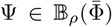, we have Ψ ∈ *U* and the uniform lower bounds *N*_*h*_(Ψ(0)) ≥ *K*_*h*_ > 0. In particular, all force of infection terms in ℱ are well-defined on 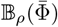.

Now let 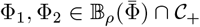 be arbitrary history functions of the state variables. Specifically (for *i* = 1, 2 and *θ* ∈ [−*τ*, 0]),

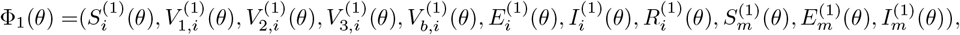

and,

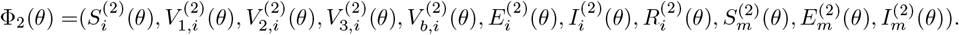

Let,

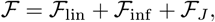

where ℱ_lin_, ℱ_inf_, and ℱ_*J*_ are the 19-dimensional vectors of the linear terms (with and without time-delay), force of infection terms, and the survival probability terms (*J*_*fi*_, with *f* = 1, 2, 3 and *i* = 1, 2), respectively, in the model (2.2)–(2.6). We now show that ℱ_lin_, ℱ_inf_, and ℱ_*J*_ are locally Lipschitz on 𝒞_+_.

### Linear terms (ℱ_lin_)

Since ℱ_lin_ is affine in Φ(0) and in finitely many delayed values Φ(−*τ*_ℓ_), there exists *L*_lin_ > 0 such that

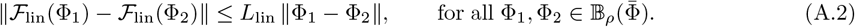

One can take *L*_lin_ as the maximum of the absolute values of the coefficients of the linear terms in (2.2)–(2.6). For example, a valid choice is (where all linear terms with negative coefficients now become positive)

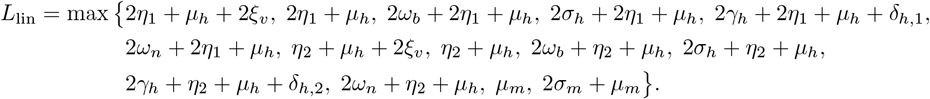

### Force of infection terms (ℱ_inf_)

For *i* = 1, 2, set 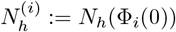. Also define the uniform bounds

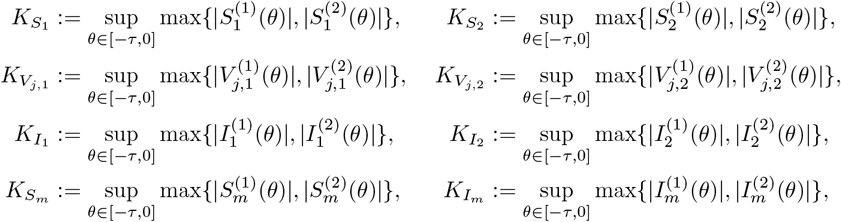

for *j* = 1, 2, 3, *b*. Since 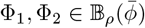, all these uniform bounds are finite. Furthermore, by the definition of the norm, for any component *Y* of the state vector,

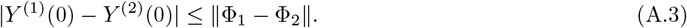

Also, since *N*_*h*_ is a sum of nonnegative human state variables of the model, then:

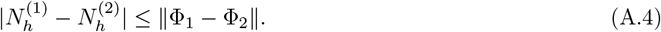

It is convenient to define,

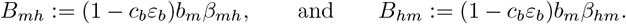

#### Bound for force of infection λ_mh_

Recall 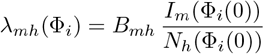 (for *i* = 1, 2). Then,

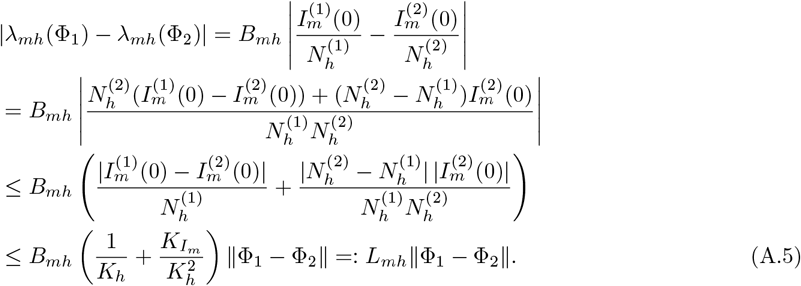

#### Bounds for λ_mh_S_1_ and λ_mh_S_2_

Using the definition of *λ*_*mh*_(Φ_*i*_) (for *i* = 1, 2), it follows that

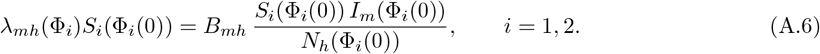

Using (A.6) together with (A.3) and (A.4) gives

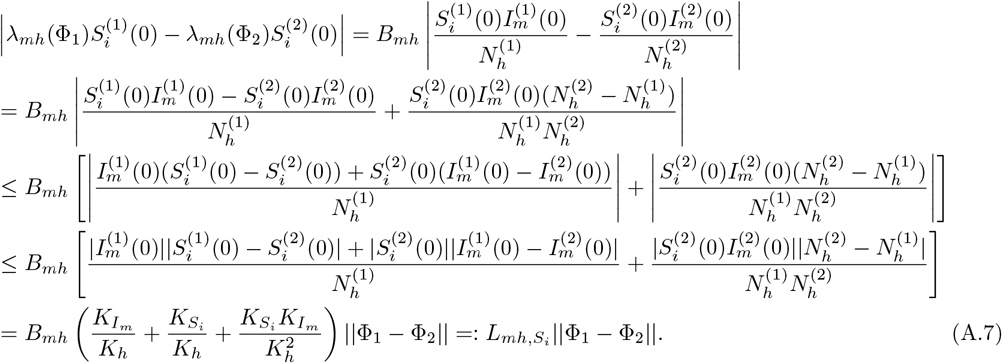

#### Bound for λ_mh_V_j,i_

Similarly, for *j* = 1, 2, 3, *b* and *i* = 1, 2,

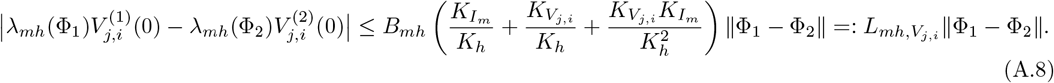

#### Bound for λ_hm_S_m_

Recall 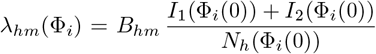 (for *i* = 1, 2). By using (A.3) and (A.4) gives

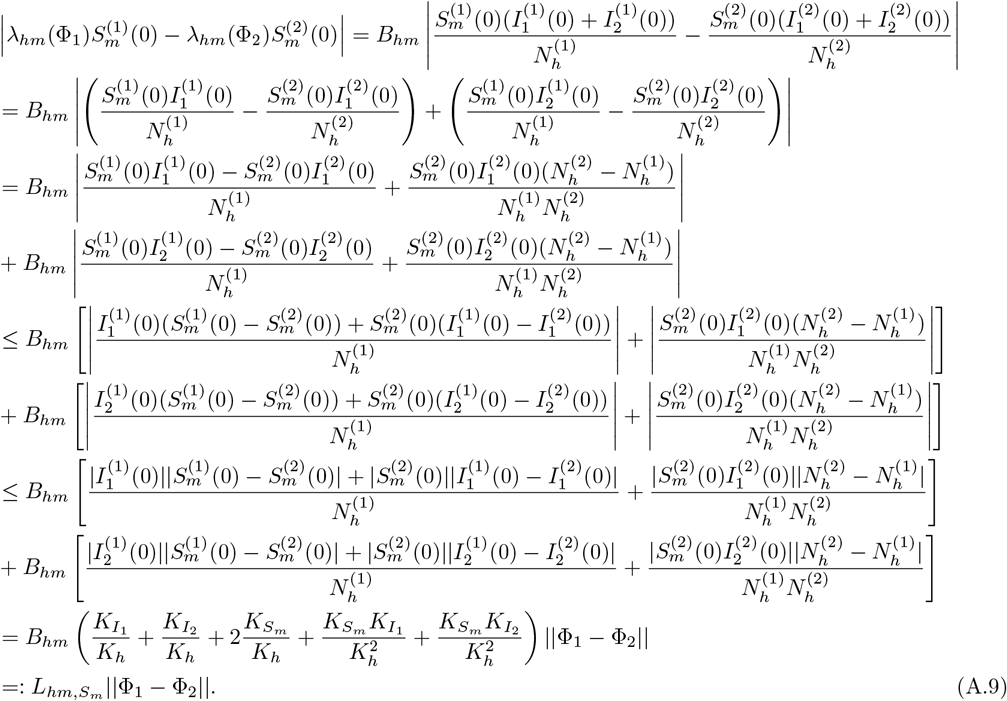

Since every component of ℱ_inf_ is a finite linear combination of the above types of force of infection terms, it follows, from (A.5) and (A.7)–(A.9), that there exists *L*_inf_ > 0 such that

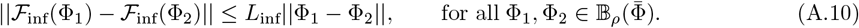

One may take *L*_inf_ to be the maximum of the finitely many constants that multiply the above bounds in the right hand sides of (2.2)–(2.6), including the factors 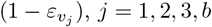. For instance,

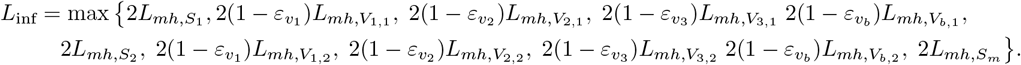

### Survival terms (ℱ_*J*_)

Consider *J*_11_ for illustration. Write

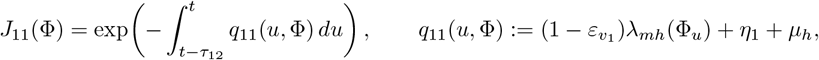

and set 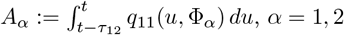. Using the bound for *λ*_*mh*_ gives

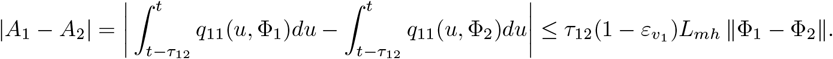

By the mean value theorem applied to *e*^−•^, there exists *ζ* between *A*_1_ and *A*_2_ such that

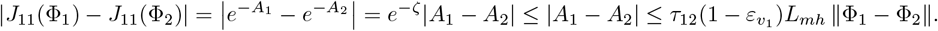

Hence *J*_11_ is locally Lipschitz on 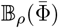 with Lipschitz constant 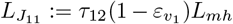. The same argument applies to *J*_21_, *J*_31_ and to *J*_12_, *J*_22_, *J*_32_, giving Lipschitz constants 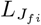 for *f* = 1, 2, 3 and *i* = 1, 2. Furthermore, 0 < *J*_*fi*_(Φ) ≤ 1 for all Φ ∈ 𝒞_+_. Thus, any finite product of the survival terms that appears in ℱ_*J*_ is locally Lipschitz on 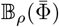. Consequently, there exists *L*_*J*_ > 0 such that

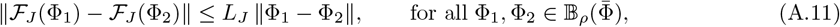

where one can choose,

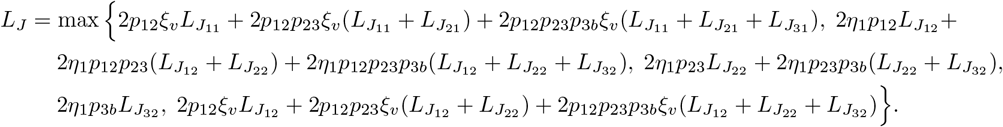

Combining (A.2), (A.10), and (A.11) gives

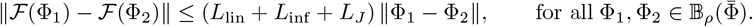

Thus, ℱ is locally Lipschitz on 𝒞_+_. Hence, by the local existence and uniqueness theorem for RFDEs (see [108, Theorem 2.2]), for each initial history function *ϕ* ∈ 𝒞_+_, there exists a *δ* > 0 and a unique solution *X*(*t*) of (2.11), or equivalently (A.1), defined on [−*τ, δ*). This concludes the proof.

## B Positivity of the expressions for *D*_1_ and *D*_2_ in Section 3

Here, we expand the expressions for *D*_1_ and *D*_2_, given in Section 3, and show that they are positive. In particular,

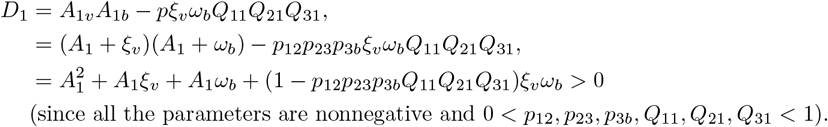

Similarly,

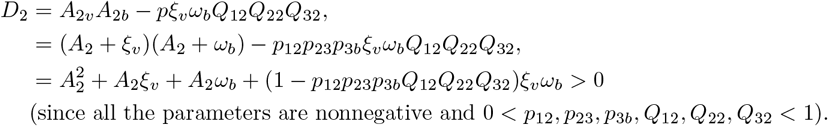

## C Control and Basic Reproduction Numbers of the Model Under Various Vaccination and Dosing Scenarios

In this section, the closed-form expressions for the basic and the control reproduction numbers of the full two-group model (2.2)–(2.6), together with those of its special cases, are listed bellow.

### Scenario 1

The Group 1 only model (E.1) (i.e., with Group 2 excluded), given in Appendix E, where individuals in Group 1 receive only the first dose. For this model the control reproduction number (denoted by 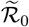) is given by:

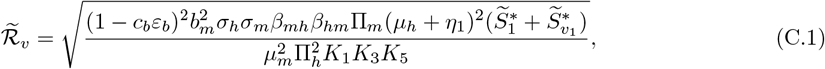

and the basic reproduction number (denoted by 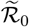) is given by:

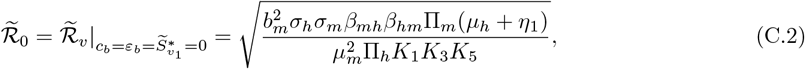

with 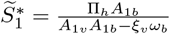, and 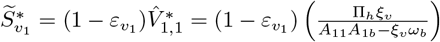, where *A*_1*b*_, and *A*_1*v*_ are defined in Subsection 3.1.

### Scenario 2

The Group 1 only model (F.1)–(F.3) (i.e., with Group 2 excluded), given in Appendix F, where individuals in Group 1 receive all four doses. The control reproduction number for this model, denoted by 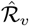, is given by

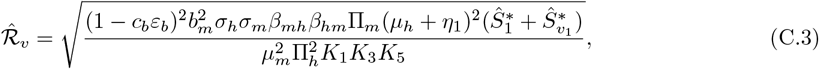

and the corresponding basic reproduction number (denoted by 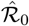) is given by:

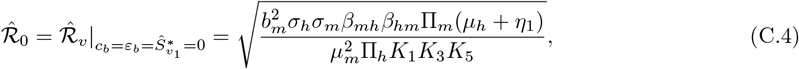

with 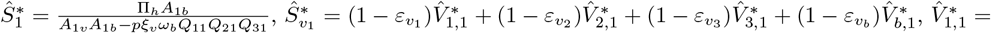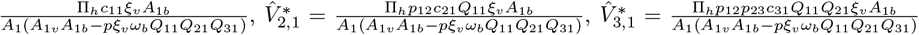, and 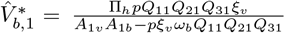, where *A*_1*b*_, *A*_1*v*_, *p, Q*_11_, *Q*_21_, *Q*_31_, *c*_11_, *c*_21_, *c*_31_, and *A*_1_ are defined in Section 3.

### Scenario 3

The model (G.1), given in Appendix G, where only Group 1 is vaccinated (i.e., individuals in Group 2 are not vaccinated), and all vaccinated individuals received only the first dose. For this setting, the control reproduction number, denoted by 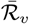, is given by:

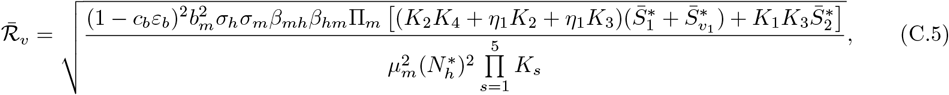

and the basic reproduction number (denoted by 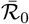) is given by:

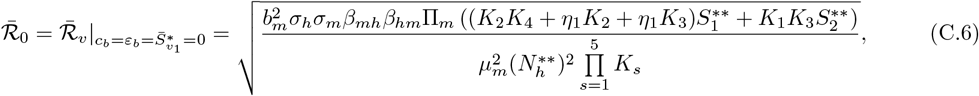

with 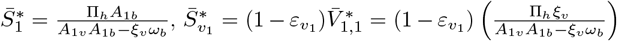, and 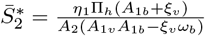, where 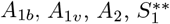, and 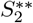 are defined in Section 3.

### Scenario 4

The model (H.1)–(H.4), given in Appendix H, where only Group 1 is vaccinated (i.e., individuals in Group 2 are not vaccinated), and all vaccinated individuals received all four doses. The associated control reproduction number, denoted by 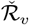, is given by:

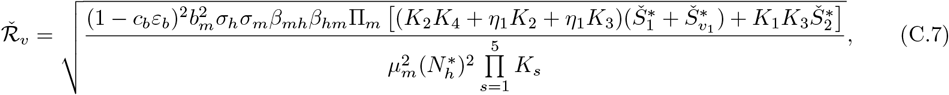

and the associated basic reproduction number, denoted by 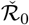, is given by:

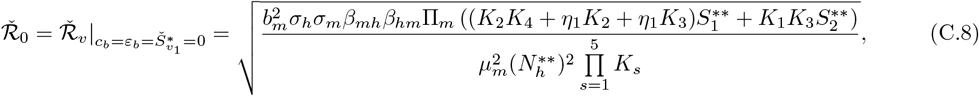

with 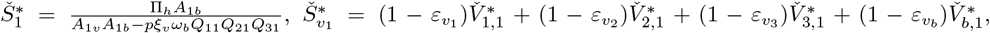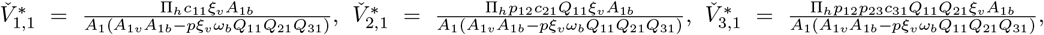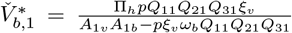, and 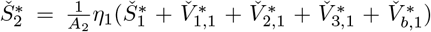, where 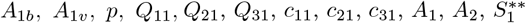, and 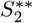 are defined in Section 3.

### Scenario 5

The model (I.1), given in Appendix I, where both Groups 1 and 2 are vaccinated, and all vaccinated individuals receive only the first dose. The control reproduction number for this model (denoted by 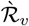) is given by:

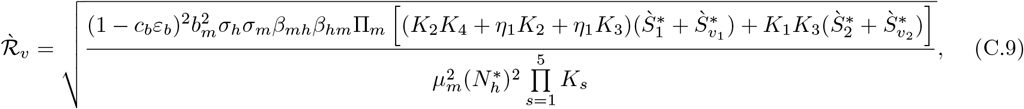

and the corresponding basic reproduction number, denoted by 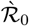, is given by:

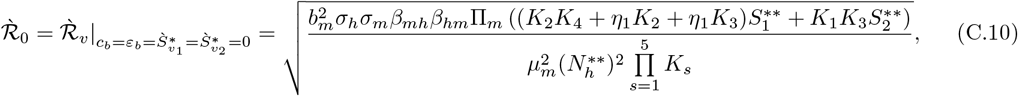

with 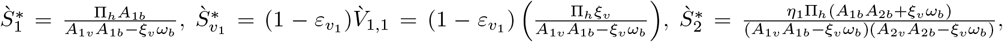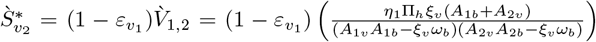, where 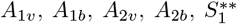, and 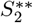 are defined in Subsection 3.1.

### Scenario 6

The model (2.2)–(2.6), given in Section 2, where both Groups 1 and 2 are vaccinated, and all vaccinated individuals receive all four doses. The control reproduction number for this model (under this setting), denoted by ℛ_*v*_, is given by:

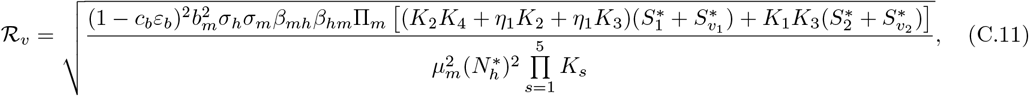

and the corresponding basic reproduction number (denoted by ℛ_0_) is given by:

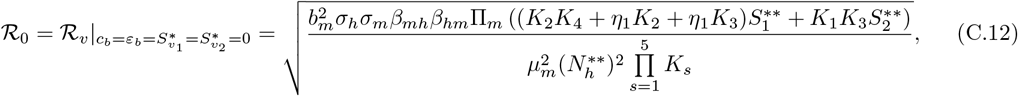

where *K*_*s*_ (with *s* = 1, …, 5), 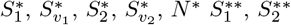, and *N*\*\* are defined in Subsection 3.1.

## D Equations of the Vaccination-free Version of the Model

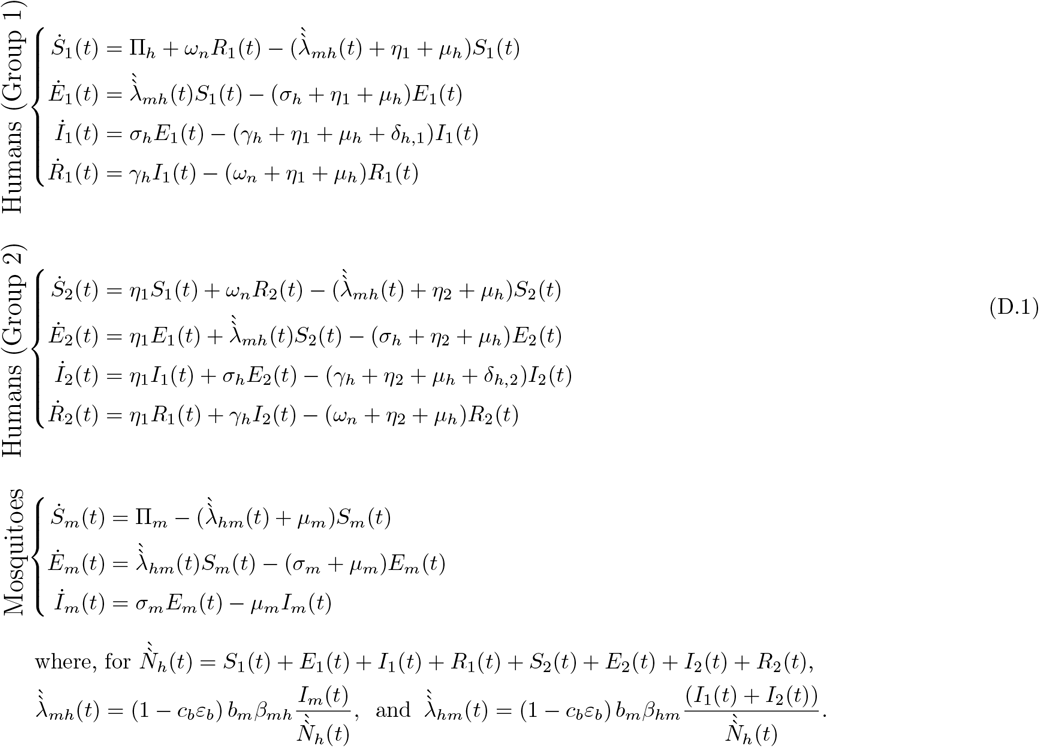

## E Equations for the Special Case of the Model: First Dose Vaccination in Group 1 (Group 2 Excluded)

For this special case, we assume that vaccine-induced immunity from the first dose wanes at the same rate, *ω*_*b*_, as that of the booster dose.

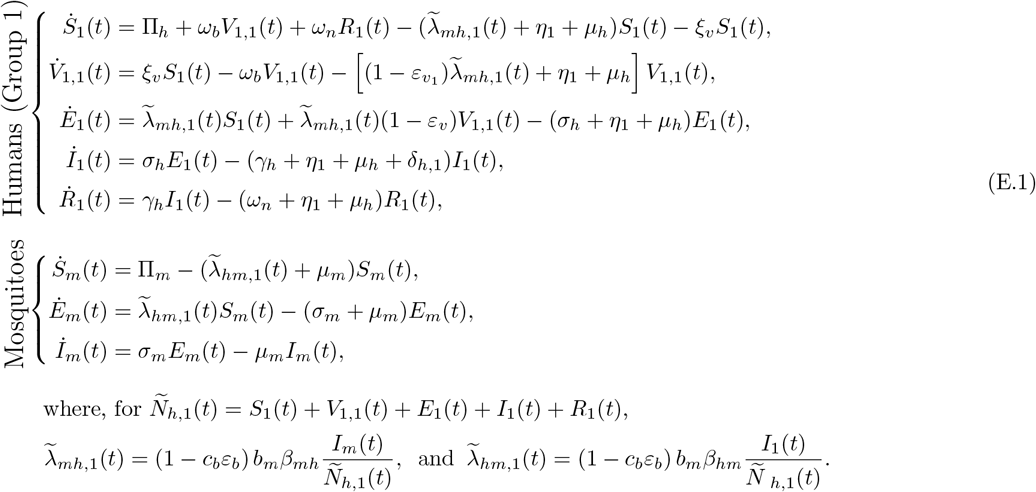

## F Equations for the Special Case of the Model: Four Dose Vaccination in Group 1 (Group 2 Excluded)

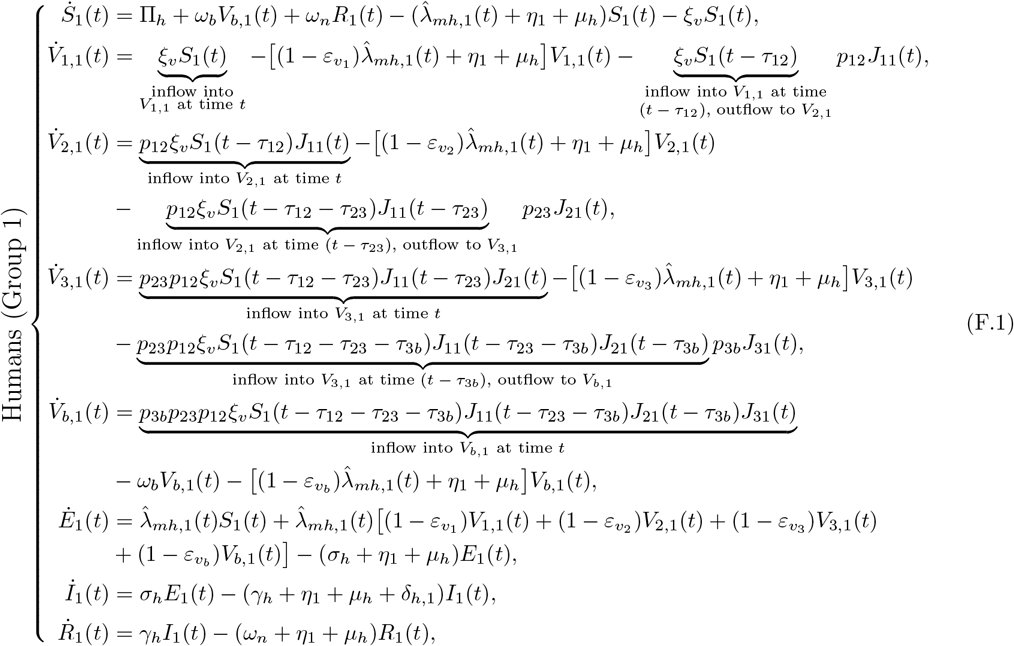

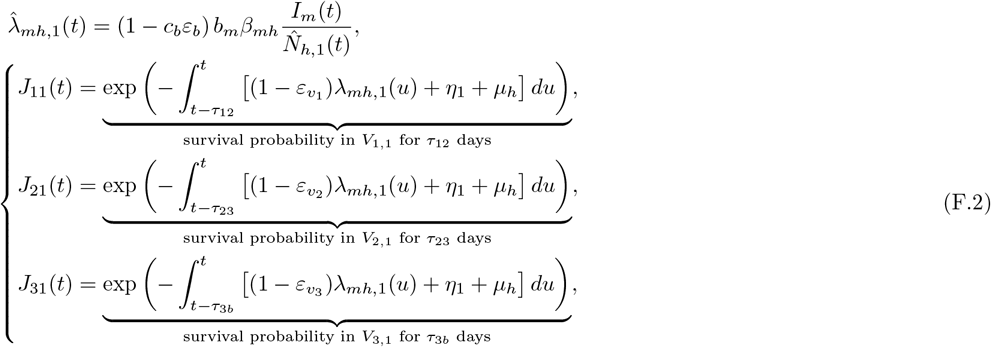

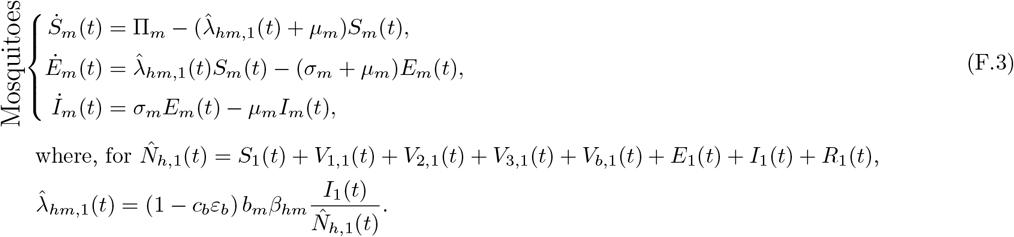

## G Equations for the Special Case of the Model: First Dose Vaccination for Group 1 (No Vaccination for Group 2)

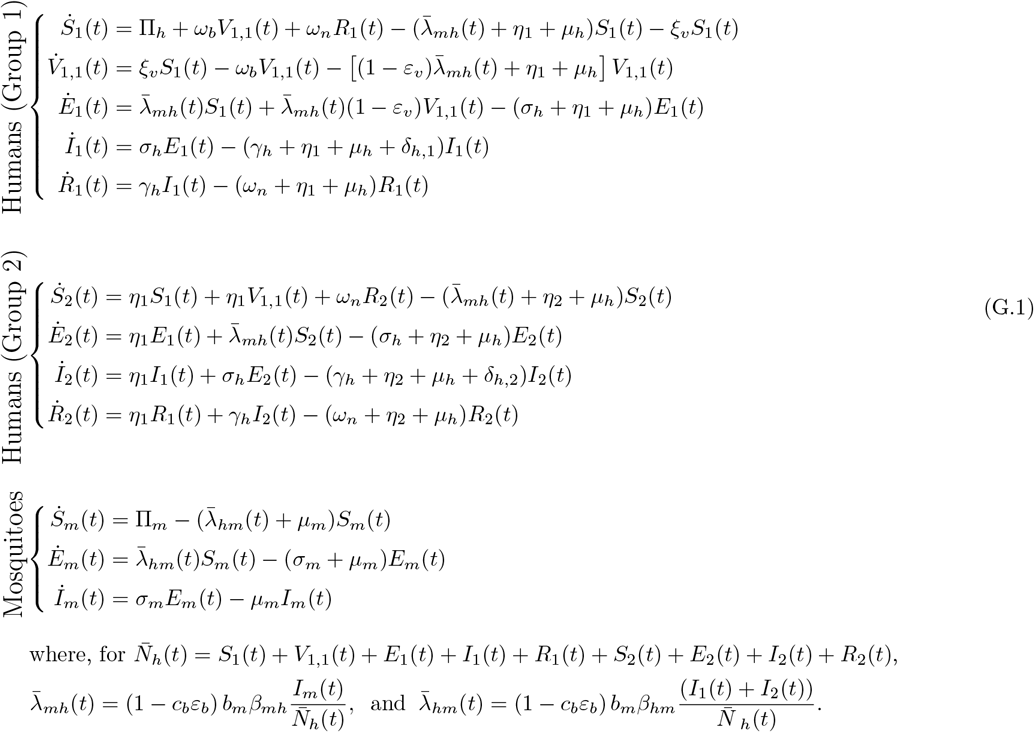

## H Equations for the Special Case of the Model: Four Dose Vaccination for Group 1 (No Vaccination for Group 2)

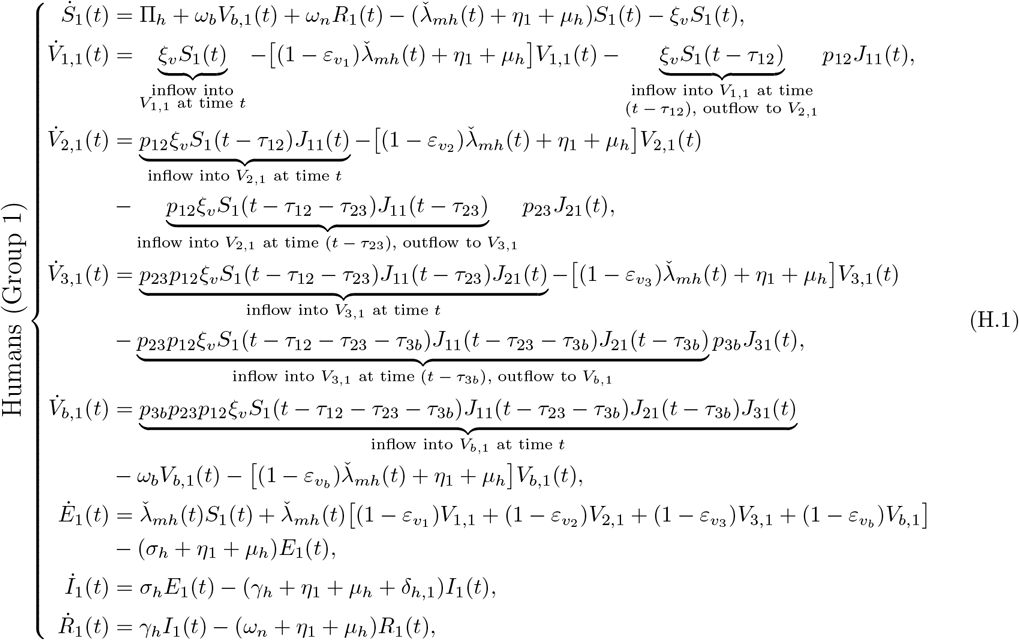

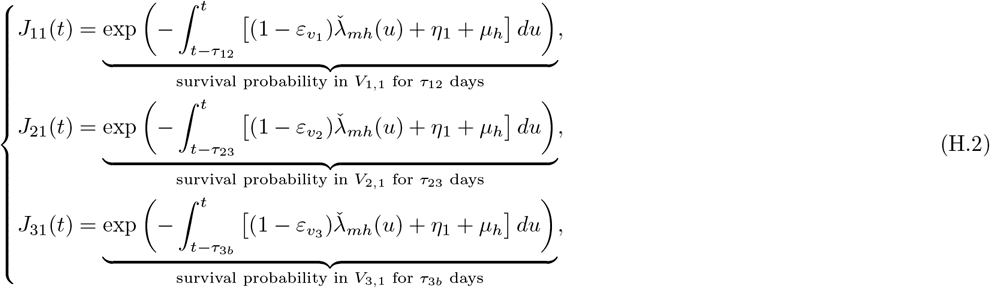

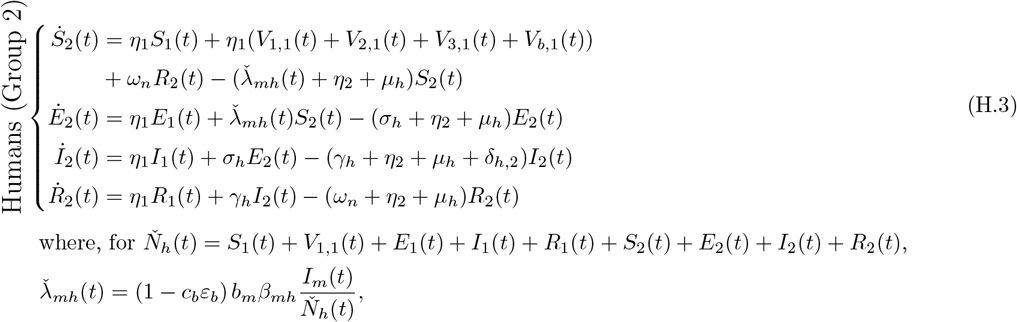

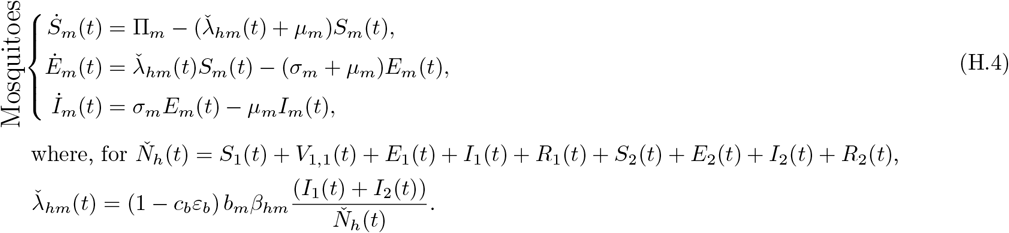

## I Equations for the Special Case of the Model: First Dose Vaccination in Groups 1 and 2

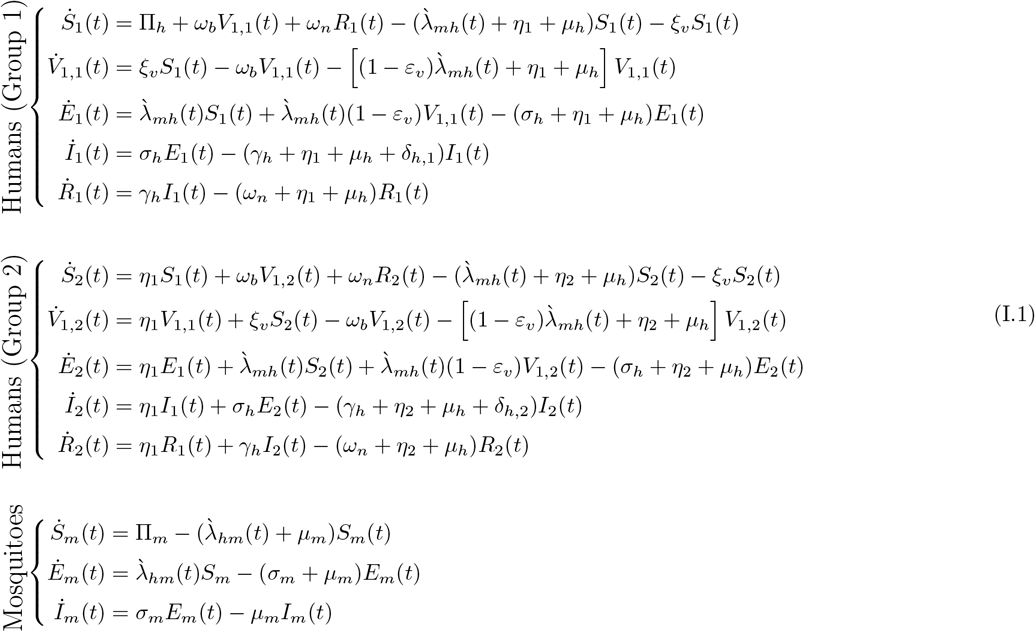

## J Partial Rank Correlation Coefficients (PRCCs)

